# Self-Reported Financial Difficulties Among Patients with Multiple Myeloma and Chronic Lymphocytic Leukemia Treated at U.S. Community Oncology Clinics (Alliance A231602CD)

**DOI:** 10.1101/2024.09.13.24311098

**Authors:** Rena M. Conti, Shaylene McCue, Travis Dockter, Heather J. Gunn, Stacie B. Dusetzina, Antonia V. Bennett, Bruce Rapkin, Gabriela Gracia, Shelley Jazowski, Michelle Johnson, Robert Behrens, Paul Richardson, Niveditha Subbiah, Selina Chow, George J. Chang, Heather B. Neuman, Elisa S. Weiss

## Abstract

**Objectives:** To estimate the proportion and correlates of self-reported financial difficulty among patients with multiple myeloma (MM) or chronic lymphocytic leukemia (CLL).

**Setting:** 23 U.S. community and minority oncology practice sites affiliated with the National Cancer Institute Community Oncology Research Program (NCORP).

**Participants:** 521 patients (≥18 years) with MM or CLL were consented and 416 responded to a survey (completion rate=79.8%). Respondents had a MM diagnosis (74.0%), an associate degree or higher (53.4%), were White (89.2%), insured (100%) and treated with clinician-administered drugs (68.0%).

**Interventions:** Observational, prospective, protocol-based survey administered in 2019-2020.

**Primary and secondary outcome measures:** Financial difficulty was assessed using a single-item standard measure, the EORTC QLQC30: “Has your physical condition or medical treatment caused you financial difficulties in the past year?” and using an ‘any-or-none’ composite measure of 22 items assessing financial difficulty, worries and the use of cost-coping strategies. Multivariable logistic regression models assessed the association between financial difficulty, diagnosis, and socioeconomic and treatment characteristics.

**Results:** 16.8% reported experiencing financial difficulty using the single-item measure and 60.3% using the composite measure. Most frequently endorsed items in the composite measure were financial worry about having to pay large medical bills related to cancer and difficulty paying medical bills. Financial difficulty using the single-item measure was associated with having MM versus CLL (adjusted odds ratio [aOR], 0.34; 95% CI, 0.13-0.84; *P*=.02), having insurance other than Medicare (aOR, 2.53; 95% CI, 1.37-4.66; *P*=.003), being non-White (aOR, 2.21; 95% CI, 1.04-4.72; *P*=.04), and having a high school education or below (aOR, 0.36; 95% CI, 0.21-0.64; *P*=.001). Financial difficulty using the composite measure was associated with having a high school education or below (aOR, 0.62; 95% CI, 0.41-0.94; *P*=.03).

**Conclusions:** U.S. patients with blood cancer report financial difficulty, especially those with low socio-economic status. Evidence-based and targeted interventions are needed.

**Study Strengths and Limitations:** *Strengths:*

- NCORP, a program of the National Cancer Institute (NCI), is a national network for cancer clinical trials and care delivery studies that is comprised of 7 research bases and 46 community sites across the U.S., 14 of which are designated as Minority/Underserved community sites. The study had strong engagement and participation across diverse NCORP Sites across the country and their affiliates. Strong site engagement resulted in high patient recruitment and retention rates for this study (79.8%), despite coincident timing with the initial stages of the COVID-19 pandemic.
- The survey tool was composed of previously validated items that were modified for this population and new questions that were evaluated for comprehension, which facilitates comparison of our findings to others previously published. Patients targeted for recruitment were treated in the community and recruitment aimed to represent the socioeconomic characteristics of the prevalent patient population. The study relied on both self-report and medical chart abstraction to establish key dependent and independent variables.
- The primary outcome variable was a previously developed, and empirically tested measure, supporting study internal and external validity. We also used a secondary composite measure of financial difficulty to present a more holistic picture of how cancer diagnosis and treatment impact patients’ daily lives and inform decisions to delay or forego care and use cost-coping strategies. By capturing specific worries, decisions and strategies, the composite measure indicates areas where there is a need for greater patient engagement and resource provision at the site of care.

*Limitations:*

- Among 105 participants who were not included in the final sample, the majority (n=66) were excluded from analysis because they could not be contacted within the 8-week period due to unexpectedly high recruitment volumes across sites.
- While NCORP sites from across the country participated in the study, 60% of patients recruited to the study were from the Midwest. Although this is the most representative study of financial difficulty in U.S. blood cancer patients to date, our findings may not be fully generalizable to the national CLL and MM communities due to these limitations.

## Background

The blood cancers multiple myeloma (MM) and chronic lymphocytic leukemia (CLL) represent a small percentage of all cancers in the United States (U.S.); however, their treatment costs are among the highest.^1–4^ While treatment advances, including several new high-cost prescription drugs, have resulted in greater survivorship and improved quality of life, out-of-pocket costs and financial difficulties encountered over the course of a cancer diagnosis and prolonged treatment are a growing concern among patients, their families, physicians and national multi-stakeholder groups.^5–12^

Reports suggest that U.S. patients with cancer may be at risk of treatment-related financial difficulty.^13–16^ Specifically, patients younger than 65, with lower household incomes and financial literacy scores, people of color, and those living in rural areas are more likely to experience financial difficulty.^17–21^ Financial difficulty includes an inability to pay for basic necessities such as food and utilities, as well as the presence of medical debt and high out-of-pocket spending relative to income.^22^ Financial difficulty is associated with various cost-coping strategies including skipping medication, taking less medication or not filling a prescription.^6,23–26^

Yabroff et al.^27^ offered a model of financial difficulty founded on the assumption that patients with cancer face a decision to be treated based on an expectation of the benefits and costs of treatment. Self-reported financial difficulty each year can occur because a patient elects to be treated, but lacks sufficient resources to manage the expense of treatment.^28–30^ Yabroff suggested that there are a range of factors, including policy, practice, provider and patient characteristics, which interact to shape patients’ experiences of financial difficulty and their behaviors to cope with difficulty. Patients with MM or CLL cannot be cured and have a protracted course of treatment which is associated with decrements in quality of life.^31,32^ Treatments are progressive and multimodal and often include diagnostic monitoring, frequent physician visits, use of expensive clinician or orally-administered drugs, hospitalizations to address adverse events associated with diagnosis and treatment, and, if necessary, stem cell transplantation, CAR-T therapy and associated care. The coordination and costs of these activities may deplete patients’ and their families’ financial resources, interfere with their ability to work, and make it difficult to afford other necessities.^33,34^

This Alliance/NCI protocol-based study sought to describe self-reported financial difficulty among U.S. patients with MM or CLL and identify factors associated with such difficulty. Our pre-specified hypotheses were that patient self-reported financial difficulty is associated with diagnosis, treatment and socioeconomic characteristics including patient sex, race and ethnicity, education and the presence and type insurance coverage.

## Methods

### Study Design

A231602CD was a multi-centered observational, prospective study of patients diagnosed with MM or CLL and receiving treatment at a National Cancer Institute Community Oncology Research Program (NCORP) site (clinicaltrials.gov study identifier NCT03870633). NCORP, a program of the National Cancer Institute (NCI), is a national network for cancer clinical trials and care delivery studies that is comprised of 7 research bases and 46 community sites across the U.S., 14 of which are designated as Minority/Underserved community sites. NCORP sites are consortia of researchers, hospitals, practices, medical centers, and other groups that provide healthcare services.^35^ The study was administered through the Alliance for Clinical Trials in Oncology research base, and the NCI Central Institutional Review Board (CIRB) served as the IRB of record. All NCORP community sites were invited to participate in the study. NCORP site staff recruited patients based on study eligibility criteria and a limited medical record review, and they obtained written, remote, verbal, or electronic consent. Participants were mailed a $20 gift card upon completion of the 60-minute telephone survey administered by the study team.

Between March 2019 and January 2021, 521 patients from 23 NCORP sites and their 66 affiliated locations were registered to the study. Participating sites and the number of patients they accrued are listed in **eTable 1** in the Supplement.

### Participating Patients

Study eligibility was restricted to adult patients (≥18 years of age) who (1) had been prescribed or recommended to receive drug-based anticancer therapy, whether administered orally or by infusion, within the prior 12 months; (2) were not currently enrolled in a clinical trial in which a drug was supplied; and (3) were able to read and comprehend English or Spanish.

Patients with a psychiatric illness or other mental impairment that would preclude their ability to give informed consent or respond to the telephone survey were excluded from the study.

### Survey Design

The study design was approved by the Alliance/NCI and conducted per protocol. The study used a comprehensive and theoretically grounded patient financial assessment survey comprising multiple domains, including financial difficulty, patient socioeconomic indicators, and health and well-being (eAppendix). Most questionnaire items were from validated national surveys, or from well-established patient-reported outcome instruments. Items developed or modified for this survey were pilot tested among patients with MM or CLL. All items had closed-ended responses, with recall periods of either “now” or “in the past 12 months.” In some cases, the recall period of previously published items was modified to accommodate the study design; these changes were reviewed by a survey methodologist in accordance with guidelines published by Stull et al.^36^

Self-reported socioeconomic information included sex, race, education level, and insurance type. Additionally, NCORP site staff abstracted the following MM or CLL related information from the medical records of consented patients: date of diagnosis, treatment history and current treatments, including treatment initiation dates. Non-cancer related information included date or birth and current comorbidities. The medical abstraction applied to the past 12 months, with some exceptions, such as date of diagnosis, which may have been outside this window. Exposure to clinician-administered anticancer drugs was also measured (see **eTable 2** in the Supplement for full list of anticancer drugs that were considered clinician-administered and their frequency of use).

### Outcome Measures

Financial difficulty was assessed using a single-item from the European Organization for Research and Treatment of Cancer Quality of Life Questionnaire – Core 30^37–39^ (EORTC QLQ-C30) with a modified recall period: “Has your physical condition or medical treatment caused you financial difficulties in the past year?” The EORTC-QLQ-C30 is a cancer specific instrument developed three decades ago by international researchers. ^1^ Due to the salience of financial difficulties and the typically long duration for chronically ill patients, a 12-month recall period was appropriate and based on theoretical work on recall periods.^36^ Based on previously published scoring of the instrument and per Alliance/NCI approved study protocol in the analytical models, participant responses were dichotomized (i.e., “Not at all” or “A little” classified as No, “Quite a bit” and “Very much” classified as Yes).

Also in pre-planned analyses approved in the Alliance/NCI study protocol, a composite measure was created to capture additional aspects of financial difficulty. The composite measure included the following topics based on theoretical relevance (see **eAppendix** in the Supplement for full details): Difficulty Paying Medical Bills (Questions 1, 2, 21), Delaying or Foregoing Medical Care (Questions 4, 6, 6a-6f), Financial Worry (Questions 9, 10, 11, 12, 20), Cost- Coping Strategies (Questions 16a-c, 16f), and Treatment-Related Debt or Bankruptcy (Questions 17 and 18). Each question was designed to be answered by respondents as either yes/no. To determine the number of factors underlying these 22 items, an exploratory factor analysis and a scree test was performed;^40^ the latter identifies the “elbow” of a scree plot and retains all factors above the elbow. The scree plot (**eFigure** in the Supplement) supported a unidimensional construct and the one-factor model had acceptable fit,χ^2^(209)=501.93, *P*<.001, RMSEA=0.059, CFI=0.96. If the patient endorsed any of the 22 items, then they were categorized in a binary variable as having financial difficulty per the composite measure. In order to facilitate ease of interpretation of the composition measure, an ‘any-or-none’ scoring was applied.

### Statistical Analysis

Patient socioeconomic characteristics were examined descriptively overall, by cancer type, and according to report of financial difficulty, based on responses to the single-item measure and the composite measure. As all patients completed the survey in English, language was not included among the covariates examined.

Two multivariable logistic regression models were estimated separately for the two outcomes to assess the associations between patient characteristics and financial difficulty. The predictors for both models included sex (male/female), race (White/non-White), cancer type (MM/CLL), comorbidity (Charlson score=0/Charlson score≥1), treatment (not clinician-administered/clinician-administered), education (High School Diploma, GED, or below/above High School Diploma), and insurance presence (presence or absence) and insurance type (Medicare/Medicare + Other/Other). Adjusted and unadjusted odds ratios (aORs and ORs, respectively), confidence intervals (CIs), and *P* values were calculated for each predictor. We used listwise deletion to account for missing data.

The relationship between the single-item measure and composite measure of financial difficulty was examined by calculating the proportion and CI of patients endorsing each item in the composite measure for the total sample, split by patients who endorsed the single-item measure and those who did not, and split by MM and CLL diagnosis.

For all models and comparisons, 2-sided α=.05 was used to determine statistical significance with no adjustment for multiplicity. All analyses were performed by the Alliance Statistics and Data Center using SAS ® version 9.4 with data frozen on October 6, 2021. STROBE reporting guidelines were followed.

## Results

Of the 521 patients consented to the study, 416 completed all or part of the Patient Survey, for a 79.8% completion rate (**Figure 1**). Patients were enrolled from 23 NCORP sites and their 66 affiliated locations.

**Figure 1:**
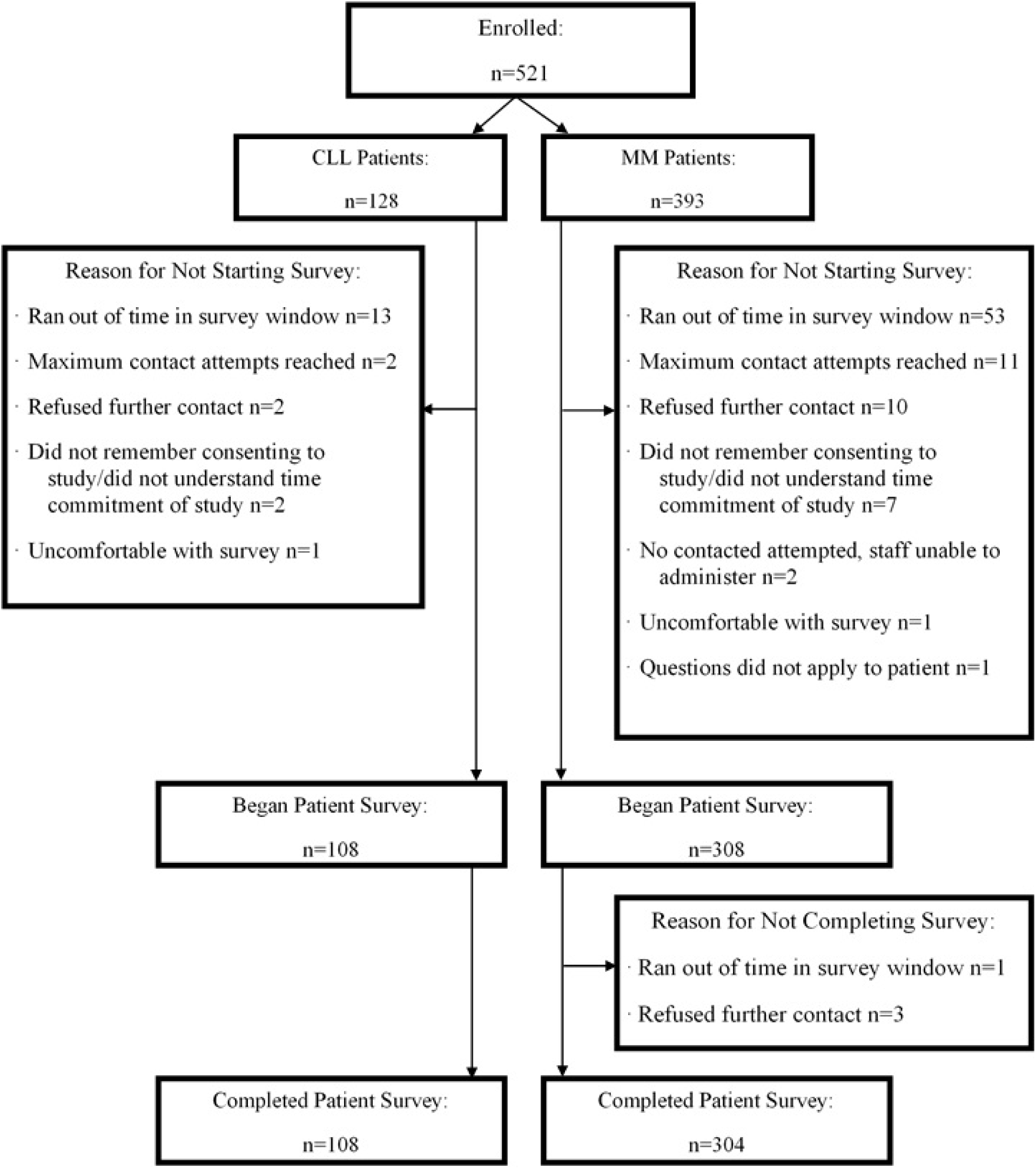
Consort Diagram.

Patients with MM represented 75% (n=308) of the full sample (n=416). Among MM and CLL patients, most respondents were male (56.5%; n=235), White (89.2%; n=371), had an associate degree or higher (54.3%; n=222/409), and were treated with a clinician-administered therapeutic (68.0%; n=283) (**Table 1**). All patients reported they were insured (**Table 1**).

**Table 1.** Socioeconomics and Treatment Characteristics of the 416 Patients Who Responded to All or Part of the Patient Survey.

| **Characteristics** | **MM**<br>**No. (%)** | **CLL**<br>**No. (%)** | **Total**<br>**No. (%)** | **P value<sup>a</sup>** |
| --- | --- | --- | --- | --- |
| No. of patients | 308 | 108 | 416 | NA |
| Age in years, Mean (SD) | 67.5 (9.79) | 71.2 (8.01) | 68.5 (9.49) | <.001<sup>b</sup> |
| Sex |  |  |  | .04 |
| Female | 143 (46.4) | 38 (35.2) | 181 (43.5) |  |
| Male | 165 (53.6) | 70 (64.8) | 235 (56.5) |  |
| Race |  |  |  | .33 |
| White | 272 (88.3) | 99 (91.7) | 371 (89.2) |  |
| Non-White<sup>c</sup> | 36 (11.7) | 9 (8.3) | 45 (10.8) |  |
| Geographic Region<sup>d</sup> |  |  |  | .34 |
| Northeast | 20 (6.5) | 9 (8.3) | 29 (7.0) |  |
| Midwest | 195 (63.3) | 59 (54.6) | 254 (61.1) |  |
| South | 78 (25.3) | 31 (28.7) | 109 (26.2) |  |
| West | 15 (4.9) | 9 (8.3) | 24 (5.8) |  |
| Education |  |  |  | .21 |
| High School Diploma/GED or below | 132 (43.9) | 55 (50.9) | 187 (45.7) |  |
| Above High School | 169 (56.1) | 53 (49.1) | 222 (54.3) |  |
| Missing | 7 | 0 | 7 |  |
| Home Ownership Status |  |  |  | .31 |
| Homeowner | 255 (83.6) | 96 (89.7) | 351 (85.2) |  |
| Non-Homeowner | 50 (16.4) | 11 (10.3) | 61 (14.8) |  |
| Missing | 3 | 1 | 4 |  |
| Reported Household Income |  |  |  | .87 |
| Less than \$20,000 | 40 (14.4) | 10 (10) | 50 (13.3) | |
| \$20,000 to \$39,999 | 52 (18.8) | 19 (19) | 71 (18.9) | |
| \$40,000 to \$59,999 | 49 (17.7) | 22 (22) | 71 (18.9) | |
| \$60,000 to \$79,999 | 42 (15.2) | 15 (15) | 57 (15.2) | |
| \$80,000 to \$99,999 | 30 (10.8) | 10 (10) | 40 (10.6) | |
| \$100,000 or more | 64 (23.1) | 23 (23) | 87 (23.1) | |
| Missing | 31 | 9 | 40 |  |
| Insurance Type |  |  |  | .03 |
| Medicare only | 18 (5.8) | 5 (4.6) | 23 (5.5) |  |
| Medicare + other insurance | 195 (63.3) | 83 (76.9) | 278 (66.8) |  |
| Other<sup>e</sup> | 95 (30.8) | 20 (18.5) | 115 (27.6) |  |
| Charlson Comorbidity Index score |  |  |  | .94 |
| 1 or more | 110 (35.7) | 39 (36.1) | 149 (35.8) |  |
| 0 | 198 (64.3) | 69 (63.9) | 267 (64.2) |  |
| Treatment Received |  |  |  | <.001 |
| Clinician-Administered Therapeutic | 254 (82.5) | 29 (26.9) | 283 (68.0) |  |
| Not a Clinician-Administered Therapeutic | 54 (17.5) | 79 (73.1) | 133 (32.0) |  |
| Single-item Measure of Financial Difficulty |  |  |  | .002 |
| Yes | 62 (20.1) | 8 (7.4) | 70 (16.8) |  |
| No | 246 (79.9) | 100 (92.6) | 346 (83.2) |  |
| Composite Measure of Financial Difficulty |  |  |  | .62 |
| Yes | 188 (61.0) | 63 (58.3) | 251 (60.3) |  |
| No | 120 (39.0) | 45 (41.7) | 165 (39.7) |  |

Abbreviations: NA: Not Applicable, MM: Multiple Myeloma, CLL: Chronic Lymphocytic Leukemia.
<sup>a</sup> All *P* values come from a Chi-Square test unless otherwise noted.
<sup>b</sup> Kruskal-Wallis *P* value.
<sup>c</sup> Non-White race captures patients who selected American Indian or Alaska Native, Asian, Black, or African American, Native Hawaiian, or other Pacific Islander, Unknown, Not Reported
<sup>d</sup> Geographic Regions were divided according to the United States census divisions and regions: **Northeast:** Maine, New Hampshire, Vermont, Massachusetts, Rhode Island, Connecticut, New York, New Jersey, and Pennsylvania; **Midwest:** Ohio, Michigan, Indiana, Wisconsin, Illinois, Minnesota, Iowa, Missouri, North Dakota, South Dakota, Nebraska, and Kansas; **South:** Delaware, Maryland, Virginia, West Virginia, Kentucky, North Carolina, South Carolina, Tennessee, Georgia, Florida, Alabama, Mississippi, Arkansas, Louisiana, Texas, and Oklahoma, Washington DC; **West:** Montana, Idaho, Wyoming, Colorado, New Mexico, Arizona, Utah, Nevada, California, Oregon, Washington, Alaska, and Hawaii.
<sup>e</sup> Other includes: Private Insurance, Medicaid, Military Sponsored (including CHAMPUS & TRICARE), Veterans Sponsored, Not Otherwise Specified (NOS), and any combination of those listed here.

Characteristics were similar by cancer types except CLL patients were older (71.2 vs. 67.5; *P*<.001), less likely to currently be taking clinician-administered therapies (26.9% vs. 82.5%; *P*<.001), and more likely to have Medicare plus another form of insurance (76.9% vs. 63.3%; *P*=.03) compared to MM patients.

### Single-item Measure of Financial Difficulty

Across the 416 patients, 16.8% (95% CI, 13.4%-20.8%) endorsed financial difficulty by responding “Quite a bit” or “Very much” to the item “Has your physical condition or medical treatment caused you financial difficulties in the past year?” The socioeconomic characteristics of those who reported financial difficulty based on this single item are summarized in **eTable 3** in the Supplement.

After applying listwise deletion to the seven predictors and outcome variable used in the multivariable logistic regression model, the analytic sample consisted of 408 patients. The multivariable analyses are presented in **Table 2**. Patients with CLL had significantly lower odds of reporting financial difficulty than patients with MM (aOR, 0.34; 95% CI, 0.13-0.84; *P*=.02) as did patients who have above a high school education compared to patients with a high school education or below (aOR, 0.36; 95% CI, 0.21-0.64; *P*=.001). Additionally, patients who did not have Medicare had significantly greater odds of reporting financial difficulty than patients with Medicare plus one other type of insurance (aOR, 2.53; 95% CI, 1.37-4.66; *P*=.003) as did non-White patients compared to White patients (aOR, 2.21; 95% CI, 1.04-4.72; *P*=.04).

**Table 2.** Multivariable Logistic Regression Results of Survey Respondent Self-Report of Financial Difficulty.

| **Model** | **Unadjusted Odds Ratios (95% CI)** | **P value** | **Adjusted Odds Ratios (95% CI)<sup>a</sup>** | **P value** |
| --- | --- | --- | --- | --- |
| **Single-item Measure of Financial Difficulty** |  |  |  |  |
| Female | 1.32 (0.78, 2.21) | .30 | 1.09 (0.62, 1.91) | .77 |
| Non-White | 2.97 (1.49, 5.90) | .002 | 2.21 (1.04, 4.72) | .04 |
| Comorbidity | 1.39 (0.82, 2.36) | .27 | 1.53 (0.86, 2.75) | .15 |
| CLL | 0.31 (0.15, 0.68) | .003 | 0.34 (0.13, 0.84) | .02 |
| Clinician-Administered Treatment | 1.72 (0.94, 3.15) | .08 | 1.02 (0.49, 2.12) | .96 |
| Above HS Education | 0.42 (0.24, 0.71) | .001 | 0.36 (0.21, 0.64) | .001 |
| Medicare + Other<sup>b</sup> | NA [Reference] | NA | NA [Reference] | NA |
| Medicare | 3.17 (1.21, 8.27) | .02 | 2.17 (0.77, 6.07) | .14 |
| Other<sup>c</sup> | 2.50 (1.43, 4.37) | .001 | 2.53 (1.37, 4.66) | .003 |
| **Composite Measure of Financial Difficulty** |  |  |  |  |
| Female | 1.35 (0.90, 2.01) | .15 | 1.28 (0.84, 1.94) | .25 |
| Non-White | 1.84 (0.92, 3.69) | .09 | 1.56 (0.75, 3.23) | .24 |
| Comorbidity | 1.28 (0.84, 1.94) | .29 | 1.27 (0.81, 1.97) | .30 |
| CLL | 0.88 (0.56, 1.37) | .57 | 1.21 (0.71, 2.09) | .48 |
| Clinician-Administered Treatment | 1.52 (0.998, 2.32) | .05 | 1.61 (0.97, 2.68) | .07 |
| Above HS Education | 0.62 (0.41, 0.92) | .02 | 0.62 (0.41, 0.94) | .03 |
| Medicare + Other<sup>b</sup> | NA [Reference] | NA | NA [Reference] | NA |
| Medicare | 3.64 (1.21, 10.98) | .02 | 3.00 (0.98, 9.21) | .06 |
| Other<sup>c</sup> | 1.47 (0.94, 2.32) | .10 | 1.57 (0.97, 2.54) | .07 |

Abbreviations: CLL: Chronic Lymphocytic Leukemia, HS: High School, NA: Not Applicable.
<sup>a</sup> The adjusted models include all predictors listed in the table.
<sup>b</sup> The insurance variable was defined by two dummy variables with patients who have Medicare and another insurance (i.e., Medicare + Other) as the reference group.
<sup>c</sup> Other insurance includes: Private Insurance, Medicaid, Military Sponsored (including CHAMPUS & TRICARE), Veterans Sponsored, Not Otherwise Specified (NOS), and any combination.

As a sensitivity analysis, we investigated other dichotomizations of the single measure.

These results are summarized in **eTable 4**. Additionally, we provide the proportions of endorsement for each category of the primary endpoint for the overall sample and split by diagnosis in **eTable 5**.

### Composite Measure of Financial Difficulty

All respondents who completed the single-item measure also completed the composite measure of financial difficulty (i.e., 416 patients). More than half of respondents (n=251) affirmed at least one of the 22 items in the composite measure of financial difficulty (60.3%; 95% CI, 55.6%-64.9%). The most frequently endorsed item was, “In the past 12 months, have you ever worried about having to pay large medical bills related to your cancer? (Part 1, Question 20).” Other commonly endorsed items include difficulty paying medical bills in general and forgoing dental care. **Table 3** presents the proportion and CI of patients endorsing each of the items in the composite measure of financial difficulty for those patients who endorsed the single-item measure of financial difficulty and for those patients who indicated financial difficulty according to the any-or-none composite.

**Table 3.** Number of Patients Who Endorsed the Items in Composite Measure of Financial Difficulty by Endorsement of Financial Difficulty of Single-item Measure of Financial Difficulty and Composite Measure.

| Survey Question | Endorsed Single-item Measure (n=70)<br>No.<br>Proportion<br>(95% exact CI) | Endorsed One or More Composite Items (n=251)<br>No.<br>Proportion<br>(95% exact CI) |
| --- | --- | --- |
| In the past 12 months, did you have problems paying or were unable to pay any medical bills? Include bills for doctors, hospitals, therapists, medication, equipment, nursing home or home. (Part 1, Question 1) | 49<br>0.700<br>(0.579, 0.804) | 88<br>0.351<br>(0.292, 0.413) |
| Do you or anyone in your family currently have medical bills that you are unable to pay at all? (Part 1, Question 2) | 30<br>0.429<br>(0.311, 0.553) | 47<br>0.187<br>(0.141, 0.241) |
| During the past 12 months, have you or someone in your family delayed medical care because you were worried about the cost (do not include dental care)? (Part 1, Question 4) | 29<br>0.414<br>(0.298, 0.538) | 56<br>0.223<br>(0.173, 0.279) |
| During the past 12 months, was there a time when you or someone in your family needed medical care but didn't get it because you couldn't afford it? (Part 1, Question 5) | 17<br>0.243<br>(0.148, 0.360) | 29<br>0.116<br>(0.079, 0.162) |
| During the past 12 months, was there a time when you needed one of the following, but did not get it because you couldn't afford it? **Prescription medicine** (Part 1, Question 6a) | 18<br>0.257<br>(0.160, 0.376) | 41<br>0.163<br>(0.119, 0.215) |
| During the past 12 months, was there a time when you needed one of the following, but did not get it because you couldn't afford it? **Mental health care or counseling** (Part 1, Question 6b) | 6<br>0.086<br>(0.032, 0.177) | 12<br>0.048<br>(0.025, 0.082) |
| During the past 12 months, was there a time when you needed one of the following, but did not get it because you couldn't afford it? **Dental care** (Part 1, Question 6c) | 33<br>0.471<br>(0.351, 0.595) | 79<br>0.315<br>(0.258, 0.376) |
| During the past 12 months, was there a time when you needed one of the following, but did not get it because you couldn't afford it? **Eyeglasses** (Part 1, Question 6d) | 22<br>0.314<br>(0.209, 0.436) | 46<br>0.183<br>(0.137, 0.237) |
| During the past 12 months, was there a time when you needed one of the following, but did not get it because you couldn't afford it? **Cancer-related medical care** (Part 1, Question 6e) | 13<br>0.186<br>(0.103, 0.297) | 21<br>0.084<br>(0.053, 0.125) |
| During the past 12 months, was there a time when you needed one of the following, but did not get it because you couldn't afford it? **Non-cancer related medical care** (Part 1, Question 6f) | 19<br>0.271<br>(0.172, 0.391) | 31<br>0.124<br>(0.086, 0.171) |
| If you get sicker or have an accident, how worried are you that you will not be able to pay your medical bills? (Part 1, Question 9) | 40<br>0.571 | 66<br>0.263 |
|  | (0.448, 0.689) | (0.209, 0.322) |
| How often in the last 12 months would you say you were worried or stressed about having enough money to pay your rent or mortgage? (Part 1, Question 10) | 42<br>0.600<br>(0.476, 0.715) | 82<br>0.327<br>(0.269, 0.389) |
| How often in the last 12 months would you say you were worried or stressed about having enough money to buy nutritious meals? (Part 1, Question 11) | 37<br>0.529<br>(0.406, 0.649) | 66<br>0.263<br>(0.209, 0.322) |
| How often in the last 12 months would you say you were worried or stressed about having enough money to pay household utilities such as water, gas, and electricity? (Part 1, Question 12) | 41/69<br>0.594<br>(0.469, 0.711) | 74/250<br>0.296<br>(0.240, 0.357) |
| During the past 12 months, were any of the following true for you: **You skipped medication doses to save money** (Part 1, Question 16a) | 17<br>0.243<br>(0.148, 0.360) | 26<br>0.104<br>(0.069, 0.148) |
| During the past 12 months, were any of the following true for you: **You took less medicine to save money** (Part 1, Question 16b) | 16<br>0.229<br>(0.137, 0.345) | 29<br>0.116<br>(0.079, 0.162) |
| During the past 12 months, were any of the following true for you: **You delayed filling a prescription to save money** (Part 1, Question 16c) | 28<br>0.400<br>(0.285, 0.524) | 46<br>0.183<br>(0.137, 0.237) |
| During the past 12 months, were any of the following true for you: You used alternative therapies to save money (Part 1, Question 16f) | 4<br>0.057<br>(0.016, 0.139) | 18<br>0.072<br>(0.043, 0.111) |
| In the past 12 months, have you or has anyone in your family had to borrow money or go into debt because of your cancer, its treatment, or the lasting effects of that treatment? (Part 1, Question 17) | 31<br>0.443<br>(0.324, 0.567) | 47<br>0.187<br>(0.141, 0.241) |
| In the past 12 months, did you or your family file for bankruptcy because of your cancer, its treatment, or the lasting effects of that treatment? (Part 1, Question 18) | 2<br>0.029<br>(0.004, 0.099) | 3/250<br>0.012<br>(0.003, 0.035) |
| In the past 12 months, have you ever worried about having to pay large medical bills related to your cancer? (Part 1, Question 20) | 60<br>0.857<br>(0.753, 0.929) | 173<br>0.689<br>(0.628, 0.746) |
| Please think about medical care visits for cancer, its treatment, or the lasting effects of that treatment in the past 12 months. Have you ever been unable to cover your share of those visits? (Part 1, Question 21) | 26<br>0.371<br>(0.259, 0.495) | 40<br>0.159<br>(0.116, 0.211) |

Note: Each cell presents the following information: number of patients who answered yes or always/usually/sometimes, the proportion, and the 95% Wilson score confidence interval. If there was missing data, the total number of patients who answered the question was also provided.

The socioeconomic characteristics of those who endorsed the composite measure of financial difficulty are summarized in **eTable 3** in the Supplement. **eTable 6** in the Supplement presents the proportion of patients endorsing each of the items in the composite measure of financial difficulty split by cancer type. For most items, a greater proportion of MM patients endorsed the item compared to CLL patients.

The logistic regression with the composite measure of financial difficulty as the outcome is presented in **Table 2** for the 408 patients with complete data. Education was significantly associated with the composite measure of financial difficulty such that patients with above a high school education had lower odds of reporting financial difficulty compared to patients with a high school education or below (aOR, 0.62; 95% CI, 0.41-0.94; *P*=.03).

### Relation Between Single-item Measure and Composite Measure of Financial Difficulty

The single-item measure and the any-or-none composite measure of financial difficulty were dependent such that all 70 patients who endorsed the single-item measure endorsed one or more items of the composite measure. As a sensitivity analysis, we investigated differences between patients who indicated financial difficulty according to both the single-item measure and composite measure and those who did not. We created 3 non-overlapping groups: endorsed the single-item measure, endorsed the composite but not the single-item measure, and did not endorse either measure. We tested for statistical differences in patient sociodemographic and comorbidity information by this 3-group categorization and the results are presented in **Table 4** with the corresponding *P* value. If we assume endorsing the single-item measure indicates the most financial difficulty and not endorsing either measure indicates the least, then as financial difficulty increased, patients were more likely to be younger, be female, be non-White, be less educated, more likely to have MM rather than CLL, and more likely to have some form of insurance other than Medicare.

**Table 4.** Patient Demographics and Treatment Characteristics by Self Report of Financial Difficulty by Measure.

| Variable | No. of Patients (%) |  |  | P value<sup>a</sup> |
| --- | --- | --- | --- | --- |
|  | Endorsed<br>Single-item Measure | Endorsed<br>Composite Only | Endorsed<br>Neither |  |
| No. of Patients | 70 | 181 | 165 |  |
| Age in years, Mean (SD) | 64.3 (11.09) | 68.0 (9.05) | 70.8 (8.58) | <.001<sup>b</sup> |
| Sex |  |  |  | .43 |
| Female | 34 (49) | 81 (45) | 66 (40) |  |
| Male | 36 (51) | 100 (55) | 99 (60) |  |
| Race |  |  |  | .007 |
| White | 55 (79) | 164 (91) | 152 (92) |  |
| Non-White<sup>c</sup> | 15 (21) | 17 (9) | 13 (8) |  |
| Geographic Region<sup>d</sup> |  |  |  | .69 |
| Northeast | 5 (7) | 11 (6) | 13 (8) |  |
| Midwest | 42 (60) | 111 (61) | 101 (61) |  |
| South | 19 (27) | 52 (29) | 38 (23) |  |
| West | 4 (6) | 7 (4) | 13 (8) |  |
| Education |  |  |  | .002 |
| High School Diploma/GED or below | 44 (64) | 81 (45) | 62 (39) |  |
| Above High School | 25 (36) | 98 (55) | 99 (61) |  |
| Missing | 1 | 2 | 4 |  |
| Home Ownership Status |  |  |  | .04 |
| Homeowner | 52 (75) | 155 (86) | 144 (89) |  |
| Non-Homeowner | 17 (25) | 26 (14) | 18 (11) |  |
| Missing | 1 | 0 | 3 |  |
| Reported Household Income |  |  |  | <.001 |
| Less than \$20,000 | 19 (29) | 24 (15) | 7 (5) | |
| \$20,000 to \$39,999 | 18 (28) | 37 (22) | 16 (11) | |
| \$40,000 to \$59,999 | 16 (25) | 35 (21) | 20 (14) | |
| \$60,000 to \$79,999 | 6 (9) | 20 (12) | 31 (21) | |
| \$80,000 to \$99,999 | 4 (6) | 14 (8) | 22 (15) | |
| \$100,000 or more | 2 (3) | 36 (23) | 49 (34) | |
| Missing | 5 | 15 | 20 |  |
| Insurance Type |  |  |  | .003 |
| Medicare only | 7 (10) | 12 (7) | 4 (3) |  |
| Medicare + other insurance | 34 (49) | 123 (68) | 121 (73) |  |
| Other<sup>e</sup> | 29 (41) | 46 (25) | 40 (24) |  |
| Charlson Comorbidity Index score |  |  |  | .36 |

| Variable | No. of Patients (%) |  |  | *P* value<sup>a</sup> |
| --- | --- | --- | --- | --- |
|  | Endorsed<br>Single-item Measure | Endorsed<br>Composite Only | Endorsed<br>Neither |  |
| 1 or more | 29 (41) | 67 (37) | 53 (32) |  |
| 0 | 41 (59) | 114 (63) | 112 (68) |  |
| Treatment Received |  |  |  | .10 |
| Clinician-Administered Therapeutic | 54 (77) | 125 (69) | 104 (63) |  |
| Not a Clinician-Administered Therapeutic | 16 (23) | 56 (31) | 61 (37) |  |
| Diagnosis |  |  |  | .008 |
| Multiple Myeloma | 62 (89) | 126 (70) | 120 (73) |  |
| Chronic Lymphocytic Leukemia | 8 (11) | 55 (30) | 45 (27) |  |

<sup>a</sup> All *P* values come from a Chi-Square test unless otherwise noted.
<sup>b</sup> Kruskal-Wallis *P* value.
<sup>c</sup> White race captures patients who did not select American Indian or Alaska Native, Asian, Black, or African American, Native Hawaiian, or other Pacific Islander, Unknown, Not Reported.
<sup>d</sup> Geographic Regions were divided according to the United States census divisions and regions: **Northeast:** Maine, New Hampshire, Vermont, Massachusetts, Rhode Island, Connecticut, New York, New Jersey, and Pennsylvania; **Midwest:** Ohio, Michigan, Indiana, Wisconsin, Illinois, Minnesota, Iowa, Missouri, North Dakota, South Dakota, Nebraska, and Kansas; **South:** Delaware, Maryland, Virginia, West Virginia, Kentucky, North Carolina, South Carolina, Tennessee, Georgia, Florida, Alabama, Mississippi, Arkansas, Louisiana, Texas, and Oklahoma, Washington DC; **West:** Montana, Idaho, Wyoming, Colorado, New Mexico, Arizona, Utah, Nevada, California, Oregon, Washington, Alaska, and Hawaii.
<sup>e</sup> Other includes: Private Insurance, Medicaid, Military Sponsored (including CHAMPUS & TRICARE), Veterans Sponsored, Not Otherwise Specified (NOS), and any combination of those listed here.

## Discussion

In this large, multi-centered protocol-based study of MM and CLL patients conducted across 23 NCORP sites and their 66 affiliates, 16.8% specifically reported experiencing financial difficulty over the past 12 months using a single-item measure, while 60.3% endorsed financial difficulty items using a composite measure with questions about financial worry and difficulty paying medical bills eliciting the most affirmative responses. All survey respondents were insured, largely by Medicare. Some were insured by a primary and secondary payer, indicating less potential out of pocket costs associated with cancer treatment. Respondents endorsing both the single-item and composite measure of financial difficulty were significantly more likely to report having less education. For the single-item measure, reports of financial difficulty were also significantly associated with having MM, having insurance other than Medicare, and being non-White. Patients with MM tended to have higher endorsement on all items in the composite measure of financial difficulty.

This is the first multi-center study to assess the proportion of patients with financial difficulties and its correlates among MM and CLL patients treated at NCI NCORP sites and reports significant financial difficulties among patients who have been prescribed treatment. The findings are important, because although general financial difficulties associated with cancer treatment are well-documented,^13–15,27,43,44^ they are still considered to be underreported, and a comprehensive understanding of the range of financial difficulty types, worries and cost-coping strategies among this specific population has been, to date, limited.^45^ The findings of this study are also concerning for patients with blood cancers, their families, treating physicians, and policymakers. As new treatments provide improvements in patient outcomes, including survival, there has been a corresponding rise in treatment costs.^41^ Compared to CLL, MM treatments often involve multiple, expensive therapies with greater side effects that negatively impact a patient’s ability to work and quality of life.^42^ The results of this study also add to a growing body of evidence suggesting U.S. families are financially vulnerable. The relationships between general financial stress and specific financial burden related to cancer diagnosis and treatment are likely complex. The study endeavored to separate the latter from the former, but further study of these relationships is needed.

This study has several additional strengths. First, we used a survey tool composed of previously validated items that were modified for this population and new questions that were evaluated for comprehension, which facilitates comparison of our findings to others previously published. Findings from this survey are in line with research that demonstrate how experiences of financial difficulty impact the use of cost coping strategies (including treatment nonadherence), ^2^ feelings of distress, ^3^ ^4^ and family members/caregivers. ^5^ Additionally, our questions on support seeking behaviors are supported by various studies that highlight the importance of engaging the care team ^6^ and financial navigators ^7^ in providing support and resources.

Second, our use of the composite measure of financial difficulty sought to present a more holistic picture of how cancer diagnosis and treatment impact patients’ daily lives and inform decisions to delay or forego care and use cost-coping strategies. By capturing specific worries, decisions and strategies, the composite measure indicates areas where there is a need for greater patient engagement and resource provision at the site of care. For instance, patients expressed worry or concern about paying for household utilities, cancer and non-cancer related care and indicated delaying or foregoing treatment for cancer and non-cancer related care in response to cost concerns; they also reported using cost-coping strategies for cancer and non-cancer related care (**Table 3**). Patients may be implementing cost-coping strategies without recognizing the connection between these strategies and the financial difficulty they face.

Third, this study had strong engagement and participation across 23 diverse NCORP Sites across the country and their 66 affiliates. Strong site engagement resulted in high patient recruitment and retention rates for this study (79.8%). Between December 2019 and January 2021, most participants (n=490) were recruited to the study, despite coincident timing with the initial stages of the COVID-19 pandemic.

This study also has several additional limitations. First, among 105 participants who were not included in the final sample, the majority (n=66) were excluded from analysis because they could not be contacted within the 8-week period due to unexpectedly high recruitment volumes across sites. Second, the sample itself was not representative of the national MM and CLL patient populations. Black or African-American patients make up 20.5% of the MM population^32^ and 5.8 % of the CLL population,^32^ respectively. However, in this study, they only accounted for 7.8% of the MM sample and 7.4% of the CLL sample. Although our research team had identified

Minority and Underserved NCORP Sites to participate in the study, seven were unable to recruit prior to the study closure due to site-related issues (e.g., electronic health record conversions, staff shortages) and/or COVID-19 related delays. Third, while NCORP sites from across the country participated in the study, 60% of patients recruited to the study were from the Midwest. Although this is the most representative study of financial difficulty in U.S. blood cancer patients to date, our findings may not be fully generalizable to the national CLL and MM communities due to these limitations, we did not adjust for multiplicity.

Finally, this study focused on patient-level financial difficulties associated with cancer treatments, as we believe cancer-treatment related financial difficulty is at core a patient experience. However, we are aware of and endorse the emphasis made by previous studies of patient-level financial difficulty on the broader context in which care is provided. Specifically, previous studies have emphasized that financial difficulties are also related to practice-level, payer-level and health system-wide characteristics. Our study captured additional practice-level characteristics of the sites enrolling patients into this study, and future planned analyses will focus on those outcomes.

More generally, the present aim of identifying components of patient-level financial difficulty and correlates of financial difficulty was intended to inform the future development of evidence-based and targeted interventions at both the patient and the practice levels. Previous studies have identified barriers to providing and accessing financial navigation services that disproportionately impact vulnerable patient populations, including rural, minority and younger patients.^46–48^ Planned follow-on studies have been designed to better understand patient experiences related to the composite measure topics (e.g., financial worry, use of cost coping strategies) and examine practice and provider engagement with identifying and ameliorating patient financial difficulty. Specific intervention goals may include developing more comprehensive screening practices and counseling to address financial concerns and mitigate the use of cost coping strategies, identifying communication tools to elicit financial concerns and specific preferences for types of assistance from patients, and advanced financial planning to reduce delays in the initiation or continuity of treatment.^14,49,50^

One important implication of our study’s results is that patients who endorsed the single-item measure had a higher proportion of endorsement on items in the composite measure compared to patients who did not (**Table 3**). Moreover, we found that patients who reported financial difficulty using the composite measure did not always report financial difficulty on the single item measure. This suggests that future clinical interventions and research should consider using both the single item and the composite measure elements to screen patients for financial difficulty. Use of the composite measure in this context may also provide a way for physicians and sites to engage in specific discussions with patients and to identify resources that can help patients manage care and non-care related costs.

## Conclusion

This is the first multicenter study to systematically assess the proportion of patients with financial difficulties and its correlates among MM and CLL patients treated at NCI NCORP sites. We found that U.S. patients with blood cancers experience financial difficulty, especially among those with low socioeconomic status. Evidence-based and targeted interventions to mitigate financial difficulty among U.S. blood cancer patients are needed.

## Supporting information

Study Protocol

## Data Availability

All data produced in the present study will be available by request through Alliance/NCI.

## Trial registration

This project was conducted under Alliance/NCI A231602CD “Assessing Financial Difficulty in Blood Cancer Patients.” ClinicalTrials.gov Identifier is NCT03870633.

## Data sharing statement

Data will be made available to the public by request from NC/Alliance.

## Subject MESH Terms

oncology, leukemia, myeloma, health policy, health economics, financial burden, financial toxicity

## Contributorship Statement

Gunn, McCue, Dockter analyzed and interpreted the data, drafted the initial manuscript, and reviewed and revised the manuscript for important intellectual content.

Conti, Dusetzina, Bennett, Weiss, Rapkin, Gracia, Jazowski conceptualized and designed the study, analyzed and interpreted the data, drafted the initial manuscript, reviewed and revised the manuscript for important intellectual content, and provided study supervision.

Behrens, Richardson conceptualized the study, provided study supervision and reviewed and revised the manuscript for important intellectual content.

Subbiah, Chow, Chang, Neuman provided study supervision and reviewed and revised the manuscript for important intellectual content.

All authors approved the final manuscript as submitted and agree to be accountable for all aspects of the work.

## eAppendix: Study Protocol with Patient Survey

**eFigure.**
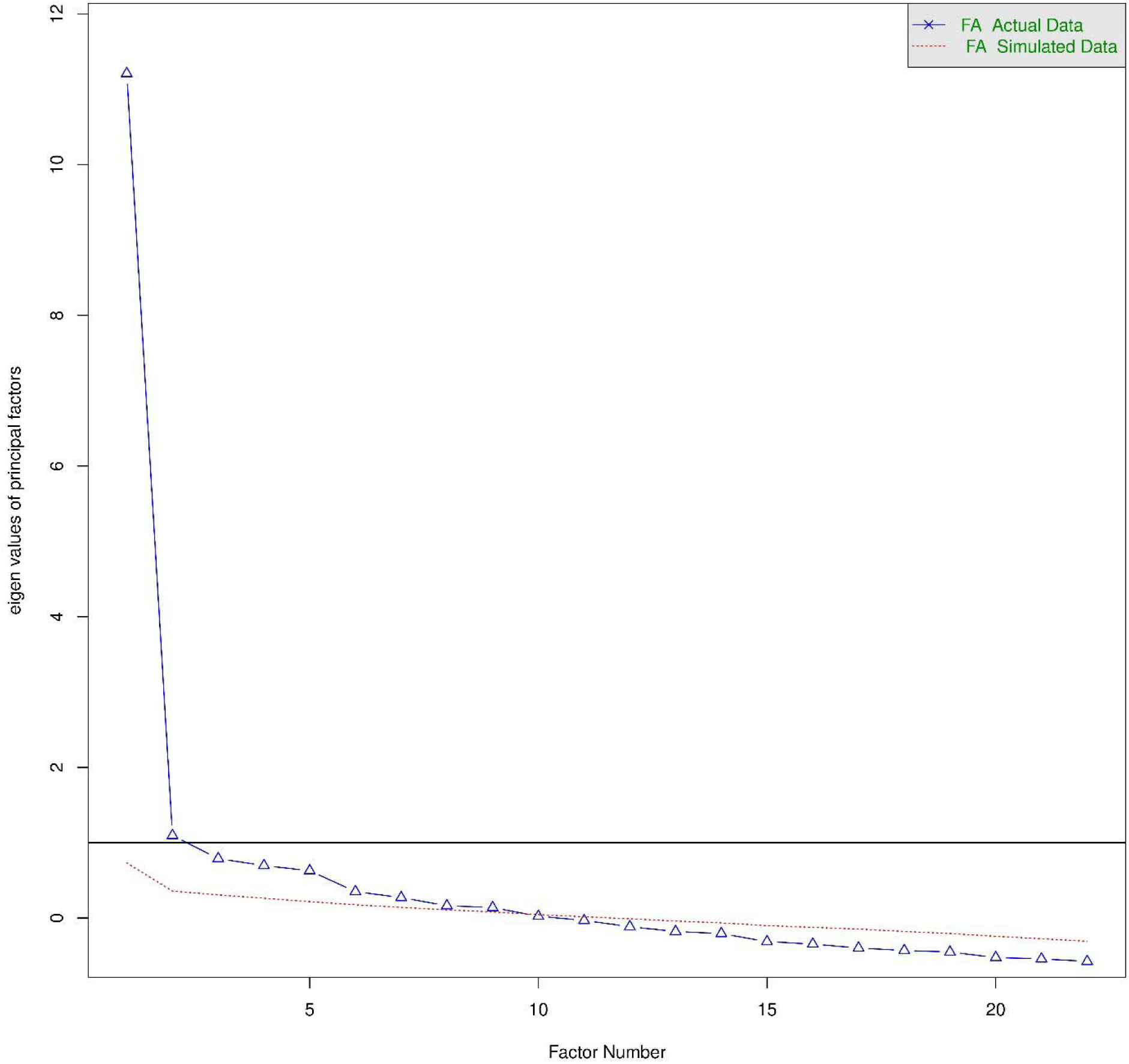
Scree Plot of Composite Measure.

**eTable 1.**
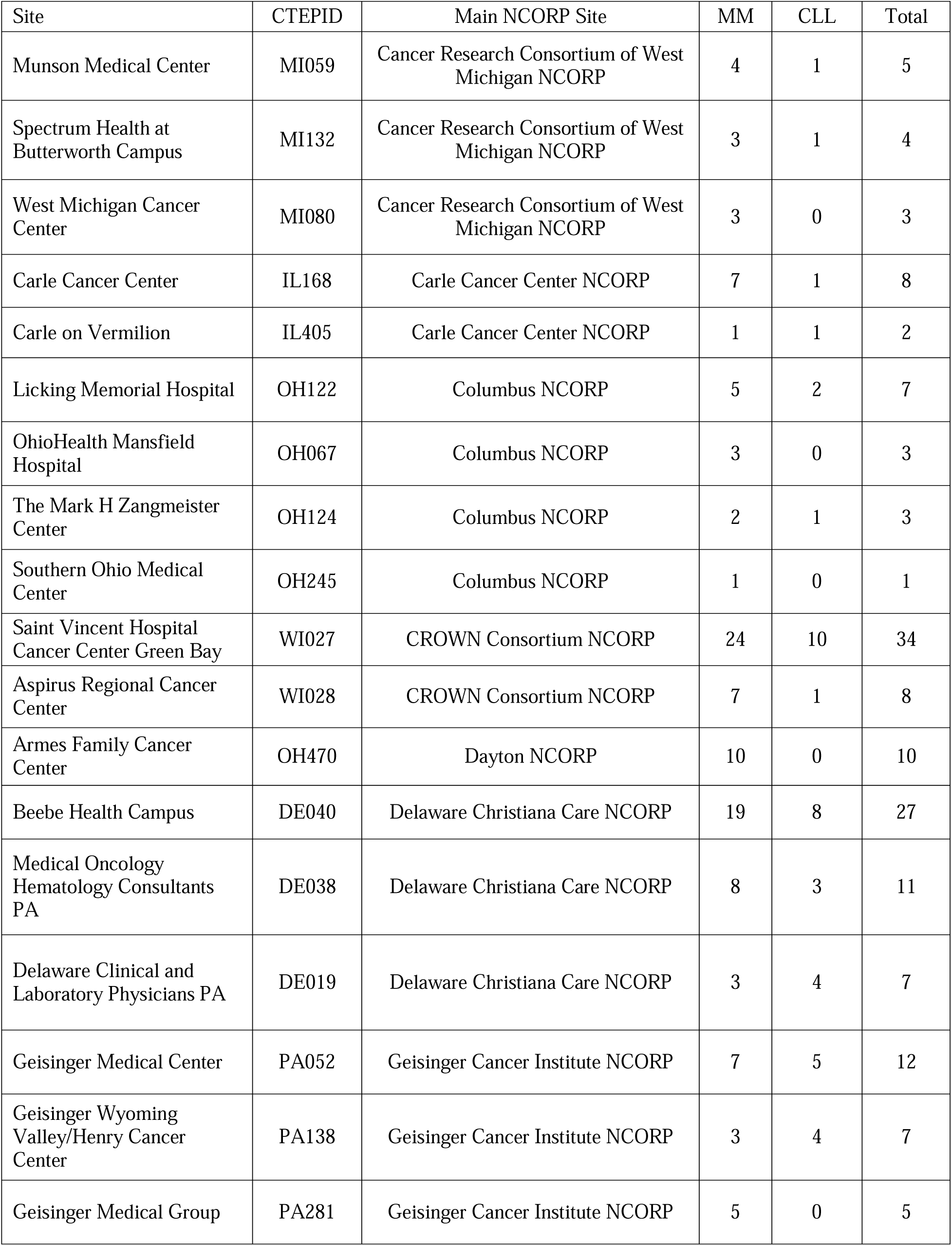
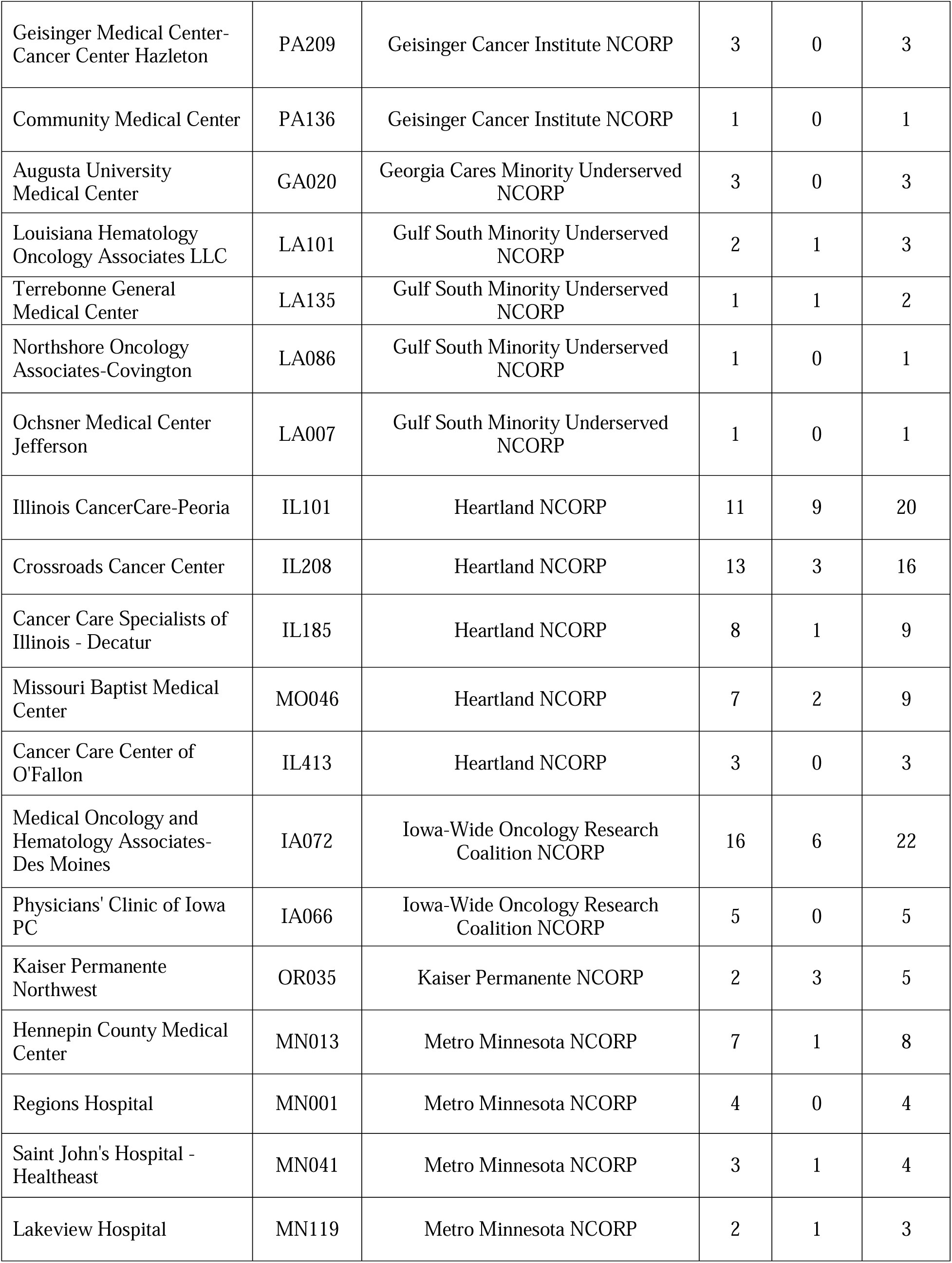
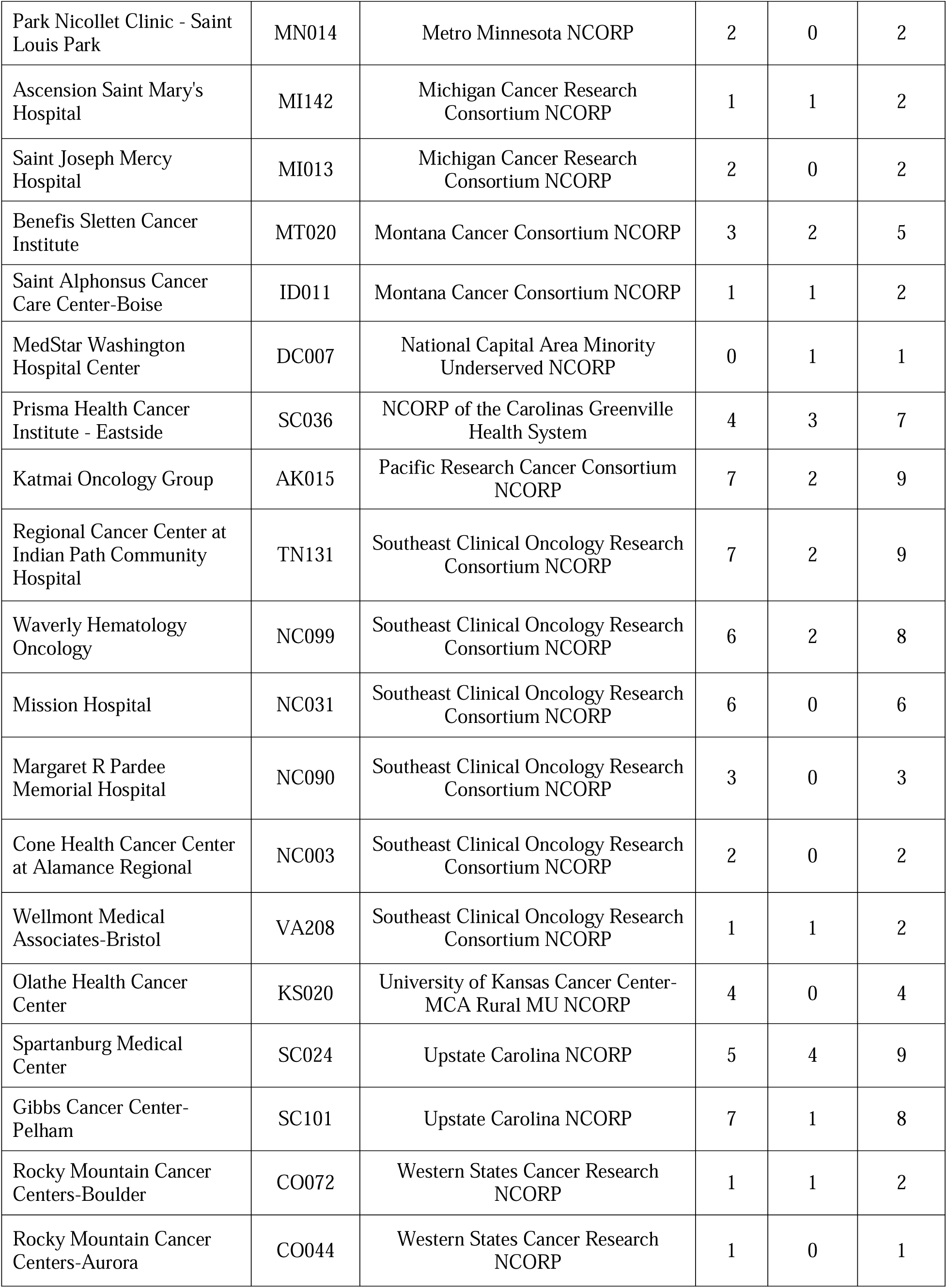
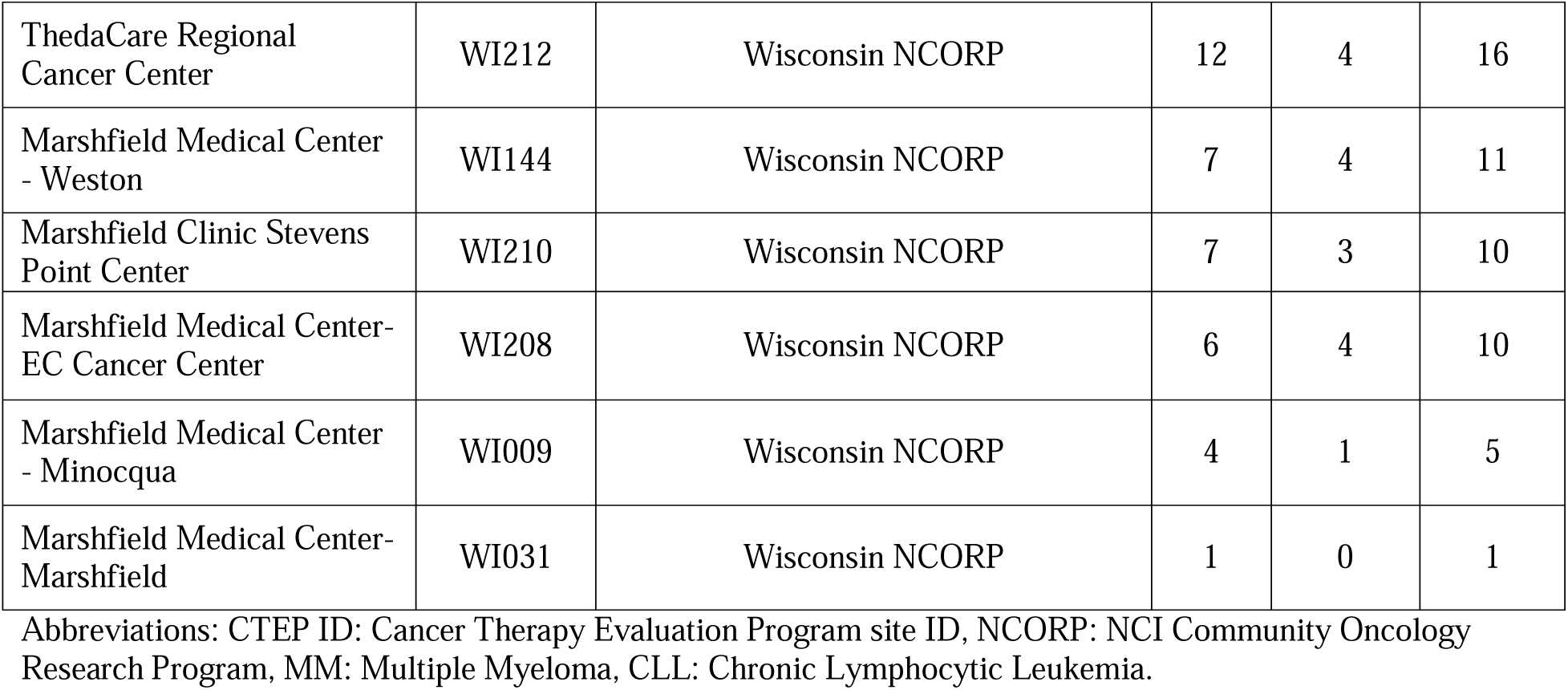
Patient Enrollment by Site and by Diagnosis.

**eTable 2.** Frequency of Clinician-Administered Treatments.

| **Clinician-Administered Treatment** | **Frequency** |
| --- | --- |
| Dexamethasone | 192 |
| Bortezomib | 115 |
| Zoledronic Acid | 108 |
| Daratumumab | 106 |
| Carfilzomib | 41 |
| Denosumab | 24 |
| Intravenous Immunoglobulin | 22 |
| Elotuzumab | 19 |
| Rituximab | 17 |
| Cyclophosphamide | 16 |
| Obinutuzumab | 16 |
| Acetaminophen | 7 |
| Palonosetron | 6 |
| Diphenhydramine | 5 |
| Pamidronate Disodium | 5 |
| Bendamustine | 4 |
| Diphenhydramine | 2 |
| Fludarabine Phosphate | 2 |
| Ondansetron | 2 |
| Aciclovir | 2 |
| Allopurinol | 1 |
| Aspirin | 1 |
| Trimethoprim/sulfamethoxazole | 1 |
| Cisplatin | 1 |
| Cyanocobalamin | 1 |
| Doxorubicin | 1 |
| Epoetin Alfa-Epbx | 1 |
| Etoposide | 1 |
| Melphalan Hydrochloride | 1 |
| Methylpredisolone Acetate | 1 |

Note: Patients can be administered more than 1 treatment, so the sum of the frequencies will not equal the total sample size.

**eTable 3.** Socioeconomics and Treatment Characteristics by Patient Report of Financial Difficulty by Measure and by Diagnosis.

| **Characteristics** | **Reported Financial Difficulty**<br>**(Single-item Measure)** |  |  | **Reported Financial Difficulty**<br>**(Composite Measure)** |  |  |
| --- | --- | --- | --- | --- | --- | --- |
|  | **MM**<br>**No. (%)** | **CLL**<br>**No. (%)** | **Total**<br>**No. (%)** | **MM**<br>**No. (%)** | **CLL**<br>**No. (%)** | **Total**<br>**No. (%)** |
| No. of Patients | 62 | 8 | 70 | 188 | 63 | 251 |
| Age in years, Mean (SD) | 63.6 (10.84) | 69.8 (12.21) | 64.3 (11.09) | 65.8 (10.03) | 70.6 (8.06) | 67.0 (9.78) |
| Sex |  |  |  |  |  |  |
| Female | 34 (55) | 0 (0) | 34 (49) | 95 (51) | 20 (32) | 115 (46) |
| Male | 28 (45) | 8 (100) | 36 (51) | 93 (49) | 43 (68) | 136 (54) |
| Race |  |  |  |  |  |  |
| White | 49 (79) | 6 (75) | 55 (79) | 161 (86) | 58 (92) | 219 (87) |
| Non-White<sup>a</sup> | 13 (21) | 2 (25) | 15 (21) | 27 (14) | 5 (8) | 32 (13) |
| Geographic Region<sup>b</sup> |  |  |  |  |  |  |
| Northeast | 4 (7) | 1 (13) | 5 (7) | 13 (7) | 3 (5) | 16 (6) |
| Midwest | 37 (60) | 5 (63) | 42 (60) | 116 (62) | 37 (59) | 153 (61) |
| South | 18 (29) | 1 (13) | 19 (27) | 52 (28) | 19 (30) | 71 (28) |
| West | 3 (5) | 1 (13) | 4 (6) | 7 (4) | 4 (6) | 11 (4) |
| Education |  |  |  |  |  |  |
| High School Diploma/GED or below | 40 (66) | 4 (50) | 44 (64) | 89 (48) | 36 (57) | 125 (50) |
| Above High School Diploma | 21 (34) | 4 (50) | 25 (36) | 96 (52) | 27 (43) | 123 (50) |
| Missing | 1 | 0 | 1 | 3 | 0 | 3 |
| Home Ownership Status |  |  |  |  |  |  |
| Homeowner | 46 (75) | 6 (75) | 52 (75) | 151 (81) | 56 (89) | 207 (83) |
| Non-Homeowner | 15 (25) | 2 (25) | 17 (25) | 36 (19) | 7 (11) | 43 (17) |
| Missing | 1 | 0 | 1 | 1 | 0 | 1 |
| Reported Household Income |  |  |  |  |  |  |
| Less than \$20,000 | 18 (32) | 1 (13) | 19 (29) | 37 (21) | 6 (10) | 43 (19) |
| \$20,000 to \$39,999 | 14 (25) | 4 (50) | 18 (28) | 39 (23) | 16 (28) | 55 (24) |
| \$40,000 to \$59,999 | 13 (23) | 3 (38) | 16 (25) | 35 (20) | 16 (28) | 51 (22) |
| \$60,000 to \$79,999 | 6 (11) | 0 (0) | 6 (9) | 20 (12) | 6 (10) | 26 (11) |
| \$80,000 to \$99,999 | 4 (7) | 0 (0) | 4 (6) | 16 (9) | 2 (3) | 18 (8) |
| \$100,000 or more | 2 (4) | 0 (0) | 2 (3) | 26 (15) | 12 (21) | 38 (17) |
| Missing | 5 | 0 | 5 | 15 | 5 | 20 |
| Insurance Type |  |  |  |  |  |  |
| Medicare only | 5 (8) | 2 (25) | 7 (10) | 15 (8) | 4 (6) | 19 (8) |
| Medicare + other insurance | 31 (50) | 3 (38) | 34 (49) | 110 (59) | 47 (75) | 157 (63) |

|  | Reported Financial Difficulty<br>(Single-item Measure) |  |  | Reported Financial Difficulty<br>(Composite Measure) |  |  |
| --- | --- | --- | --- | --- | --- | --- |
|  | MM<br>No. (%) | CLL<br>No. (%) | Total<br>No. (%) | MM<br>No. (%) | CLL<br>No. (%) | Total<br>No. (%) |
| Other<sup>c</sup> | 26 (42) | 3 (38) | 29 (41) | 63 (34) | 12 (19) | 75 (30) |
| Charlson Comorbidity Index score |  |  |  |  |  |  |
| 1 or more | 25 (40) | 4 (50) | 29 (41) | 71 (38) | 25 (40) | 96 (38) |
| 0 | 37 (60) | 4 (50) | 41 (59) | 117 (62) | 38 (60) | 155 (62) |
| Treatment Received |  |  |  |  |  |  |
| Clinician-Administered Therapeutic | 52 (84) | 2 (25) | 54 (77) | 158 (84) | 21 (33) | 179 (71) |
| Not a Clinician-Administered<br>Therapeutic | 10 (16) | 6 (75) | 16 (23) | 30 (16) | 42 (67) | 72 (29) |

Abbreviations: MM: Multiple Myeloma, CLL: Chronic Lymphocytic Leukemia.
<sup>a</sup> Non-White race captures patients who selected American Indian or Alaska Native, Asian, Black, or African American, Native Hawaiian, or other Pacific Islander, Unknown, Not Reported
<sup>b</sup> Geographic Regions were divided according to the United States census divisions and regions: **Northeast:** Maine, New Hampshire, Vermont, Massachusetts, Rhode Island, Connecticut, New York, New Jersey, and Pennsylvania; **Midwest:** Ohio, Michigan, Indiana, Wisconsin, Illinois, Minnesota, Iowa, Missouri, North Dakota, South Dakota, Nebraska, and Kansas; **South:** Delaware, Maryland, Virginia, West Virginia, Kentucky, North Carolina, South Carolina, Tennessee, Georgia, Florida, Alabama, Mississippi, Arkansas, Louisiana, Texas, and Oklahoma, Washington DC; **West:** Montana, Idaho, Wyoming, Colorado, New Mexico, Arizona, Utah, Nevada, California, Oregon, Washington, Alaska, and Hawaii.
<sup>c</sup> Other includes: Private Insurance, Medicaid, Military Sponsored (including CHAMPUS & TRICARE), Veterans Sponsored, Not Otherwise Specified (NOS), and any combination of those listed here.

**eTable 4.** Sensitivity Analysis Single Measure of Financial Difficulty Using Different Dichotomizations.

| **Model** | **Unadjusted Odds Ratios (95% CI)** | **P value** | **Adjusted Odds Ratios (95% CI)<sup>a</sup>** | **P value** |
| --- | --- | --- | --- | --- |
| **Single-item Measure of Financial Difficulty – Not at all/A little/Quite a bit vs. Very much** |  |  |  |  |
| Female | 2.47 (0.90, 6.81) | .07 | 2.00 (0.70, 5.68) | .20 |
| Non-White | 1.11 (0.25, 5.03) | .89 | 0.82 (0.17, 3.96) | .81 |
| Comorbidity | 0.74 (0.26, 2.15) | .58 | 0.78 (0.26, 2.38) | .67 |
| CLL | OR unable to be calculated – zero cell | X | OR unable to be calculated – zero cell | X |
| Clinician-Administered Treatment | 2.29 (0.65, 8.11) | .19 | 0.79 (0.21, 2.96) | .72 |
| Above HS Education | 0.34 (0.12, 0.97) | .04 | 0.31 (0.11, 0.93) | .04 |
| Medicare + Other<sup>b</sup> | NA [Reference] | NA | NA [Reference] | NA |
| Medicare | 1.19 (0.15, 9.74) | .99 | 0.97 (0.11, 8.52) | .88 |
| Other<sup>c</sup> | 1.46 (0.52, 4.10) | .67 | 1.32 (0.44, 3.93) | .69 |
| **Single-item Measure of Financial Difficulty – Not at all/A little vs. Quite a bit/Very much** |  |  |  |  |
| Female | 1.32 (0.78, 2.21) | .30 | 1.09 (0.62, 1.91) | .77 |
| Non-White | 2.97 (1.49, 5.90) | .002 | 2.21 (1.04, 4.72) | .04 |
| Comorbidity | 1.39 (0.82, 2.36) | .27 | 1.53 (0.86, 2.75) | .15 |
| CLL | 0.31 (0.15, 0.68) | .003 | 0.34 (0.13, 0.84) | .02 |
| Clinician-Administered Treatment | 1.72 (0.94, 3.15) | .08 | 1.02 (0.49, 2.12) | .96 |
| Above HS Education | 0.42 (0.24, 0.71) | .001 | 0.36 (0.21, 0.64) | .001 |
| Medicare + Other<sup>b</sup> | NA [Reference] | NA | NA [Reference] | NA |
| Medicare | 3.17 (1.21, 8.27) | .02 | 2.17 (0.77, 6.07) | .14 |
| Other<sup>c</sup> | 2.50 (1.43, 4.37) | .001 | 2.53 (1.37, 4.66) | .003 |
| **Single-item Measure of Financial Difficulty – Not at all vs. A little/Quite a bit/Very much** |  |  |  |  |
| Female | 0.82 (0.56, 1.21) | .32 | 0.87 (0.58, 1.32) | .51 |
| Non-White | 0.63 (0.33, 1.19) | .15 | 0.85 (0.43, 1.67) | .63 |
| Comorbidity | 0.82 (0.55, 1.23) | .34 | 0.74 (0.48, 1.14) | .17 |
| CLL | 2.15 (1.36, 3.39) | .001 | 1.83 (1.06, 3.17) | .03 |
| Clinician-Administered Treatment | 0.62 (0.41, 0.95) | .03 | 0.86 (0.52, 1.44) | .58 |
| Above HS Education | 1.40 (0.95, 2.07) | .09 | 1.52 (1.01, 1.13) | .05 |
| Medicare + Other<sup>b</sup> | NA [Reference] | NA | NA [Reference] | NA |
| Medicare | 0.38 (0.16, 0.92) | .24 | 0.45 (0.18, 1.13) | .45 |
| Other<sup>c</sup> | 0.42 (0.26, 0.65) | .19 | 0.41 (0.25, 0.65) | .11 |

**eTable 5.**
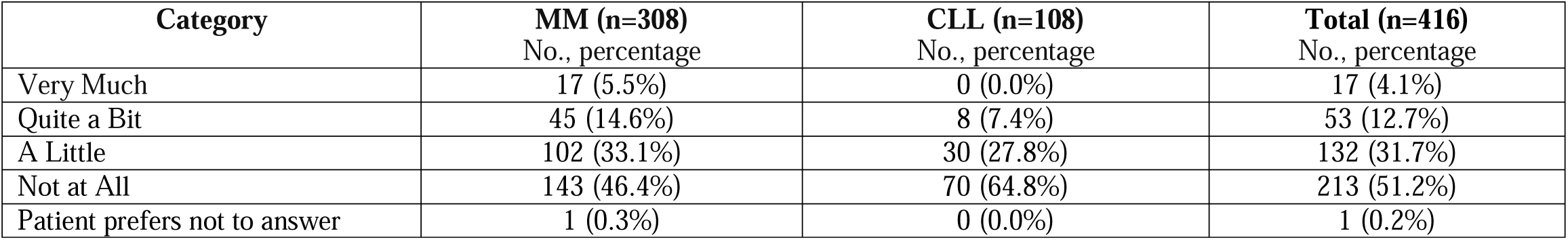
Endorsement of Categories for Single Measure of Financial Difficulty by Diagnosis.

**eTable 6.** Endorsement of Items in Composite Measure of Financial Difficulty by Diagnosis.

| **Survey Question** | **MM (n=308)**<br>**No., proportion**<br>**(95% Wilson CI)** | **CLL (n=108)**<br>**No., proportion**<br>**(95% Wilson CI)** | **P value** |
| --- | --- | --- | --- |
| (EORTC QLQ-C30) item #28: “Has your physical condition or medical treatment caused you financial difficulties in the past year? (Part 1, Question 8) | 62<br>0.201<br>(0.160, 0.250) | 8<br>0.074<br>(0.038, 0.140) | .002 |
| In the past 12 months, did you have problems paying or were unable to pay any medical bills? Include bills for doctors, hospitals, therapists, medication, equipment, nursing home or home. (Part 1, Question 1) | 72<br>0.234<br>(0.190, 0.284) | 16<br>0.148<br>(0.093, 0.227) | .06 |
| Do you or anyone in your family currently have medical bills that you are unable to pay at all? (Part 1, Question 2) | 40<br>0.130<br>(0.097, 0.172) | 7<br>0.065<br>(0.032, 0.128) | .07 |
| During the past 12 months, have you or someone in your family delayed medical care because you were worried about the cost (do not include dental care)? (Part1, Question 4) | 48<br>0.156<br>(0.120, 0.201) | 8<br>0.074<br>(0.038, 0.139) | .03 |
| During the past 12 months, was there a time when you or someone in your family needed medical care but didn’t get it because you couldn’t afford it? (Part 1, Question 5) | 27<br>0.088<br>(0.061, 0.125) | 2<br>0.019<br>(0.005, 0.065) | .02 |
| During the past 12 months, was there a time when you needed one of the following, but did not get it because you couldn’t afford it? **Prescription medicine** (Part 1, Question 6a) | 30<br>0.097<br>(0.069, 0.136) | 11<br>0.102<br>(0.058, 0.173) | .89 |
| During the past 12 months, was there a time when you needed one of the following, but did not get it because you couldn’t afford it? **Mental health care or counseling** (Part 1, Question 6b) | 9<br>0.029<br>(0.015, 0.055) | 3<br>0.028<br>(0.010, 0.079) | .94 |
| During the past 12 months, was there a time when you needed one of the following, but did not get it because you couldn’t afford it? **Dental care** (Part 1, Question 6c) | 62<br>0.201<br>(0.160, 0.250) | 17<br>0.157<br>(0.101, 0.238) | .32 |
| During the past 12 months, was there a time when you needed one of the following, but did not get it because you couldn’t afford it? **Eyeglasses** (Part 1, Question 6d) | 37<br>0.120<br>(0.088, 0.161) | 9<br>0.083<br>(0.045, 0.151) | .29 |
| During the past 12 months, was there a time when you needed one of the following, but did not get it because you couldn’t afford it? **Cancer-related medical care** (Part 1, Question 6e) | 17<br>0.055<br>(0.035, 0.087) | 4<br>0.037<br>(0.015, 0.091) | .46 |
| During the past 12 months, was there a time when you needed one of the following, but did not get it because you couldn’t afford it? **Non-cancer related medical care** (Part 1, Question 6f) | 26<br>0.084<br>(0.058, 0.121) | 5<br>0.046<br>(0.020, 0.104) | .19 |
| If you get sicker or have an accident, how worried are you that you will not be able to pay your medical bills? (Part 1, Question 9) | 60<br>0.195 | 6<br>0.056 | <.001 |

| Survey Question | MM (n=308)<br>No., proportion<br>(95% Wilson CI) | CLL (n=108)<br>No., proportion<br>(95% Wilson CI) | P value |
| --- | --- | --- | --- |
|  | (0.154, 0.243) | (0.026, 0.116) |  |
| How often in the last 12 months would you say you were worried or stressed about having enough money to pay your rent or mortgage? (Part 1, Question 10) | 68<br>0.221<br>(0.178, 0.270) | 14<br>0.130<br>(0.079, 0.206) | .04 |
| How often in the last 12 months would you say you were worried or stressed about having enough money to buy nutritious meals? (Part 1, Question 11) | 57<br>0.185<br>(0.146, 0.232) | 9<br>0.083<br>(0.045, 0.151) | .01 |
| How often in the last 12 months would you say you were worried or stressed about having enough money to pay household utilities such as water, gas, and electricity? (Part 1, Question 12) | 60<br>0.195<br>(0.154, 0.243) | 14/107<br>0.131<br>(0.080, 0.208) | .14 |
| During the past 12 months, were any of the following true for you: **You skipped medication doses to save money** (Part 1, Question 16a) | 22<br>0.071<br>(0.048, 0.106) | 4<br>0.037<br>(0.015, 0.091) | .20 |
| During the past 12 months, were any of the following true for you: **You took less medicine to save money** (Part 1, Question 16b) | 25<br>0.081<br>(0.056, 0.117) | 4/107<br>0.037<br>(0.015, 0.092) | .13 |
| During the past 12 months, were any of the following true for you: **You delayed filling a prescription to save money** (Part 1, Question 16c) | 32<br>0.104<br>(0.075, 0.143) | 14/107<br>0.131<br>(0.080, 0.208) | .44 |
| During the past 12 months, were any of the following true for you: You used alternative therapies to save money (Part 1, Question 16f) | 14<br>0.046<br>(0.027, 0.075) | 4/107<br>0.037<br>(0.015, 0.092) | .72 |
| In the past 12 months, have you or has anyone in your family had to borrow money or go into debt because of your cancer, its treatment, or the lasting effects of that treatment? (Part 1, Question 17) | 39<br>0.127<br>(0.094, 0.168) | 8<br>0.074<br>(0.038, 0.139) | .14 |
| In the past 12 months, did you or your family file for bankruptcy because of your cancer, its treatment, or the lasting effects of that treatment? (Part 1, Question 18) | 3<br>0.010<br>(0.003, 0.028) | 0/107<br>0.000 | .31 |
| In the past 12 months, have you ever worried about having to pay large medical bills related to your cancer? (Part 1, Question 20) | 134/307<br>0.437<br>(0.381, 0.492) | 39<br>0.361<br>(0.277, 0.455) | .17 |
| Please think about medical care visits for cancer, its treatment, or the lasting effects of that treatment in the past 12 months. Have you ever been unable to cover your share of those visits? (Part 1, Question 21) | 31/307<br>0.101<br>(0.072, 0.140) | 9<br>0.083<br>(0.045, 0.151) | .59 |

Note: The proportion of patients who answered yes or always/usually/sometimes to the question in the table above are presented descriptively by disease status. Each cell presents the following information: (number of patients who answered yes or always/usually/sometimes)/(total number of patients who answered the question), the proportion, and the 95% Wilson score confidence interval.

## References

1. Cook R. Economic and clinical impact of multiple myeloma to managed care. Journal of managed care pharmacy. 2008;14(7 Supp A):19-25.

2. Chen Q, Jain N, Ayer T, et al. Economic burden of chronic lymphocytic leukemia in the era of oral targeted therapies in the United States. Journal of Clinical Oncology. 2017;35(2):166.

3. Bhattacharya K, Bentley JP, Ramachandran S, et al. Phase-Specific and Lifetime Costs of Multiple Myeloma Among Older Adults in the US. JAMA Network Open. 2021;4(7):e2116357–e2116357.

4. Goyal RK, Nagar SP, Kabadi SM, Le H, Davis KL, Kaye JA. Overall survival, adverse events, and economic burden in patients with chronic lymphocytic leukemia receiving systemic therapy: Real-world evidence from the medicare population. Cancer Medicine. 2021;10(8):2690–2702. doi:10.1002/cam4.3855

5. Ubel PA. Full Disclosure — Out-of-Pocket Costs as Side Effects. New England Journal of Medicine. 2013;369(16):1484–1486. doi:10.1056/NEJMp1306826

6. Zullig LL, Peppercorn JM, Schrag D, et al. Financial distress, use of cost-coping strategies, and adherence to prescription medication among patients with cancer. Journal of oncology practice. 2013;9(6S):60s–63s.

7. Stump TK, Eghan N, Egleston BL, et al. Cost concerns of patients with cancer. Journal of oncology practice. 2013;9(5):251–257.

8. Kaiser Family Foundation. USA Today/Kaiser Family Foundation/Harvard School of Public Health National Survey of Households Affected by Cancer. Kaiser Family Foundation; 2006. Accessed November 5, 2021. https://www.kff.org/health-costs/poll-finding/usa-todaykaiser-family-foundationharvard-school-of-public-2/

9. Felder TM, Bennett CL. Can patients afford to be adherent to expensive oral cancer drugs?: unintended consequences of pharmaceutical development. Journal of oncology practice. 2013;9(6S):64s–66s.

10. Shankaran V, Ramsey S. Addressing the financial burden of cancer treatment: from copay to can’t pay. JAMA oncology. 2015;1(3):273–274.

11. PDQ® Adult Treatment Editorial Board. Financial Toxicity (Financial Distress) and Cancer Treatment (PDQ®). National Cancer Institute. Published September 2019. Accessed November 5, 2021. https://www.cancer.gov/about-cancer/managing-care/track-care-costs/financial-toxicity-pdq

12. Huntington SF, Weiss BM, Vogl DT, et al. Financial toxicity in insured patients with multiple myeloma: a cross-sectional pilot study. The Lancet Haematology. 2015;2(10):e408–e416.

13. Yabroff KR, Zhao J, Han X, Zheng Z. Prevalence and correlates of medical financial hardship in the USA. Journal of general internal medicine. 2019;34(8):1494–1502.

14. Knight TG, Deal AM, Dusetzina SB, et al. Financial toxicity in adults with cancer: adverse outcomes and noncompliance. Journal of oncology practice. 2018;14(11):e665–e673.

15. Zafar SY, McNeil RB, Thomas CM, Lathan CS, Ayanian JZ, Provenzale D. Population-based assessment of cancer survivors’ financial burden and quality of life: a prospective cohort study. Journal of Oncology Practice. 2015;11(2):145–150. doi:10.1200/JOP.2014.001542

16. Shankaran V, Unger JM, Darke AK, et al. S1417CD: a prospective multicenter cooperative group-led study of financial hardship in metastatic colorectal cancer patients. JNCI. 2022;114(3):372–380. 10.1093/jnci/djab210

17. Abrams HR, Durbin S, Huang CX, et al. Financial toxicity in cancer care: origins, impact, and solutions. Translational Behavioral Medicine. 2021;11(11):2043–2054. doi:10.1093/tbm/ibab091

18. Yabroff KR, Dowling EC, Guy Jr GP, et al. Financial hardship associated with cancer in the United States: findings from a population-based sample of adult cancer survivors. Journal of clinical oncology. 2016;34(3):259.

19. Chino F, Peppercorn JM, Rushing C, et al. Out-of-pocket costs, financial distress, and underinsurance in cancer care. JAMA oncology. 2017;3(11):1582–1584.

20. Zhao J, Han X, Zheng Z, Banegas MP, Ekwueme DU, Yabroff KR. Is health insurance literacy associated with financial hardship among cancer survivors? Findings from a national sample in the United States. JNCI Cancer Spectrum. 2019;3(4):pkz061. 10.1093/jncics/pkz061

21. Zahnd WE, Davis MM, Rotter JS, et al. Rural-urban differences in financial burden among cancer survivors: an analysis of a nationally representative survey. Supportive Care in Cancer. 2019;27(12):4779–4786. 10.1007/s00520-019-04742-z

22. Lopes L, Kearney A, Montero A, Hamel L, Brodie M. Health Care Debt In The U.S.: The Broad Consequences Of Medical And Dental Bills. KFF. Published June 16, 2022. Accessed June 16, 2022. https://www.kff.org/report-section/kff-health-care-debt-survey-main-findings/

23. Doshi JA, Li P, Huo H, Pettit AR, Armstrong KA. Association of patient out-of-pocket costs with prescription abandonment and delay in fills of novel oral anticancer agents. Journal of Clinical Oncology. 2018;36(5):476–482.

24. Lee MJ, Khan MM, Salloum RG. Recent trends in cost-related medication nonadherence among cancer survivors in the United States. Journal of managed care & specialty pharmacy. 2018;24(1):56–64.

25. Bestvina CM, Zullig LL, Rushing C, et al. Patient-oncologist cost communication, financial distress, and medication adherence. Journal of Oncology Practice. 2014;10(3):162–167.

26. Dusetzina SB, Huskamp HA, Rothman RL, et al. Many Medicare beneficiaries do not fill high-price specialty drug prescriptions. Health Affairs. 2022;41(4):487–496. 10.1377/hlthaff.2021.01742

27. Yabroff KR, Bradley C, Shih YCT. Understanding financial hardship among cancer survivors in the United States: strategies for prevention and mitigation. Journal of Clinical Oncology. 2020;38(4):292–301. doi:10.1200/JCO.19.01564

28. Einav L, Jenkins M, Levin J. The impact of credit scoring on consumer lending. The RAND Journal of Economics. 2013;44(2):249–274.

29. Chetty R, Looney A, Kroft K. Salience and taxation: Theory and evidence. American economic review. 2009;99(4):1145–1177.

30. Liebman JB, Luttmer EF. The perception of Social Security incentives for labor supply and retirement: The median voter knows more than you’d think. Tax Policy and the Economy. 2012;26(1):1–42.

31. Howard DH, Bach PB, Berndt ER, Conti RM. Pricing in the Market for Anticancer Drugs. Journal of Economic Perspectives. 2015;29(1):139–162.

32. Leukemia & Lymphoma Society. *Updated Data on Blood Cancers: Facts 2020-*2021. Leukemia & Lymphoma Society; 2021. https://www.lls.org/sites/default/files/2021-08/PS80%20FactsBook_2020_2021_FINAL.pdf

33. Fonseca R, Hagiwara M, Panjabi S, Yucel E, Buchanan J, Delea T. Economic Burden of Disease Progression Among Transplant Eligible Multiple Myeloma Patients Who Have Received at Least One Line of Therapy in the US. Blood. 2019;134(Supplement_1):2211. doi:10.1182/blood-2019-123743

34. Snyder S, Albertson T, Garcia J, Gitlin M, Jun MP. Travel-Related Economic Burden of Chimeric Antigen Receptor T Cell Therapy Administration by Site of Care. Adv Ther. 2021;38(8):4541–4555. doi:10.1007/s12325-021-01839-y

35. The NCI Community Oncology Research Program (NCORP). About NCORP. The NCI Community Oncology Research Program (NCORP). Accessed December 14, 2022. https://ncorp.cancer.gov/

36. Stull DE, Leidy NK, Parasuraman B, Chassany O. Optimal recall periods for patient-reported outcomes: challenges and potential solutions. Current medical research and opinion. 2009;25(4):929–942.

37. EORTC Quality of Life. EORTC Quality of Life Website - EORTC - Quality of Life : EORTC – Quality of Life. EORTC Quality of Life. Accessed December 1, 2021. https://qol.eortc.org/, https://qol.eortc.org/

38. Aaronson NK, Ahmedzai S, Bergman B, et al. The European Organization for Research and Treatment of Cancer QLQ-C30: a quality-of-life instrument for use in international clinical trials in oncology. JNCI: Journal of the National Cancer Institute. 1993;85(5):365–376.

39. Hjermstad MJ, Fossa SD, Bjordal K, Kaasa S. Test/retest study of the European Organization for Research and Treatment of Cancer Core Quality-of-Life Questionnaire. Journal of Clinical Oncology. 1995;13(5):1249–1254.

40. Cattell RB. The scree test for the number of factors. Multivariate behavioral research. 1966;1(2):245–276.

41. Shih YCT, Smieliauskas F, Geynisman DM, Kelly RJ, Smith TJ. Trends in the cost and use of targeted cancer therapies for the privately insured nonelderly: 2001 to 2011. Journal of Clinical Oncology. 2015;33(19):2190.

42. Hollmann S, Moldaver D, Goyert N, Grima D, Maiese EM. A U.S. Cost Analysis of Triplet Regimens for Patients with Previously Treated Multiple Myeloma. J Manag Care Spec Pharm. 2019;25(4):449–459. doi:10.18553/jmcp.2019.25.4.449

43. Altice CK, Banegas MP, Tucker-Seeley RD, Yabroff KR. Financial hardships experienced by cancer survivors: a systematic review. JNCI: Journal of the National Cancer Institute. 2017;109(2).

44. Carrera PM, Kantarjian HM, Blinder VS. The financial burden and distress of patients with cancer: understanding and stepping-up action on the financial toxicity of cancer treatment. CA: A Cancer Journal for Clinicians. 2018;68(2):153–165. doi:10.3322/caac.21443

45. Witte J, Mehlis K, Surmann B, et al. Methods for measuring financial toxicity after cancer diagnosis and treatment: a systematic review and its implications. Annals of Oncology. 2019;30(7):1061–1070.

46. Petermann V, Zahnd WE, Vanderpool RC, et al. How cancer programs identify and address the financial burdens of rural cancer patients. Supportive care in cancer. 2022;30(3):2047–2058.

47. Kircher SM, Yarber J, Rutsohn J, et al. Piloting a financial counseling intervention for patients with cancer receiving chemotherapy. Journal of oncology practice. 2019;15(3):e202–e210.

48. Corrigan KL, Fu S, Chen YS, et al. Financial toxicity impact on younger versus older adults with cancer in the setting of care delivery. Cancer. 2022;128(13):2455–2462. doi:10.1002/cncr.34220

49. de Moor JS, Mollica M, Sampson A, et al. Delivery of Financial Navigation Services Within National Cancer Institute–Designated Cancer Centers. JNCI Cancer Spectrum. 2021;5(3):pkab033.

50. McLouth LE, Nightingale CL, Dressler EV, et al. Current practices for screening and addressing financial hardship within the NCI Community Oncology Research Program. Cancer Epidemiology, Biomarkers & Prevention. 2021;30(4):669–675.

