## Supplementary material for "Self-Reported Financial Difficulties Among Patients with Multiple Myeloma and Chronic Lymphocytic Leukemia Treated at U.S. Community Oncology Clinics (Alliance A231602CD)": Study Protocol

Activation Amendment

**ALLIANCE FOR CLINICAL TRIALS IN ONCOLOGY**

**PROTOCOL UPDATE TO ALLIANCE A231602CD**

**Assessing Financial Difficulty in Patients with Blood Cancers**

| **X** | **Update:** |  | **Status Change:** |
| --- | --- | --- | --- |
| **X** | Eligibility changes |  | Activation |
| **X** | Therapy / Dose Modifications / Study Calendar changes |  | Closure |
| **X** | Informed Consent changes |  | Suspension / temporary closure |
|  | Scientific / Statistical Considerations changes |  | Reactivation |
| **X** | Data Submission / Forms changes |  |  |
| **X** | Editorial / Administrative changes |  |  |
|  | Other : |  |  |

**Expedited review is allowed. IRB approval (or disapproval) is required within 90 days. Please follow your IRB of record guidelines.**

**UPDATES:**

**Cover page:**

- The study chair Dr.Rena Conti’s institutional affiliation and contact information have been updated.
- Dr. Robert Behrens has been moved up on the cover page to reflect that he is the Alliance co- chair.
- The institutional affiliation and contact information have been updated for Stacie Dusetzina, the Health Economics co-chair of the study.
- Bruce Rapkin’s contact information has been updated.
- Niveditha Subbiah has replaced John Taylor as the protocol coordinator, and her contact information has been updated on the cover page.

**Schema page: Patient eligibility** **(p.4)**

The first row of the schema has been revised as follows, with the underlined text added: “Patients must have current diagnosis of chronic lymphocytic leukemia (CLL) and/or multiple myeloma (MM)”, to clarify that patients who have both diagnoses are eligible as well. This change has been made to Section 3.2.1, and throughout the protocol where there are references to CLL and/or multiple myeloma.

- In the first bullet for Site Eligibility criteria, “Completion of A231602CD….” has been corrected to read “Intent to complete the A231602 …” , since sites should have intent to complete the survey when they consider participating in the study. This change is also reflected in Section

3.3.1 of the protocol.

- The term “site of care” has been replaced on the schema and throughout the protocol with the term “practice” to clarify the level of site data collection. The term “practice” has been defined in the introduction paragraph to the practice survey in Appendix I.
- The first column of the schema diagram has been revised to state “See Section 10.1” within parenthesis replacing “ IRB approval” (below “Site Activation”). Section 10.1 describes the process for site enrollment and the 8 week timeline for sites to complete the Practice Survey will begin from this date. The reference to Section 10.1 has also been added to Section 5.2 (second column, first row) and to Section 7.2 (first paragraph, last sentence).

**Table of Contents** **(pp.6-7)**

The table of contents has been updated.

**Section 1.0 Background** **(p.8)**

“Practice” has been added, following the term “site”, in the first sentence of the first paragraph of this section.

**Section 2.2 Secondary objectives** **(p.12)**

The number of secondary objectives has been corrected from nine to ten.

**Section 3.3 Site Eligibility Criteria** **(p.14)**

Section 3.3.3 has been revised as follows: “ Sites seeking to enroll Spanish- speaking patients must have Spanish speaking staff on site or through the use of a translation service to be able to conduct the informed consent discussion in Spanish”, in order to clarify that sites are allowed to use translation services for conducting the informed consent discussion.

**Section 4.5 Descriptive variables** **(p.18)**

In the fourth bullet of this section, the question asked on the practice survey within quotes, has been removed and replaced with the question currently asked on the revised survey.

**Section 7.2 Practice Survey****(p.21)**

- A new second paragraph has been added, to describe the process for submission of Spanish language practice surveys for practices completing the survey in Puerto Rico.
- The first sentence of the third paragraph has been updated to reflect that information regarding the domains listed in Appendix I will be collected on the practice survey. In the second sentence of this paragraph, “to one hour” has been added following 45 minutes, as the practice survey may take upto one hour to complete, based on pilot testing. This is also reflected in Section 10.2, and included in the update summary for that section.

**Section 7.4 Patient Registration and Medical Record Abstraction** **(p.22)**

- In the first sentence of the second paragraph, country of origin has been replaced with country of residence, to correctly reference the patient level information that is being collected.
- The first sentence of the fourth paragraph of this section has been revised to read as follows: “ We expect the patient survey to take ~~30-45~~ 60 minutes to complete based on study team work….”. This information has been updated in the last paragraph of Section 10.2 and in the “What will happen…” and “What exams, tests and procedures…” sections of the model consent form as well.
- In the last sentence of the last paragraph of this section, the reference to “Appendices II and III” has been corrected to reference “Appendix II”.

**Section 10.1** **Site credentialing/****Eligibility** **requirements (p. 35)**

- Two new paragraphs have been added as the second and third paragraphs of this section, to provide instructions to practices for site (practice) enrollment in the study and to enable site registration in Medidata Rave.

**Section 10.2 Site Selection and Site Recruitment** **(p.35)**

- In the middle of the third sentence of the first paragraph of this Section, “…one page summary” has been replaced with “two-page summary” to reflect the length of the current site recruitment letter.
- In the last sentence of the third paragraph of this section, “30-45 minutes” has been replaced with “45 minutes”.

**Section 12: Model Consent form** **(pp.40-46)**

- In the “ What am I being asked to do..” section, a new second sentence has been added to the first paragraph, to incorporate the most recent NCI informed consent template language. The second paragraph has been revised as follows: “We are asking you to take part in this research study because you have either chronic lymphocytic leukemia ~~or~~and multiple myeloma, or both.” The rationale for this change is provided in the Patient eligibility: Schema section of the Update summary.
- The estimated time taken for completing the telephone survey has been updated from 30-45 minutes to one hour in the third sentences of the “What will happen..” and the “What exams, tests and procedures…” sections of the consent form, based on pilot testing of the survey.
- In the “Who will see my medical information..” section of the consent form, the first sentence of the fourth paragraph has been revised as follows: “There are organizations that may look at or receive copies of some of the information in your study records.” In the bulleted list following the fourth paragraph, the second bullet has been updated to include the NCI Central IRB (to replace the IRB). The two paragraphs following the bulleted list have been updated to include the required language from the most recent NCI informed consent template.
- The note to investigators above the signature portion has been removed to comply with the instructions received with the new NCI informed consent template.

**Appendix I: Practice survey (English and Spanish)** **(pp. 47-81)**

The overview table of the practice survey has been updated with the revised domain titles and number of questions. Based on pilot-testing feedback, the following changes have been made to the previous “Site of Care” Survey:

- The entire introduction paragraph has been reformatted and replaced with new text which introduces the survey to be completed by sites and defines the term “practice”. The term “site” has been replaced with the term “practice” throughout the survey and the protocol. A new second paragraph has been added following the introduction paragraph to provide guidance for CTEP- IDs covering multiple physical practice locations.
- In Section 1 of the survey, contact information, “Organization” has been removed below Job title. NCORP Name has been added as the first field, and Practice CTEP-ID and Practice name associated with CTEP ID have been added as the last two fields.
- In Section 2, the second conditional question following question 1 (c) has been removed. Question 5 has also been removed. Subsequent questions have been renumbered.
- Section 3. Clinical Practice Characteristics:
  - Question #5, three of the options starting with “Physicians” have been revised to clarify what exactly is being asked, and moved below “All nurse practitioners...”.
  - A statement has been added before Question #6 to provide an introduction to the next set of questions. The response options have been enhanced to better represent the different ways practices may access services.
  - “Patient portal” along with yes/no checkboxes has been added as Option (d) for question 9.
  - “Opportunities to volunteer for research studies or patient survey”, along with yes or no checkboxes has been added as option (e) for question 10.
- Section 5. Cancer Care Delivery Systems:
  - Two questions on patient communication around medication issues and contractual pharmacy relationships have been added as questions 14 and 16. The title before question 17 has been revised to read “**Patient Reported Health and Financial** ~~and Health~~ **Literacy.”**
- Section 6. Patient-centered financial navigation, transportation and psychosocial support services:
  - The previous title for this section has been revised as follows: “Patient-centered financial navigation, transportation and psychosocial support, ~~transportation~~ services and ~~financial~~ ~~navigation~~.”
  - The introduction statement following the title has been reformatted and a definition for “Financial navigation” has been included below the introduction. The topics have been reorganized in the following order: financial navigation, transportation and psychosocial services.
  - Response options for financial navigation, transportation and psychosocial services have been included.
  - For all three services (financial navigation, transportation and psychosocial services), questions have been added regarding challenges practices may experience in providing these services and the potential for reducing any current services.
  - A final conclusion statement has been added to thank practices for completing the survey.
- The Spanish translated version of the practice survey has been inserted following the English version.

**Appendix II: Patient Survey English and Spanish** **(pp.82-160)**

Based on pilot-testing feedback and formatting the survey for forms development, the following changes have been made to the Patient Survey:

- Text has been added describing the response options for each question (e.g. select one, yes/no for each). Questions inserted in tables have been removed and placed in the body of the document (e.g. Part 1, Questions 6, 7, 22-36). The skip logic has been updated as needed.
  - Starting Phone Interview with Participant: The text in this section has been removed and replaced with new text to make it more conversational.
- Part 1: Financial Difficulty:
  - Questions regarding patient medication have been moved from Section 3 to Section 1.
- Part 2: Patient Report of Seeking and Receiving Financial Support:
  - The note to reviewers has been removed as this is no longer applicable.
  - Under “Formal Support for Specific Purposes,” the introduction paragraph has been revised and the questions have been restructured to improve the question flow and descriptions and examples have been added as needed.
- Part 3: Patient Sociodemographic Indicators:
  - Under “Patient Insurance” the response options have been updated to capture a greater range of insurance types.
- Closing Question:
  - An open text closing question has been added asking respondents if there is anything else they would like to share with us.
  - The closing interview has been replaced with text to include more resources for the patients after survey completion.
- The Spanish version of the survey and introduction script has been translated to match the current version of the English patient survey, and the previous version has been removed. The closing interview has been translated and included as well.

Apart from the above revisions to the protocol, the following documents are being submitted for review as separate attachments:

1) Revised Site recruitment letter (English and Spanish versions).
2) Patient thank you letter to be sent with gift card (English and Spanish versions).
3) Certificates of translation into Spanish for patient and practice surveys, site recruitment letter and patient thank you letter.

**A replacement protocol document has been issued**

**Attach to the front of every copy of this protocol**

ALLIANCE FOR CLINICAL TRIALS IN ONCOLOGY ALLIANCE A231602CD

**Assessing Financial Difficulty in Patients with Blood Cancers**

Study Chair

Rena M. Conti, PhD

Boston University Questrom School of Business

Boston, MA

Community Oncology Study Co-chair

Robert Behrens, MD

Iowa Wide Oncology Research Coalition

| Health Outcomes Co-Chair | CCDR Co-chair |
| --- | --- |
| Antonia Bennett, PhD | Bruce Rapkin, PhD |
| University of North Carolina | Montefiore Medical Center |
| Tel: 919-962-5427 | Tel: 718-839-7453 |
| | |

| Health Economics Co-Chair | Multiple Myeloma Co-Chair | CCDR Committee Chair |
| --- | --- | --- |
| Stacie Dusetzina, PhD | Paul Richardson, MD | George J. Chang, MD, MS |
| Vanderbilt University | Dana Farber Cancer Institute | MDACC |
| Tel: 615-875-9281 | Tel: 617-632-2127 | Tel: 713-563-1875 |
| | | |

| Primary Statistician | Secondary Statistician |
| --- | --- |
| Jeff Sloan, PhD | Travis Dockter |
| Tel:507-284-9985 Fax: 507-  266-2477 | Tel: 507-266-9803 Fax: 507-  266-2477 |
| | |

| Data Manager | Protocol Coordinator |
| --- | --- |
| Cristina Zabel | Niveditha Subbiah ,LLM , MA |
| Tel: 507-284-4565 | Tel: 773-702-9934 Fax: 312-345-0117 |
| | |

Participants:

NCORP components of the Alliance (lead), ECOG-ACRIN, NRG, and SWOG NCORP Research Bases

*Version Date: 2/5/2019*

**Study Resources:**

| **Expedited Adverse Event Reporting**  <http://eapps-ctep.nci.nih.gov/ctepaers/> | **Medidata Rave®** **iMedidata portal**  https://login.imedidata.com |
| --- | --- |
| **OPEN (Oncology Patient Enrollment Network)**  https://open.ctsu.org | **Biospecimen Management System**  [http://bioms.allianceforclinicaltrialsinoncology.org](http://bioms.allianceforclinicaltrialsinoncology.org/) |
| **Protocol Contacts:** | |
| **A231602CD Nursing Contact** | |
| Mary Beth Wilwerding Missouri Valley Cancer Consortium  Tel: 402-991-8070  | |

| **Protocol-related questions may be directed as follows:** | |
| --- | --- |
| **Questions:** | **Contact (via email):** |
| Questions regarding patient eligibility: | Study Chair, Nursing Contact, Protocol Coordinator, and (where applicable) Data  Manager |
| Questions related to data submission, RAVE or patient follow-up: | Data Manager |
| Questions regarding the protocol document and  model informed consent: | Protocol Coordinator |
| Questions related to IRB review | Alliance Regulatory Inbox |
| Questions related to patient interview | UNC research staff (study specific) |

**CANCER TRIALS SUPPORT UNIT (CTSU) ADDRESS AND CONTACT INFORMATION**

| **For regulatory requirements:** | **For patient enrollments:** | **For study data submission:** |
| --- | --- | --- |
| Regulatory documentation must submitted to the CTSU via the Regulatory Submission Portal:  Regulatory Submission Portal (Sign in at [www.ctsu.org,](http://www.ctsu.org/) and select the Regulatory Submission sub- tab under the Regulatory tab.)  Institutions with patients waiting that are unable to use the Portal should alert the CTSU Regulatory Office immediately at 1-866-651- 2878 to receive further instruction and support.  Contact the CTSU Regulatory Help Desk at 1- 866-651-2878 for regulatory assistance. | Please refer to the patient enrollment section for instructions on using the Oncology Patient Enrollment Network (OPEN) which can be accessed at https:/[/w](http://www.ctsu.org/OPEN_)w[w.ctsu.org/OPEN_](http://www.ctsu.org/OPEN_) SYSTEM/ or https://OPEN.ctsu.org.  Contact the CTSU Help Desk with any OPEN-related questions at | Data collection for this study will be done exclusively through Medidata Rave. Please see the data submission section of the protocol for further instructions. |
| The **study protocol and all related forms and documents** must be downloaded from the protocol- specific page of the CTSU Member website located at https:/[/w](http://www.ctsu.org/)w[w.ctsu.org.](http://www.ctsu.org/) Access to the CTSU members’ website is managed through the Cancer Therapy and Evaluation Program - Identity and Access Management (CTEP-IAM) registration system and requires user log on with CTEP-IAM username and password. Permission to view and download this protocol and its supporting documents is restricted and is based on person and site roster assignment housed in the CTSU RSS. | | |
| **For clinical questions (i.e., patient eligibility or treatment-related)** see the Protocol Contacts, Page 2*.* | | |
| **For non-clinical questions (i.e., unrelated to patient eligibility, treatment, or clinical data submission)** contact the CTSU Help Desk by phone or e-mail:  CTSU General Information Line – 1-888-823-5923, or All calls and correspondence will be triaged to the appropriate CTSU representative. | | |
| **The CTSU website is located at** https[://w](http://www.ctsu.org/)ww.[ctsu.org.](http://www.ctsu.org/) | | |

**ASSESSING FINANCIAL DIFFICULTY IN PATIENTS WITH BLOOD CANCERS**

**Patient Eligibility**

| Patients must have current diagnosis of chronic lymphocytic leukemia (CLL) and/or multiple myeloma (MM) | **Required Initial Laboratory Values** |
| --- | --- |
| Patients’ medical records must be available to the registering institution. | None |
| Eligible patients must have been prescribed drug-based anticancer therapy, whether administered orally or by infusion, within the prior 12 months. Specifically, eligible patients are those who: | |

- Are presently being treated with infused or orally-administered anticancer therapy, OR
- Completed infused or orally-administered anti-cancer therapy in the past 12 months, OR
- Were prescribed infused or orally-administered anticancer therapy within the prior 12 months yet chose to forego treatment

Not currently enrolled in a clinical trial in which drug is supplied by the study.

Patients with psychiatric illness or other mental impairment that would preclude their ability to give informed consent or to respond to the telephone survey are not eligible.

Patients must be able to read and comprehend English or Spanish. Age ≥ 18 years

**Patient level Schema**

| **Patient consent and registration** |  | 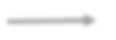 Site staff will complete medical chart abstraction within 1 week of registration. |  | Patients complete the survey via telephone interview with UNC study team survey staff within 8 weeks following registration. 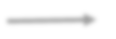 |
| --- | --- | --- | --- | --- |

**Please refer to the full protocol text for a complete description of the eligibility criteria and intervention plan.**

**Site Eligibility**

- Intent to complete the A231602CD Practice Survey (Appendix I)
- Access to patient medical records: Registering institution must have access to patient medical records if recruiting patients at any other clinic (as medical abstraction is required for collecting study data).
- Sites must be able to conduct informed consent discussion in Spanish, if necessary.

**Site level Schema**

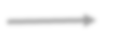

A one-time practice survey of the component that is recruiting patients onto the study will be completed within 8 weeks after site activation

**Site Activation (See** **Section 10.1****)**

**Please refer to the full protocol text for a complete description of the site eligibility criteria and study implementation**

Table of Contents

Section	Page

1.0 BACKGROUND	8
1.1 Significance and rationale	8
1.2 Theoretical Development	8
1.3 Contribution to Cancer Care Delivery	9
1.4 Rationale for Study Design	10
1.5 Study Population	10
1.6 Impact on Subsequent Research	11
1.7 Impact on Patient Care and Patient Outcomes	11
1.8 Justification of Endpoints	11
1.9 Anticipated participation and accrual from NCORP sites and other members	11
2.0 OBJECTIVES	12
2.1 Primary objective	12
2.2 Secondary objectives	12
3.0 PATIENT SELECTION	14
3.1 On-Study Guidelines	14
3.2 Patient Eligibility Criteria	14
3.3 Site Eligibility Criteria	14
4.0 PATIENT REGISTRATION	15
4.1 CTEP/ DCP Investigator Registration Procedures	15
4.2 CTSU Site Registration Procedures	16
4.3 Patient Registration Requirements	17
4.4 Patient Registration/Randomization Procedures	18
4.5 Descriptive Variables	18
5.0 STUDY CALENDAR	19
5.1 Patient-level study calendar	19
5.2 Site-level study calendar	19
6.0 DATA SUBMISSION	20
6.1 Data Collection and Submission	20
7.0 STUDY IMPLEMENTATION	21
7.1 Identification of participating institutions	21
7.2 Practice survey	21
7.3 Patient recruitment and enrollment	21
7.4 Patient Registration and Medical Record Abstraction	22
7.5 Patient Survey	22
8.0 MEASURES	23
8.1 Definition of Primary and Key Secondary Endpoints	23
8.2 Patient-reported measures	25
9.0 STATISTICAL CONSIDERATIONS	28
9.1 Study Design	28

9.2 Primary objective: To estimate the proportion of patients with MM and/or CLL who report experiencing financial difficulty in the past 12 months	29
9.3 Plan for missing data and other statistical considerations	30
9.4 Secondary objectives	30
9.5 Monitoring	33
9.6 Reporting	34
9.7 Inclusion of Women and Minorities	34
10.0 GENERAL REGULATORY CONSIDERATIONS AND CREDENTIALING	35
10.1 Site Credentialing/Eligibility requirements	35
10.2 Site Selection and Site Recruitment	35
10.3 Waivers of patient consent	36

11.0 REFERENCES	37
12.0 MODEL INFORMED CONSENT FORM	40

APPENDIX I	PRACTICE SURVEY (ENGLISH AND SPANISH)	47

APPENDIX II	PATIENT SURVEY (ENGLISH AND SPANISH	82

1.0 **BACKGROUND**

Please note: Throughout the document we refer to NCORP “site”, “practice” or “component” to describe a clinic, cancer center, physician practice, or other institution where patients with cancer are enrolled in a CCDR study.

Also, throughout the document we use the following terms defined below: “Financial worry” is the anticipation of a problem of financial difficulty.

“Financial difficulty” is the objective financial problem, or the patients’ reported experience of financial burdens and their ramifications.

“Financial burden” describe the objective economic indicators of financial demands or experience of financial difficulty that can lead to debt, bankruptcy or foregoing treatment (cancer.gov).

### 1.1 Significance and rationale

Treatment for cancer is undergoing a renaissance, yet financial difficulty encountered over the course of cancer diagnosis and treatment is a growing concern among patients, families, physicians and national provider groups.^1-7^ Recent reports have demonstrated that patients with cancer may be at risk of self-reported treatment-related financial difficulty and financial burden. Reports of financial burden include an inability to pay for basic necessities such as food and utility bills as a result of cancer treatment, the presence of medical debt, and high out of pocket burdens relative to income among other measures. Stakeholders are searching for answers to mitigate financial burden and difficulty among patients with cancer including the design of financial navigation tools. Others have suggested physicians and medical practices assume a larger role in navigating patients through the financial responsibilities associated with cancer treatment.^8^

The empirical literature on the prevalence of financial difficulty among cancer patients is in an early stage of development. In addition, the relationship between financial difficulty and patient socioeconomic circumstances, disease and treatment characteristics is poorly understood. We also know very little about the extent to which patients receive or apply for charitable aid and from what sources, or about practice engagement with activities that aim to alleviate patient financial difficulty.

### 1.2 Theoretical Development

In 2015-2016, the study chair and collaborators performed a review of published studies of patient-reported financial difficulty and burden associated with medical care using a PUBMED search augmented by a hand search of published studies’ citations. They then narrowed the search to those studies focused on patients with cancer. The resulting meta-analysis is available online at the National Cancer Institute’s Physician Data Query (PDQ) website.^9^ These efforts identified significant gaps in the published literature on cancer patients’ treatment-related financial difficulty and consequently informed our study’s objectives, hypotheses and methods.

To provide a conceptual basis for our study, Grossman and Becker propose theoretical models of individual demand for medical care, based on an idiosyncratic shock to health (cancer diagnosis) and how it may be influenced by a patient’s socioeconomic resources (e.g. household assets, income, educational attainment).^10-13^ Patients with a cancer diagnosis face a decision to be treated based on an expectation of benefit and their expected “price” for treatment that they will be required to pay. Self-reported financial difficulty in a given year can occur because a patient elects to be treated, but exceeds their budget constraint due to inadequate information and lack of preparation, liquidity constraints^14^ or salience.^15,16^ A full economic model is provided here.

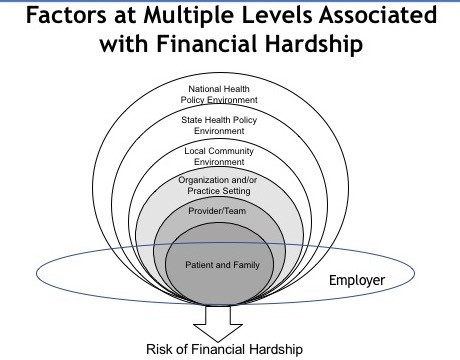

Yabroff, 2018

This model effectively illustrates the range of factors, including policy, practice, provider and patient characteristics that interact to shape experiences of financial difficulty. These factors are directly related to all of our study concepts and objectives: financial difficulty (2.2.1); financial navigation (2.2.2 and 2.2.3); financial burden (2.2.4 and 2.2.5); financial support (2.2.6 and 2.2.7); patient concerns regarding treatment and costs of care (2.2.8 and 2.2.9) and patient health and well-being (2.2.10).

Based upon this simple model we posit that patient self-reported financial difficulty associated with a cancer diagnosis and treatment is likely related to their household’s income and assets; presence and generosity of insurance coverage, disease characteristics, receipt of charity care to underwrite financial and non-financial costs of treatment and practice engagement with identifying and ameliorating patient financial difficulty.

We have chosen to focus on patients with multiple myeloma (MM) and chronic lymphocytic leukemia (CLL) because these cancers have important medical care utilization and quality of life implications for affected patients and their families.^17^ Furthermore, new orally-administered anticancer treatments for these blood cancers have been introduced into the U.S. medical market in recent years, promising gains in both survival and quality of life, but with high prices and significant out of pocket costs for many patients. These treatments are used in addition to other medical care including frequent diagnostic monitoring, hospitalizations to address adverse events associated with diagnosis and treatment, and, if necessary, allogeneic transplantation and associated care. The coordination and costs of these activities may deplete patients’ and their families’ financial resources, interfere with ability to work and make it difficult to afford other necessities. No national study has systematically assessed these endpoints in this patient population.

### 1.3 Contribution to Cancer Care Delivery

This study will measure the prevalence of patient-reported financial difficulty, specific financial burdens and resources currently available to patients and from practices to assist with patient

financial navigation. We will examine associations among patient-reported financial difficulty and patient socioeconomic and disease characteristics. We will also examine associations among patient-reported financial difficulty and practice resources. Data collection for this study will entail two comprehensive and theoretically grounded financial assessment surveys: one at the patient-level and one at the NCI (National Cancer Institute) Community Oncology Research Program (NCORP) practice level. The practice-level survey will gather specific information regarding current practice organization and current and planned resources dedicated to cancer patient financial navigation. Previous completion of the NCORP CCDR Landscape Assessment is not a requirement for practice participation in this study. Results of this study will directly inform efforts to improve financial navigation and resources for MM and CLL patients. We expect potentially fruitful interventions to be directed at both the patient and the practice levels. These may include improved patient-level articulation of financial worry and specific financial and non-financial preferences for assistance, practice-level comprehensive financial counseling, and advanced financial planning to reduce delays in the initiation or continuity of treatment. Measures of patient differences in current concerns and health-related quality of life will make it possible to target and tailor financial interventions according to personal priorities and circumstances. Beyond MM and CLL, patient and practice surveys introduced in this study will aid development of interventions to reduce financial difficulties associated with cancer diagnosis and treatment in general. These surveys may be adopted into routine clinical care and alongside cooperative group trials to understand how financial factors influence patients’ treatment decisions, including enrollment on trials.

### 1.4 Rationale for Study Design

The study team has designed a conceptually based, hypothesis driven, observational, cross- sectional study, entailing the collection of survey data from patients and NCORP sites/practices. We have used data from the 2015 and 2016 NCORP CCDR Landscape Assessment, a meta- analysis of previously published literature on cancer patient financial difficulty, and our own qualitative pilot interviews with patients, physicians, minority and community cancer clinic administrators, and various other stakeholders and study advisors to inform the study design. In addition, four NCORP sites are active participants in study pilot work and concept and protocol development.

Surveying a national sample of patients and sites is the only systematic way to measure specific aspects of patient financial difficulty, since such experience cannot be systematically nor reliably inferred by a review of patient medical charts, electronic medical records or claims data. For example, many types of cost assistance offered to patients experiencing financial distress by patient advocacy groups, pharmaceutical companies, physician organizations and local charities are also not recorded in these sources. Furthermore, our study directly complements existing knowledge regarding NCORP practice resources derived from previously completed Landscape Assessments.

### 1.5 Study Population

Patients treated in NCORP sites are the ideal sampling frame for conducting this study. NCORP site staff will recruit patients based on study eligibility criteria and a limited medical record review. NCORP provides a common point of access to patients being treated in a wide variety of community oncology settings. The national scope of NCORP makes it feasible to sample participants receiving care in diverse geographic regions, encompassing background differences in costs of living and available insurance programs. Recruitment will occur over a 24-month period. We focus on individuals with a current diagnosis of chronic lymphocytic leukemia (CLL) and/or multiple myeloma (MM) (both incident and prevalent cases) whose current or recent treatment course includes pharmaceutical based care and who are not currently enrolled

in a treatment-based trial (registry trial is allowed). The practice survey will be completed at the site level that reflects recruited patients’ site of cancer care.

### 1.6 Impact on Subsequent Research

Our study will measure the prevalence of patient reported financial difficulty and the association between financial difficulty and patient socioeconomic status, disease characteristics and practice resources dedicated to patient financial navigation. The survey instruments can be subsequently adapted for use in other cancer types, time periods, as secondary endpoints in cooperative group trials, and in planned interventions.

### 1.7 Impact on Patient Care and Patient Outcomes

We aim to identify modifiable drivers of financial difficulty among patients with cancer and design feasible and effective alternative interventions at the patient and practice levels.

### 1.8 Justification of Endpoints

The primary objective of this study is to estimate the proportion of individuals with MM and/or CLL treated at NCORP practices who report experiencing financial difficulty in the past 12 months. The secondary objectives are: 1) To describe the association of patient report of financial difficulty with insurance status; and 2) To describe current resources dedicated to patient financial navigation among participating NCORP sites. 3) To describe the association of patient report of financial difficulty with receiving treatment at practices that report offering patients financial guidance through navigators or social workers, and controlling for patient socioeconomic status; 4) To identify distinct patterns of financial burden among patients undergoing treatment for MM and/or CLL; 5) To examine the relationship between distinct patterns of financial burden with patient report of financial difficulty, patient socio- demographics, and patient disease characteristics; 6) To estimate the proportion of patients with MM and/or CLL undergoing treatment who report receiving financial support in the past 12 months; 7) To describe the association of patient report of receiving financial support with receiving treatment at practices offering patients financial guidance through navigators or social workers, and with socioeconomic status; 8) To describe the magnitude of patient concerns regarding treatment and costs of care; 9) To describe the association of patient concerns regarding treatment and costs of care with patient socio-demographics, disease and practice characteristics; and 10) To describe the association of financial difficulty with patients self- reported health and well-being ; and 11) To describe the types of psychosocial, transportation and financial navigation interventions practices are developing.

### 1.9 Anticipated participation and accrual from NCORP sites and other members.

A sample size of 500 patients, assuming reasonable accrual rates, will provide substantial precision for our estimates. We will collaborate NCORP CCDR site leads to identify sites to recruit into the study given feasibility concerns and recruitment goals. In addition to fulfilling an NCORP requirement to engage in CCDR studies, we expect that most sites will agree to participate because study results will be of intrinsic interest. Site staff will not be responsible for conducting interviews with patients. One-time patient surveys will be conducted as a telephone interview by a centralized team of survey implementation experts. The one-time survey of practice resources and services to address patients’ financial needs will be completed by site staff via Medidata Rave, maximizing convenience and minimizing need for coordination and scheduling.

This target recruitment of patients is feasible in NCORP sites. Counts of incident MM and CLL patient cases per eligible site were drawn from the 2015 NCORP CCDR Landscape Assessment (only n=27 out of 46 sites reported data). Based on these statistics, we expect to identify

approximately 2,700 incident MM (1,400) and CLL (1,300) patients who are potentially eligible for study participation. Unfortunately, the Landscape Assessment did not collect prevalent cases of cancer. Since we are interested in surveying both incident and prevalent cases, we reviewed the Surveillance, Epidemiology, and End Results (SEER) data. According to SEER, the ratio of 5- year prevalent to incident cases of these cancers is approximately 4:1.^19^ Consequently, we expect there will be approximately 13,500 incident and prevalent cases of these cancers (2,700+4(2,700)) treated by the 27 sites reporting data in a given year. With a conservative assumption that only 25% of these patients will be eligible for study participation, we expect an upper bound of 3,375 patients eligible for study.

Once sites have been recruited to the study, we will work with each practice to determine the optimal combination of staff to initiate patient recruitment. We have engaged with NCORP investigators at primarily minority-serving sites to increase the likelihood of their participation. We will evaluate recruitment targets with attention paid to the inclusion of adequate representation across minority and socioeconomically disadvantaged groups.

2.0 **OBJECTIVES**

### 2.1 Primary objective

To estimate the proportion of patients with MM and/or CLL who report experiencing financial difficulty in the past 12 months.

### 2.2 Secondary objectives

Our ten secondary objectives address five aspects of patient financial experience of cancer treatment:

*Financial difficulty*

2.2.1 To describe the association of patient report of financial difficulty with insurance status.

*Financial navigation*

2.2.2 To describe the association of patient report of financial difficulty with receiving treatment at practices that report offering patients financial guidance through navigators or social workers, and controlling for patient socioeconomic status.
2.2.3 To describe the types of psychosocial, transportation and financial navigation interventions practices are developing.

*Financial burden*

2.2.4 To identify distinct patterns of financial burden among patients undergoing treatment for MM and/ or CLL.
2.2.5 To examine the relationship between distinct patterns of financial burden with patient report of financial difficulty, patient socio-demographics, and patient disease characteristics.

*Financial support*

2.2.6 To estimate the proportion of patients with MM and/or CLL undergoing treatment who report receiving financial support in the past 12 months.
2.2.7 To describe the association of patient report of receiving financial support with receiving treatment at practices offering patients financial guidance through navigators or social workers, and with socioeconomic status.

*Patient concerns regarding treatment and costs of care*

2.2.8 To describe the magnitude of patient concerns regarding treatment and costs of care.
2.2.9 To describe the association of patient concerns regarding treatment and costs of care with patient socio-demographics, disease and practice characteristics.

*Patient health and well-being*

2.2.10 To describe the association of financial difficulty with patient self-reported health and well- being.
3.0 **PATIENT SELECTION**

For questions regarding eligibility criteria, see the Study Resources page. Please note that the Study Chair cannot grant waivers to eligibility requirements.

### 3.1 On-Study Guidelines

This clinical trial can fulfill its objectives only if patients appropriate for this trial are enrolled. All relevant medical and other considerations should be taken into account when deciding whether this protocol is appropriate for a particular patient.

### 3.2 Patient Eligibility Criteria

**3.2.1 Documentation of disease:** Patients must have current diagnosis of chronic lymphocytic leukemia (CLL) and/or multiple myeloma (MM).

**3.2.2 Patients’ medical records must be available to the registering institution.**

**3.2.3 Eligible patients must have been prescribed drug-based anticancer therapy, whether administered orally or by infusion, within the prior 12 months.** Specifically, eligible patients are those who:

- Are presently being treated with infused or orally-administered anticancer therapy, OR
- Completed infused or orally-administered anti-cancer therapy in the past 12 months, OR
- Were prescribed infused or orally-administered anticancer therapy within the prior 12 months yet chose to forego treatment.

**3.2.4 Not currently enrolled in a clinical trial in which drug is supplied by the study.**

**3.2.5 Patients with psychiatric illness or other mental impairment that would preclude their ability to give informed consent or to respond to the telephone survey are not eligible.**

**3.2.6 Patients must be able to read and comprehend English or Spanish.**

**3.2.7 Age ≥ 18 years**

### 3.3 Site Eligibility Criteria

**3.3.1 Intent to complete the A231602CD Practice Survey** (Appendix I).

**3.3.2 Access to patient medical records:** Registering institution must have access to patient medical records, either on site or via request from other institutions, if recruiting patients at a site **(as medical abstraction is required for collecting study data)**.

**3.3.3 Sites seeking to enroll Spanish- speaking patients must have Spanish speaking staff on site or through the use of a translation service to be able to conduct the informed consent discussion in Spanish.**

4.0 **PATIENT REGISTRATION**

### 4.1 CTEP/ DCP Investigator Registration Procedures

Food and Drug Administration (FDA) regulations and National Cancer Institute (NCI) policy require all individuals contributing to NCI-sponsored trials to register and to renew their registration annually. To register, all individuals must obtain a Cancer Therapy Evaluation Program (CTEP) Identity and Access Management (IAM) account (https://ctepcore.nci.nih.gov/iam). In addition, persons with a registration type of Investigator (IVR), Non-Physician Investigator (NPIVR), or Associate Plus (AP) (i.e., clinical site staff requiring write access to OPEN, RAVE, or TRIAD or acting as a primary site contact) must complete their annual registration using CTEP’s web-based Registration and Credential Repository (RCR) (<https://ctepcore.nci.nih.gov/rcr>). Documentation requirements per registration type are outlined in the table below.

| **Documentation Required** | **IVR** | **NPIVR** | **AP** | **A** |
| --- | --- | --- | --- | --- |
| FDA Form 1572 | ✔ | ✔ |  |  |
| Financial Disclosure Form | ✔ | ✔ | ✔ |  |
| NCI Biosketch (education, training, employment, license, and certification) | ✔ | ✔ | ✔ |  |
| HSP/GCP training | ✔ | ✔ | ✔ |  |
| Agent Shipment Form (if applicable) | ✔ |  |  |  |
| CV (optional) | ✔ | ✔ | ✔ |  |

An active CTEP-IAM user account and appropriate RCR registration is required to access all CTEP and CTSU (Cancer Trials Support Unit) websites and applications. In addition, IVRs and NPIVRs must list all clinical practice sites and IRBs covering their practice sites on the FDA Form 1572 in RCR to allow the following:

- Added to a site roster
- Assigned the treating, credit, consenting, or drug shipment (IVR only) tasks in OPEN
- Act as the site-protocol PI on the IRB approval

Additional information can be found on the CTEP website at < https://ctep.cancer.gov/investigatorResources/default.htm >. For questions, please contact the RCR Help Desk by email at <>.

Registration requires the submission of:

Human Subject Protection (HSP) training certificate

### 4.2 CTSU Site Registration Procedures

This study is supported by the NCI Cancer Trials Support Unit (CTSU).

**IRB Approval:**

Each investigator or group of investigators at a clinical site must obtain IRB approval for this protocol and submit IRB approval and supporting documentation to the CTSU Regulatory Office before they can be approved to enroll patients.

Assignment of site registration status in the CTSU Regulatory Support System (RSS) uses extensive data to make a determination of whether a site has fulfilled all regulatory criteria including but not limited to:

- an active Federal Wide Assurance (FWA) number,
- an active roster affiliation with the Lead Network or a participating organization,
- a valid IRB approval, and
- compliance with all protocol specific requirements.

In addition, the site-protocol Principal Investigator (PI) must meet the following criteria:

- Active registration status
- The IRB number of the site IRB of record listed on their Form FDA 1572
- An active status on a participating roster at the registering site.

Sites participating on the NCI CIRB initiative and accepting CIRB approval for the study are not required to submit separate IRB approval documentation to the CTSU Regulatory Office. For sites using the CIRB, IRB approval information is received from the CIRB and applied to the RSS in an automated process. Signatory Institutions must submit a Study Specific Worksheet for Local Context (SSW) to the CIRB via IRBManager to indicate their intention to open the study locally. The CIRB’s approval of the SSW is then communicated to the CTSU Regulatory Office. In order for the SSW approval to be processed, the Signatory Institution must inform the CTSU which CIRB-approved institutions aligned with the Signatory Institution are participating in a given study.

4.2.1 **Downloading Site Registration Documents**

Site registration forms may be downloaded from the A231602CD protocol page located on the CTSU members’ website. Go to https:[//w](http://www.ctsu.org/)ww[.ctsu.org](http://www.ctsu.org/) and log in to the members’ area using your CTEP-IAM username and password

- Click on the Protocols tab in the upper left of your screen
- Either enter the protocol # in the search field at the top of the protocol tree, or
- Click on the By Lead Organization folder to expand
- Click on the Alliance link to expand, then select trial protocol # A231602CD
- Click on LPO Documents, select the Site Registration documents link, and download and complete the forms provided.
4.2.2 **Requirements for A231602CD Site Registration**
      - **IRB approval** (For sites not participating via the NCI CIRB; local IRB documentation, an IRB-signed CTSU IRB Certification Form, Protocol of Human Subjects Assurance

Identification/IRB Certification/Declaration of Exemption Form, or combination is accepted)

- **Institutional Approval Document:** Institutions that are interested in participating in this study must first contact the Study Chair, Rena Conti, to review study requirements and procedures and to identify who will be responsible for the study at their respective institution. See Section 10.0.
4.2.3 **Submitting Regulatory Documents**

Submit required forms and documents to the CTSU Regulatory Office via the Regulatory Submission Portal, where they will be entered and tracked in the CTSU RSS.

Regulatory Submission Portal: [www.ctsu.org](http://www.ctsu.org/) (members’ area) 🡪 Regulatory Tab 🡪

Regulatory Submission

When applicable, original documents should be mailed to: CTSU Regulatory Office

1818 Market Street, Suite 3000

Philadelphia, PA 19103

Institutions with patients waiting that are unable to use the Portal should alert the CTSU Regulatory Office immediately at 1-866-651-2878 in order to receive further instruction and support.

4.2.4 **Checking Your Site’s Registration Status**

You can verify your site registration status on the members’ section of the CTSU website.

- Go to https:[//w](http://www.ctsu.org/)ww[.ctsu.org](http://www.ctsu.org/) and log in to the members’ area using your CTEP-IAM username and password
- Click on the Regulatory tab
- Click on the Site Registration tab
- Enter your 5-character CTEP Institution Code and click on Go

Note: The status given only reflects compliance with IRB documentation and institutional compliance with protocol-specific requirements outlined by the Lead Network. It does not reflect compliance with protocol requirements for individuals participating on the protocol or the enrolling investigator’s status with the NCI or their affiliated networks.

### 4.3 Patient Registration Requirements

- **Informed consent:** The patient must willingly consent after being informed of the study procedure to be followed, the nature of the study, alternatives, potential benefits, side effects, risks, and discomforts. Current human subjects protection committee approval of this protocol and a consent form is required prior to patient consent and registration.
- **Protected health information:** In order to be able to contact participants who agree to participate in the survey study, it will be necessary to collect those participants’ names, and telephone numbers In addition, following the telephone survey conducted by UNC study team survey staff, the patient’s mailing addresses will be requested in order to send him/her a gift card. This information will be destroyed upon completion of the study. None of this PHI will be stored with the study data.

### 4.4 Patient Registration/Randomization Procedures

Patient enrollment will be facilitated using the Oncology Patient Enrollment Network (OPEN). OPEN is a web-based registration system available on a 24/7 basis. To access OPEN, the site user must have an active CTEP-IAM account (check at https://ctepcore.nci.nih.gov/iam) and a 'Registrar' role on either the LPO or participating organization roster. Registrars must hold a minimum of an AP registration type.

All site staff will use OPEN to enroll patients to this study. It is integrated with the CTSU Enterprise System for regulatory and roster data and, upon enrollment, initializes the patient in the Rave database. OPEN can be accessed at https://open.ctsu.org or from the OPEN tab on the CTSU members’ side of the website at https[://w](http://www.ctsu.org/)ww.[ctsu.org.](http://www.ctsu.org/) To assign an IVR or NPIVR as the treating, crediting, consenting, drug shipment (IVR only), or investigator receiving a transfer in OPEN, the IVR or NPIVR must list on their Form FDA 1572 in RCR the IRB number used on the site’s IRB approval.

Prior to accessing OPEN, site staff should verify the following:

- All eligibility criteria have been met within the protocol stated timeframes.
- All patients have signed an appropriate consent form and HIPAA authorization form (if applicable).

Note: The OPEN system will provide the site with a printable confirmation of registration and treatment information. Please print this confirmation for your records.

Further instructional information is provided on the OPEN tab of the CTSU members’ side of the CTSU website at https[://w](http://www.ctsu.org/)ww.[ctsu.org](http://www.ctsu.org/) or at https://open.ctsu.org. For any additional questions contact the CTSU Help Desk at 1-888-823-5923 or

### 4.5 Descriptive Variables

Descriptive factors for each study objective are described in the statistical analysis plan. Briefly:

- Latent classes of socioeconomic status will be identified as described in the statistical analysis plan (see secondary objective #2).
- Patient-reported health, quality of life, priorities and demographic variables.
- Patient disease characteristics are obtained from the medical chart abstraction. This includes date of MM or CLL diagnosis, pre-existing comorbidities and MM- and/or CCL-related comorbidities, and MM and/ or CLL-specific treatments and treatment initiation dates.
- Whether a practice offers financial guidance will be obtained via the practice survey question Does your practice screen patients for their interest in receiving help with financial navigation for their medical care? (Yes/No). This question is based on the 2015 NCORP CCDR Landscape Assessment question (#24).
  5.0 **STUDY CALENDAR**

Laboratory and clinical parameters during treatment are to be followed using individual institutional guidelines and the best clinical judgment of the responsible physician. It is expected that patients on this study will be cared for by physicians experienced in the treatment and supportive care of patients on this trial.

### 5.1 Patient-level study calendar

|  | **Within 1 week after patient registration** | **Within 8 weeks after patient registration** |
| --- | --- | --- |
| Medical record abstraction | X (1) | X (2) |
| Patient survey (Appendices II and III) |  |  |

1 Site staff will abstract records for the 12-month period prior to registration. See Section

7.4 for a summary of the information to be extracted from the medical record.

2 To be conducted as a centralized telephone interview by study team survey staff at UNC. See Section 7.5.

### 5.2 Site-level study calendar

|  | **After site activation (See** **Section 10.1****)** | **Within 8 weeks after activation** |
| --- | --- | --- |
| Medical record review (for patient eligibility/enrollment) | X | X |
| Practice Survey (Appendix I) |  |  |

6.0 **DATA SUBMISSION**

### 6.1 Data Collection and Submission

**Data collection by site staff** for this study will be done exclusively through the Medidata Rave clinical data management system. Access to the trial in Rave is granted through the iMedidata application to all persons with the appropriate roles assigned in Regulatory Support System (RSS). To access Rave via iMedidata, the site user must have an active CTEP-IAM account (check at https://ctepcore.nci.nih.gov/iam) and the appropriate Rave role (Rave CRA, Read- Only, Site Investigator) on either the LPO or participating organization roster at the enrolling site. To hold the Rave CRA role or CRA Lab Admin role, the user must hold a minimum of an AP registration type. To hold the Rave Site Investigator role, the individual must be registered as an NPIVR or IVR. Associates can hold read-only roles in Rave.

Upon initial site registration approval for the study in RSS, all persons with Rave roles assigned on the appropriate roster will be sent a study invitation e-mail from iMedidata. To accept the invitation, site users must log into the Select Login (https://login.imedidata.com/selectlogin) using their CTEP-IAM user name and password, and click on the “accept” link in the upper right-corner of the iMedidata page. Please note, site users will not be able to access the study in Rave until all required Medidata and study specific trainings are completed. Trainings will be in the form of electronic learnings (eLearnings), and can be accessed by clicking on the link in the upper right pane of the iMedidata screen.

Users who have not previously activated their iMedidata/Rave account at the time of initial site registration approval for the study in RSS will also receive a separate invitation from iMedidata to activate their account. Account activation instructions are located on the CTSU website, Rave tab under the Rave resource materials (Medidata Account Activation and Study Invitation Acceptance). Additional information on iMedidata/Rave is available on the CTSU members’ website under the Rave tab at [www.ctsu.org/RAVE/](http://www.ctsu.org/RAVE/) or by contacting the CTSU Help Desk at 1-888-823-5923 or by e-mail at

A Schedule of Forms is available on the Alliance study webpage, within the Case Report Forms section.

**Data collection by UNC research staff:**

The patient survey will be conducted via telephone interview by the research staff at UNC and data collected through the interview will be entered into Rave. See Section 7.5 below.

7.0 **STUDY IMPLEMENTATION**

### 7.1 Identification of participating institutions

Interested sites should contact the study chair, Dr. Conti to discuss the criteria listed in Section 10.1.

A guiding principle in designing the study is minimization of staff and patient time commitments. Based on our pilot testing, we expect each recruited site staff commitments to entail the following: 1-5 hours for identification of all eligible patients; 10-15 minutes for each patient consent and 30-45 minutes for each patient registration, including medical record abstraction.

### 7.2 Practice survey

After local site activation, appropriate site staff who can answer questions regarding clinic characteristics, patient characteristics, and patient-centered services should be identified to complete the survey. It is acceptable if more than one person at each site is tasked with completing the survey; based, for example, on their familiarity with some aspects of the site’s characteristics or practices and not others. These designated personnel will complete, the practice survey in Medidata Rave within eight weeks after site activation (See Section 10.1**)**. Site staff should complete this survey prior to the registration of their first patient.

In order to accommodate language needs, the submission process will be modified for the Puerto Rico NCORP. If the Puerto Rico NCORP and any of their affiliated practices agree to participate in the study, they will be provided with a paper-based Spanish-language version of the Survey. Upon completion of the Survey, designated personnel will be instructed to upload the Survey in Medidata Rave and a Spanish-speaking study team member will enter the Survey responses into the English-language form, preserving all free-text responses in their original language. After the data is exported and as part of the data analysis, any Spanish-language free-text will be translated into English for analysis.

The practice survey will collect information regarding the domains listed in Appendix I. We expect 45 minutes to one hour will be required for practice survey completion. Each practice that intends to register patients is required to complete this one-time survey.

### 7.3 Patient recruitment and enrollment

Participating sites will identify eligible patients at the practice-level through a brief medical record review or other sources. Given that patient eligibility is dependent on a medical record review, sites will only be allowed to recruit patients from the clinic/site described in the practice survey. For example, sites may run queries of their own sources such as electronic medical records or billing systems to identify eligible patients.

Sites will follow institutional policies in requesting a “waiver of authorization” from their IRB in order to review patient medical records for eligibility. After identification of potentially eligible patients, the primary oncologist caring for each patient will be contacted by site staff to confirm there is no reason the patient should not be contacted. If there are no objections by the physician for the patient to be contacted, staff will obtain written informed consent from the patient for participation in the study, including HIPAA release for medical record abstraction. Patients should be approached by staff during their routine clinic visit, to present the study and obtain written informed consent.

### 7.4 Patient Registration and Medical Record Abstraction

Site staff will register the consented patient per Section 4.0 and complete a brief medical record abstraction form in Rave within one week following registration.

The following information will be abstracted from the medical record: patient date of birth, gender, zip code, country of residence, race, ethnicity, method of payment (insurance status), and pre-existing comorbidities. Data specifically related to multiple myeloma and chronic lymphocytic leukemia include: date of diagnosis, comorbidities; dates of current and historical treatment initiations, current and historical treatments.

Medical abstraction will apply to the past 12 months, with some exceptions, such as date of diagnosis, which may be outside this window.

| **Medical Record Information** | **Relation to Study Aims** |
| --- | --- |
| Demographic information: date of birth, gender, country of origin, zip code, race, ethnicity, insurance type | 2.2.1 To describe the association of patient report of financial difficulty with insurance status.  2.2.5 To examine the relationship between distinct patterns of financial burden with patient report of financial difficulty, patient socio-demographics, and patient disease characteristics. |
| Primary disease (multiple myeloma and/or chronic lymphocytic leukemia) | 2.2.5 To examine the relationship between distinct patterns of financial burden with patient report of financial difficulty, patient socio-demographics, and patient disease characteristics. |
| Current and prior treatment initiation dates for MM and/or CLL | 2.2.6 To estimate the proportion of patients with MM or CLL undergoing treatment who report receiving financial support in the past 12 months. |
| General comorbidities and MM or CLL- specific comorbidities | 2.2.5 To examine the relationship between distinct patterns of financial burden with patient report of financial difficulty, patient socio-demographics, and patient disease characteristics.  2.2.10 To describe the association of financial difficulty with patient self-reported health and well-being. |

### 7.5 Patient Survey

Within eight weeks after patient registration, the patient survey will be conducted as a centralized telephone interview by study team survey staff located at UNC. Data collected will be entered into Medidata Rave. The interview will be conducted at a time that is convenient for the patient and feasible for the UNC study team survey staff. The survey will be available in both English and Spanish.

The UNC research staff conducting the phone interviews will make up to eight attempts to reach a participant the first time. A voicemail message will be left on the first and fourth call attempt, as indicated in the call script. If someone other than the patient answers the phone, they will be asked about a good time to reach the participant.

The UNC research staff will begin attempting to contact the participant within the first few days after they have been registered on study. If a participant is unable to complete the phone interview in a single phone call, the interview will be continued at a later time within the 8 week window (from the time of registration).

We expect the patient survey to take 60 minutes to complete based on study team work on other structured interviews with cancer patients (details available upon request) and will be conducted by the survey team via telephone. Data collected will be entered into Medidata Rave by the UNC study team survey staff. Questionnaires of this length have been successfully completed in previous Alliance studies.

Patients will receive a $20 Visa gift card for their participation. The gift card will be mailed to the participant following completion of the survey. The gift card will be coordinated and mailed by UNC study team survey staff. Patient survey details are contained in Section 8.2. The survey can be found in Appendix II.

8.0 **MEASURES**

### 8.1 Definition of Primary and Key Secondary Endpoints

8.1.1 **Primary endpoint**

The primary endpoint, financial difficulties, will be assessed using the European Organization for Research and Treatment of Cancer Quality of Life Questionnaire –Core 30^20,21,22^ (EORTC QLQ-C30) item #28 regarding financial difficulties with a modified recall period: “Has your physical condition or medical treatment caused you financial difficulties in the past year? [Not at all / A little / Quite a bit / Very much].” Due to the very memorable nature of financial difficulties and the typically long duration/constancy of financial difficulties, we believe a 12 month recall period is appropriate and this is supported by theoretical work on recall periods.^24^ For the primary endpoint, participant responses will be dichotomized (“Not at all” or “A little” classified as No, and “Quite a bit” and Very much” classified as Yes).

8.1.2 **Secondary endpoints**

In Secondary Objective 2.2.1, **financial difficulties** will be assessed as above with the EORTC QLQ-C30 item #28, but it will not be dichotomized.

**Current resources dedicated to patient financial navigation among participating NCORP components** will be identified via the Practice Survey (Appendix I) including financial and health literacy assessment, financial support services, patient-centered psychosocial support, and transportation and financial navigation services.

**Resources being developed for patient psychosocial, transportation and financial navigation** will be identified via the Practice Survey (Appendix I). We will use descriptive statistics to summarize the proportion of sites who plan on implementing new initiatives to aid patient navigation in psychosocial support and transportation and financial navigation, and also the details of said initiatives (i.e. timing and nature of initiatives, staff responsible and reasons for implementing new initiatives). We will also look for themes of similar services being planned in an exploratory manner.

Latent classes of **financial burden** (see secondary objective 2.2.4) will be identified as described in the statistical analysis plan. This analysis will help us in finding similar groups of patients who have the same types of financial problems, as well as sub-themes of related questions.

Patient report of receiving **financial support** is a binary item created during survey development work. It simply reflects whether or not any financial support is received. Details about the type of support will be gathered by other questionnaires.

**Patient concerns regarding treatment and costs** will be assessed via the Valuing Dimensions of the Patient Experience Questionnaire (Appendix II- See Part 1 questions 22- 36), composed of 15 items regarding health care navigation, finances, daily life, effects of treatment, and psychosocial effects, and was developed by the study team and pilot tested in preliminary work. Participants will rate each of the 3-point Likert scale items as something they were “not worried,” “somewhat worried,” or “very worried” about. For scoring purposes, these items will be assigned a value from 0 to +2. Aggregate item scores will be calculated by computing the mean Likert score rating.

**Patient health and well-being** will be assessed via the PROMIS-10, EQ-5D and the Brief Appraisal Inventory (BAI). The PROMIS-10 is composed of 10 items regarding general, physical and mental health as well as social activities, carrying out general activities, pain and fatigue. Participants will rate each on a 5-point Likert scale, and their responses will be summed and converted to a T score using standardized conversion tables ^25^ The EQ-5D is a 5-item self-reported measure focused on mobility, self-care, usual activities, pain/discomfort and anxiety/depression using a 5-point Likert scale, and their responses will be converted to a health state as per a standardized scoring algorithm^26^, ^27^(REF) Finally, the BAI is a 23-item survey, that assesses participants’ current mental state using a 5-point Liker scale. The BAI yields five composite scores, reflecting the five domains of appraisal.

A key set of analyses will examine how patients’ experience of financial difficulties are related to health status, health-related quality of life and quality of life appraisal. Inclusion of the PROMIS-10, the EQ-5D and the BAI will add context to our analysis of differences in financial difficulties and financial concerns, and will help us to better understand people’s choices in coping with financial difficulties.

### 8.2 Patient-reported measures

The Patient Survey is composed of four domains, described below. The majority of questionnaire items are from national surveys, i.e., the National Health Interview Survey (NHIS) and Medical Expenditure Panel Survey (MEPS)^23^, or from well-established patient-reported outcome instruments, i.e, the European Organization for Research and Treatment of Cancer Quality of Life Questionnaire –Core 30 (EORTC QLQ C30), the EuroQol Group EQ-5D-5L, and the PROMIS v1.2 – Global Health. Questionnaire items that were written for this survey or substantially modified for this survey have been evaluated for comprehension in study development work.

The rationale for each set of items and their source is described below, organized by domain and sub-domain. The scoring method of particular instruments is described where relevant.

All the questionnaire items have closed-ended responses. The response options include Likert scales, yes/no, categorical (select one or select all that apply), and integer values.

The recall period for the items is either “now” or “in the past 12 months”. In some cases, the recall period of the original items was modified to “in the past 12 months” in order to accommodate the study design; these changes were reviewed by a survey methodologist in accordance with guidelines published by Stull et al^24^.

The location of the questionnaire items within the patient survey (appendix 2) are noted below in parentheses at the end of each description.

8.2.1 **Domain 1: Financial Difficulty (See Appendix II Part 1)**

Patient Report of Financial Difficulty

This section contains 1 question from the European Organization for Research and Treatment of Cancer Quality of Life Questionnaire –Core 30 (EORTC QLQ C30). This question will allow us to assess if physical condition or medical treatment has caused patients financial difficulty. It is described above as the Primary Endpoint. (See Part 1 question 9)

Difficulties paying medical bills

This section contains 3 questions from the National Health Interview Survey (NHIS)^28^. These questions will allow us to assess difficulties patients and or their families have had paying any medical bills in the past 12 months. (See Part 1 questions 1-3)

Delays in treatments, foregoing recommended treatment

This section contains 4 questions from the NHIS that will allow us to assess the degree to which medical treatment and associated care has been delayed due to cost over the past 12 months. (See Part 1 questions 4-7)

Difficulties covering non-medical expenses

This section contains 1 question form the NHIS that will allow us to assess financial sacrifices made in the past 5 years due to medical related debt. (See Part 1 question 8)

Financial worries

This section contains 2 questions from the NHIS, 1 question from the Behavioral Risk Factor Surveillance System (BRFSS)^29^, 1 question from the Medical Expenditure Panel Survey (MEPS) and 1 internally created question to assess the degree of financial worry patients experience related to the cost of care. (See Part 1 questions 10-16)

Patient reported medical care utilization and out of pocket costs associated with medical care

This section contains 3 questions from Yabroff *et al* to assess different kinds of financial burden families experience because of cancer, its treatment, or lasting effects of treatment. (See Part 1 questions 17-21)

Patient concerns regarding treatment and costs

Patient priorities for treatment- and cost-related factors will be assessed using the Valuing Dimensions of the Patient Experience questionnaire. It is composed of 15 items regarding health care navigation, finances, daily life, effects of treatment, and psychosocial effects. Participants will rate each of the 3-point Likert scale items as something they were “not worried,” “somewhat worried,” or “very worried” about. For scoring purposes, these items will be assigned a value from 0 to +2. Aggregate preference scores will be calculated by computing the mean Likert score rating for each item. (See Part 1 questions 22-36)

8.2.2 **Domain 2: Patient Report of Seeking and Receiving Financial Support (See Appendix II Part 2)**

Advice regarding financial or nonfinancial support

This domain contains 2 questions developed and pilot tested by the study team. These questions allow us to assess advice regarding financial and nonfinancial support received for cancer care.

Formal support for specific purposes

This domain contains 37 questions developed and pilot tested by the study team. These questions allow us to assess the type formal support received from an agency/organization/foundation as well as gather more information about how patients were notified of this support, the process for applying for this support, and the total dollar amount of the support received.

Support from family and friends for specific purposes

This domain contains 6 questions developed and pilot tested by the study team. These questions allow us to assess the type of informal support received from family, friends, coworkers, neighbors, etc., as well as they type of support received.

8.2.3 **Domain 3: Patient Sociodemographic Indicators (See Appendix II Part 3)**

Patient Household Composition

This section contains 1 question from the MEPS. This question allows us to assess the number of people living in each subjects’ household, including children and seniors.

Patient Education Status

This section contains 1 question from the MEPS. This question allows us to assess level of education attainment.

Patient Employment Status

This section contains 7 questions from the MEPS. These questions assess current employment status, including full-time versus part-time working status and reasons for not currently working.

Household monthly income, assets

This section contains 7 questions from the MEPS. These questions assess total household income, and homeownership status.

Household Debt

This section contains 5 questions from the MEPS. These questions assess the amount and status of debt from all sources.

Patient Insurance

This section contains 3 questions from the MEPS, one of which was expanded to allow for more insurance options. These questions assess current insurance status, type of insurance held by participants and the deductible for their insurance plan.

Patient

This section contains 1 question newly developed for this survey. This question will allow us to assess the number of medications currently being taken by respondents.

Patient Financial Literacy

This section contains 5 questions from the Rand Corporation’s American Life Panel Survey^30^. These questions assess financial literacy. Each item is a brief math problem that requires basic familiarity with interest rates; the questions are multiple choice and have a correct answer.

Patient demographics

This section contains 7 MEPS questions that assess main residence, sex, race and marital status.

8.2.4 **Domain 4: Health and Well-being (See Appendix II Part 4)**

Self-reported health

This section contains EuroQol Group’s EQ-5D-5L^31,32^ that assess patients’ health status across 5 dimensions: mobility, self-care, usual activities, pain/discomfort, and anxiety/depression. It is composed of 5 items with Likert scale response options. Index scores will be created according to the User Guide^33^.

Self-reported health and quality of life

This section contains the Patient-Reported Outcomes Measurement Information System (PROMIS) Scale v1.2 – Global Health^34,35^, composed of 10 items with Likert scale response options. It measures key areas of quality of life, including general health, fatigue, pain, physical functioning, and negative affect. It will be scored according to the Health Measures Scoring Service^36^. A score of 0 to 100 is generated, where higher scores indicated better health.

Concerns and priorities that bear on patients’ quality of life

This section contains Brief Appraisal Inventory (BAI) developed by Rapkin, Schwartz and colleagues^37,38,39^. The BAI was developed with support from PCORI, as a method to improve the ability to understand how patients’ ways of appraising quality of life influence their responses to patient-reported outcomes, such as financial difficulty. The BAI is composed of 23 statements, each describing concerns shown in earlier studies to moderate the impact of health status and stressful events on patient-reported outcomes in heterogeneous samples of chronic disease patients. Each statement is scored on a five-point Likert scale, indicating the extent that it is presently on the persons’ mind as they responded to the survey today, from (1) always to (5) never. Items are combined to yield five patterns of appraisal: accomplishing goals and solving problems in roles, living situation and relationships; worries about health and healthcare; maintaining independence and spending quality time with family; maintaining a calm, peaceful and active lifestyle; and spiritual growth and

altruistic activities. Items are recoded and averaged so that higher scores reflect a greater focus on a given pattern of appraisal.

9.0 **STATISTICAL CONSIDERATIONS**

### 9.1 Study Design

This is a hypothesis driven, observational, cross-sectional study, entailing the collection of survey data from patients and from the NCORP sites recruiting patients into the study. NCORP sites have dedicated staff to help study teams pursue approved CCDR research.

**Table 1: Survey Items Measuring Each Primary and Key Secondary Endpoint, by Study Objective**

| **Study Objective** | **Primary and Key Secondary Endpoints**  **/ Domains of Interest** | **Survey Items Measuring the Domain of Interest** | **Analysis Plan** |
| --- | --- | --- | --- |
| 2.1 To estimate the proportion of patients with MM and/or CLL who report experiencing financial  difficulty in the past 12 months. | Financial difficulties | EORTC QLQ-C30 item #28 | Wilson score confidence interval (95%) |
| 2.2.1 To describe the association of  patient report of financial difficulty with insurance status. | Financial difficulties | EORTC QLQ-C30 item #28 | Mann-Whitney U test |
| 2.2.2 To describe the association of patient report of financial difficulty with receiving treatment at practices that report offering patients financial guidance through navigators or social workers, and with socioeconomic status. | Financial difficulties | EORTC QLQ-C30 item #28 | Logistic regression and latent class analysis models with covariates as listed in section 9.4.3 |
| 2.2.3 To describe the types of psychosocial, transportation and financial navigation interventions sites are developing. | Financial support | Site-reported plans of developing psychosocial, transportation and financial navigation services found in  the Practice Survey (questions 36-50) | Descriptive statistics |
| 2.2.4 To identify distinct patterns of financial burden among patients undergoing treatment for MM and/or CLL. | Financial burden | Patient reported difficulties paying medical bills (4 binary items), delays or foregoing treatment (4 binary items), difficulties covering non- medical expenses due to costs of treatment (2 binary items), and financial worries (5  binary items) | Exploratory latent class analysis models with covariates as listed in section 9.4.4 |
| 2.2.5 To examine the relationship between distinct patterns of financial burden with patient report of financial difficulty, patient socio- demographics, and patient disease characteristics. | Financial burden | Patient reported difficulties paying medical bills (4 binary items), delays or foregoing treatment (4 binary items), difficulties covering non- medical expenses due to costs  of treatment (2 binary items), | Logistic regression and latent class analysis models with covariates as listed in section 9.4.5 |

|  |  | and financial worries (5 binary items) |  |
| --- | --- | --- | --- |
| 2.2.6 To estimate the proportion of patients with MM and/or CLL undergoing treatment who report receiving financial support in the  past 12 months. | Financial support | Proportion of patients who report receiving financial support (yes/no) | Summary statistics |
| 2.2.7 To describe the association of patient report of receiving financial support with receiving treatment at practices offering patients financial guidance through navigators or social workers, and with  socioeconomic status. | Financial support | Proportion of patients who report receiving financial support (yes/no) | Logistic regression with covariates as specified in section 9.4.7 |
| 2.2.8 To describe the magnitude of patient concerns regarding treatment and costs of care. | Patient concerns regarding treatment and costs | Aggregate scores of the Valuing Dimensions of the Patient Experience  Questionnaire (Appendix II- See Part 1 questions 22-36) | Summary statistics |
| 2.2.9 To describe the association of patient concerns regarding treatment and costs of care with patient socio-demographics, disease and practice characteristics. | Patient concerns regarding treatment and costs | Aggregate scores of the Valuing Dimensions of the Patient Experience Questionnaire (Appendix II- See Part 1 questions 22-36) | Two-sample t-tests and latent class analysis models with covariates as listed in section  9.4.9 |
| 2.2.10 To describe the association of financial difficulty with patients self-reported health and well-being. | Patient health and well being | Aggregate scores of the PROMIS-10, EQ-5D and the BAI | Logistic regression with covariates as  specified in section 9.4.10 |

### 9.2 Primary objective: To estimate the proportion of patients with MM and/or CLL who report experiencing financial difficulty in the past 12 months.

Primary endpoint: Financial difficulties will be assessed using the EORTC QLQ-C30 item #28 regarding financial difficulties with a modified recall period: “Has your physical condition or medical treatment caused you financial difficulties in the past year? [Not at all / A little / Quite a bit / Very much]”. Participant responses will be dichotomized (“Not at all” or “A little” classified as No, and “Quite a bit” and Very much” classified as Yes).

Secondary endpoint: Because the primary objective is to estimate a proportion, the sample size for this study is based on power calculations for secondary objective 2.2.1. Here we describe the expected level of precision for the estimated proportion. Based on a previously fielded survey of financial distress among a convenience sample of US MM patients, we expect 50% or more of the patients surveyed will report experiencing financial difficulties. We have calculated expected lower and upper bounds of a 95% confidence interval (CI) for various proportions for a sample of size N using Wilson scores (Table 3). While we expect there to be some loss of precision due to study design (patient clustering within sites), we expect this effect to be minimal. With a sample size of 500 participants, 95% confidence intervals will be +/- 4.4%. We conservatively estimate 20% missing data (survey non-response among enrolled patients), which would provide an analytic sample of N =400 and 95% confidence intervals of approximately +/- 5%.

**Table 2: Wilson score confidence intervals (95%) for estimated proportions by sample size**

| N | Estimated proportion | | | | | | | | | |
| --- | --- | --- | --- | --- | --- | --- | --- | --- | --- | --- |
|  | 10% | | 20% | | 50% | | 70% | | 80% | |
|  | Lower | Upper | Lower | Upper | Lower | Upper | Lower | Upper | Lower | Upper |
| 300 | 0.070 | 0.139 | 0.159 | 0.249 | 0.444 | 0.556 | 0.646 | 0.749 | 0.751 | 0.841 |
| 500 | 0.077 | 0.129 | 0.167 | 0.237 | 0.456 | 0.544 | 0.658 | 0.739 | 0.763 | 0.833 |
| 700 | 0.080 | 0.124 | 0.172 | 0.231 | 0.463 | 0.537 | 0.665 | 0.733 | 0.769 | 0.828 |

### 9.3 Plan for missing data and other statistical considerations

Queries will be sent to sites for missing data with the medical chart abstraction and the Practice Survey. Survey participation rates and rates of missing data within each survey (item level missingness) will be optimized in this study by the use of live telephone interviewers to administer the survey. They will contact participants and endeavor to obtain complete/valid responses to all survey items; reasons for missing data will be documented. Missing data patterns found within the collected data will be summarized and reported. The missing completely at random (MCAR) assumption will be tested using a nonparametric test of MCAR.^40^ Data will be listwise deleted before further analysis if the MCAR test returns a nonsignificant result and an adequate number of cases remain after listwise deletion (to maintain adequate power). Otherwise, a weaker assumption of missing at random will be made, and the missing values will be multiply imputed 100 times using multivariate imputation by chained equations (the mice package in R).^41,42^. All subsequent analyses will be repeated 100 times, and the results will be pooled according to Rubin’s rules.^43^ All study analyses will be conducted with alpha = 0.05 and two-tailed tests unless otherwise noted. If patient report of financial difficulty or of receiving financial support is strongly associated with whether their practice offers financial navigation, we will consider conducting analyses of patient data that clusters by practice. Furthermore, we will apply a recently developed macro which uses 20 different imputation approaches to missing data that was developed by the Statistics and Data Center (SDC). This approach involves the production of imputed datasets that are subsequently re-analyzed alongside the initial data, and compare the results across the spectrum of approaches^44^.

### 9.4 Secondary objectives

9.4.1 **To describe the association of patient report of financial difficulty with insurance status.**

This key secondary objective will be the basis of sample size calculations for this study. We hypothesize that among patients age 65+ with Medicare, those with Medicaid will report less financial difficulty than those without Medicaid. Because the measure of patient report of financial difficulty is specific to “physical condition or medical treatment” we anticipate that patients with Medicaid will report less financial difficulty because they face much lower out of pocket costs for their MM and/or CLL treatment.

In addition, we hypothesize that among patients under the age of 65, those with Medicaid and/or Medicare (qualified due to disability) will report less financial difficulty that those with only commercial insurance. Again, we anticipate that patients with Medicaid and/or Medicare will face much lower out of pocket costs than those who are only commercially insured.

The outcome, patient report of financial difficulty, will be measured as in the primary objective, but the responses will not be dichotomized -- the four response categories will be preserved. Given that the outcome variable is measured on an ordinal scale, each of the two hypotheses (one for patients 65+ and one for patients <65) will be tested using a Mann-

Whitney U test to compare the two groups defined by insurance status in terms of the difference in their responses on the outcome.

With a total recruitment size of 500 and a completion rate of 80%, we expect a valid sample size of N = 400. Based on the age distribution of the target population, we also expect that 70% of the valid cases would be aged at 65 or above, and 30% would be aged below 65 (N65+ = 280 and N<65 = 120). Within the elder group, about 30% of participants are expected to belong to Group A (N65+A = 84) (those with Medicare and Medicaid), and the remaining 70% to Group B (N65+B = 196) (those with Medicare but without Medicaid). Within the younger age group, the sample size ratios of Group A (those with Medicare and/or Medicaid) and Group B (those with only commercial insurance) are expected to be 40% and 60% respectively (N<65A = 48 and N<65B = 72). With a medium effect size of

0.5 and a nominal alpha criterion (α = 0.05), a two-tailed Mann-Whitney U test would achieve a statistical power of 0.962 for the group comparison within the elder group, and a power of 0.739 within the younger group. Further analysis revealed that sample sizes of N<65A = 56 and N<65B = 84 would be required, if we were to expect a power above 0.80. An effect size of 0.5 is appropriate for this analysis because it is considered a “moderate” effect size^47^ and because in the field of health-related quality of life research it is widely supported as a ‘minimally important difference’ between two groups.^45^

9.4.2 **To describe the association of patient report of financial difficulty with receiving treatment at practices that report offering patients financial guidance through navigators or social workers, and with socioeconomic status.**

Responses to our updated practice survey will be used to identify practices that offer patients financial guidance through navigators or social workers. The question in our updated practice survey are questions 23-36 (Appendix I) These questions were drafted based on the 2015 CCDR Landscape Assessment question #23 and pilot tested in preliminary work. Logistic regression will be used to test the hypothesis that individuals with MM or CLL treated at practices reporting that they offer patients financial guidance through navigators or social workers will be less likely to report financial difficulty compared to those treated at practices without these resources. The model will control for disease/treatment characteristics and indicators of patient socioeconomic status.

We will also test the hypothesis that individuals with MM or CLL and low to moderate socioeconomic status will be more likely to report financial difficulty compared to those with high socioeconomic status. Patient socioeconomic status will be categorized into low, medium, and high based on the results of latent class analysis of the following indicators: insurance coverage and generosity, income and assets, educational attainment and financial literacy. An exploratory latent class analysis (LCA) will be used to assess different patterns of socioeconomic status. More specifically, we will construct basic LCA models with the number of latent classes varying from one to five (or more). These models (with different number of latent classes) will be examined for fit and parsimony (AIC, BIC, G2 likelihood- ratio statistic), interpretability, and sparseness of the indicator variables to decide the optimal number of latent classes^.42^ Next, latent class membership will be used to predict financial difficulty using the technique proposed by Lanza et al, (2013)^46^. This technique includes patient report of financial difficulty as a latent class predictor within a multinomial logistic regression in addition to the original LCA measurement model.

9.4.3 **To identify distinct patterns of financial burden among patients undergoing treatment for MM and/or CLL.**

This analysis will identify distinct patterns of reported financial burden. Because patients will make financial decisions regarding health care expenses based on their resources and

ability to make tradeoffs, and in many cases indicators of financial burden such as incurring debt versus selling assets are reciprocal, we expect considerable heterogeneity in how patients experience financial burden. Patterns will be assessed utilizing measures of patient reported difficulties paying medical bills (4 binary items), delays or foregoing treatment (4 binary items), difficulties covering non-medical expenses due to costs of treatment (2 binary items), and financial worries (5 binary items). Exploratory LCA will be conducted consistent with the methods described in secondary objective #1.

9.4.4 **To examine the relationship between distinct patterns of financial burden with patient report of financial difficulty, with patient socio-demographics, and with patient disease characteristics.**

This analysis will identify the relationship between distinct patterns of financial burden (latent classes) identified in secondary objective #2 with patient report of financial difficulty. Financial difficulty will be modeled as a latent class predictor within a multinomial logistic regression in addition to the original LCA measurement model.

Additional analyses will examine whether factors like demographics, type of insurance or household composition, and disease characteristics will distinguish individuals with different patterns of financial burden. Univariate analyses will be conducted to identify the association between each variable and the latent classes, and promising variables will be added to the optimal LCA in multinomial logistic models. This analysis will help us in finding similar groups of patients who have the same types of financial problems, as well as sub-themes of related questions.

9.4.5 **To estimate the proportion of patients with MM and/or CLL undergoing treatment who report receiving financial support in the past 12 months.**

The proportion of patients who report receiving financial support (yes/no) in the past 12 months will be described. Additional descriptive analyses will examine the proportion of patients who report receiving non-financial support (e.g., transportation, groceries), the proportion of patients who applied for financial support or who asked family or friends for financial support, and the estimated amount of time spent paying for care (e.g. reviewing bills, talking with insurance companies, talking with financial counselors, applying for financial support, and asking/organizing non-financial support).

9.4.6 **To describe the association of patient report of receiving financial support with receiving treatment at practices offering patients financial guidance through navigators or social workers, and with socioeconomic status.**

Logistic regression with receipt of financial support in the past 12 months (yes/no) as the dependent variable will be used to test the hypothesis that individuals with MM and/or CLL will be more likely to report receipt of financial support if they are treated at practices reporting that they offer patients financial guidance through navigators or social workers compared to those treated at practices without these resources. The model will control for disease/treatment characteristics and indicators of patient socioeconomic status. We will also test the hypothesis that individuals with MM and/or CLL and low to moderate socioeconomic status will be more likely to report receiving financial support compared to those with high socioeconomic status. Patient report of receiving financial support will be modeled as a predictor of latent classes of socioeconomic status within a multinomial logistic regression in addition to the original LCA measurement model.

9.4.7 **To describe the magnitude of patient concerns regarding treatment and costs of care.**

Aggregate scores of the Valuing Dimensions of the Patient Experience Questionnaire (Appendix II- See Part 1 questions 22-36) will be summarized and analyzed at the item- level, and additional analyses will be conducted by evaluating each item relative to one another. It is hypothesized that the level of concern for each factor will not be the same.

9.4.8 **To describe the association of patient concerns regarding treatment and costs of care with patient socio-demographics, disease characteristics, and practice characteristics.**

Aggregate scores of patient concerns obtained in objective #6 will be stratified by demographics (e.g. gender, race/ethnicity), socioeconomic characteristics (e.g. education, income), disease characteristics (e.g. MM, CLL), and practice-specific factors (e.g. presence of a social worker/patient navigator). Differences in Likert scores between groups will be tested using t-tests. Adjustment for multiple comparisons will not be conducted, which is consistent with social science and/or preference based research. It is hypothesized that individuals with MM and/or CLL will be less likely to report worries if they are treated at practices reporting that they offer patients financial guidance through navigators or social workers compared to those treated at practices without these resources. In addition, a latent class analysis will be conducted to explore clusters of individuals who are similar based on patterns of the outcome variable. This approach isolates the respondent characteristics that are correlated with the differences in reported concerns. With a sample size of approximately N=500, multivariate statistical analyses will be used to analyze the characteristics of the latent classes and to identify 2-3 subgroups. It is hypothesized that identifiable differences in the concerns of these multiple latent classes will be detected.

9.4.9 **To describe the association of financial difficulty with patients self-reported health and well-being**

Aggregate scores of the PROMIS-10, EQ-5D-5L and the BAI will be utilized as covariates in a logistic regression model to assess if there is any association with them and patient- reported financial difficulty. Confirmatory factor analysis of the BAI will be conducted prior to using it in the analysis.

9.4.10 **Considerations for potential biases introduced by site-related autocorrelation and clustering.**

There may be concern regarding clustering of results within a given site which may introduce bias into the estimates of financial toxicity. There is no evidence that the clustering in this study will be of any particular concern, and that recent experiences with Alliance cluster randomized designs have indicated autocorrelation coefficients below 0.05 so that the amount of bias introduced by such clustering is so small so as to be clinically meaningless. Nonetheless, site factors may induce autocorrelation in this study, due to local variation in financial supports and resources available to patients as well as community differences in costs of living and available health insurance programs.

### 9.5 Monitoring

As requested by the NCI, an interim analysis will be done after the first 30 patients have completed the survey OR the initial group of four sites have completed their pilot (see Section 10.2), whichever comes first. The interim report will include: 1) range and median time to survey completion; 2) an item-by-item count of missing data; and 3) a count of individuals who decline to participate and reason for not participating. These items will be collected along with the other questionnaire items. The CCDR Steering Committee asked that, prior to opening the study beyond the pilot sites, NCI use the report to assess feasibility and work with the study

team to address any concerns. The Committee expects NCI to provide an update on study progress when the report is received.

### 9.6 Reporting

This study will be monitored by the Alliance Data and Safety Monitoring Board (DSMB), an NCI-approved functioning body. Reports containing efficacy, adverse event, and administrative information will be provided to the DSMB every 6 months as per NCI guidelines. Reports from these meetings will be made available to the study chair, statistician, and participating institutions.

Results Reporting on ClinicalTrials.gov: At study activation, this study will have been registered within the “ClincialTrials.gov” website. The Primary and Secondary Endpoints along with other required information for this study will be reported on ClinicalTrials.gov.

### 9.7 Inclusion of Women and Minorities

This study will not exclude potential subjects from participating in this study solely on the basis of ethnic origin, gender or socioeconomic status. Efforts will be made to enroll individuals of all genders, races, and ethnic backgrounds. We do not expect a differential effect of the intervention by gender, race, or ethnicity. Predictions of accrual and anticipated accrual in subgroups defined by gender and race is:

| . Racial Categories | DOMESTIC PLANNED ENROLLMENT REPORT | | | | |
| --- | --- | --- | --- | --- | --- |
|  | Ethnic Categories | | | | |
|  | Not Hispanic or Latino | | Hispanic or Latino | | Total |
|  | Female | Male | Female | Male |  |
| American Indian/Alaska Native | 5 | 3 | 0 | 0 | 8 |
| Asian | 5 | 3 | 0 | 0 | 8 |
| Native Hawaiian or Other Pacific Islander | 5 | 3 | 0 | 0 | 8 |
| Black or African American | 58 | 41 | 7 | 6 | 112 |
| White | 162 | 142 | 24 | 19 | 347 |
| More Than One Race | 5 | 4 | 5 | 3 | 17 |
| Total | 240 | 196 | 36 | 28 | 500 |

10.0 **GENERAL REGULATORY CONSIDERATIONS AND CREDENTIALING**

### 10.1 Site Credentialing/Eligibility requirements

Institutions that are interested in participating in this study must meet eligibility requirements outlined in Section 3.3. Interested sites must first contact the Study Chair, Dr. Rena Conti to review study requirements and procedures and to identify who will be responsible for the study at the institution.

Sites should complete the information required in the Institutional Approval Document posted along with Supplementary materials on the Alliance and CTSU Websites, and send it to Dr. Conti at the email address listed on the protocol cover page. The document will ask for the following information: Name of the site, CTEP Site Code, CTEP Site Code of Main NCORP and contact information (Name and email) of site staff who will complete the practice survey in Medidata Rave.

To approve the site for participation, the study chair will return this document to the institution with a signature of approval. As soon as the site receives IRB approval, it must submit this Institutional Approval Document to the Alliance Registration Office via fax (507-284-0885) or email at The registration office will register the site to the study and send confirmation to the site via email within 1 business day, in order to be invited to enter site level data in Medidata Rave. The study will not use OPEN for site enrollment. The contact person listed on the institutional approval document will be sent an invitation to enter site level data in Medidata Rave. In addition, the site will also submit this document to the CTSU Regulatory portal per Sections 4.2.2 and 4.2.3 in order to begin patient enrollment. Please note that the Institutional Approval Document must be sent to the reg/rando office as soon as IRB approval is obtained, as the Practice Survey will need to be completed within 8 weeks (56 days) of this date (the date that the site sends the Institutional Approval Document to the reg/rando office is the “site enrollment date” for the purposes of Rave data entry).

### 10.2 Site Selection and Site Recruitment

NCORP sites serve as the sampling frame for this study. We will focus priority recruitment efforts on some of the larger sites to increase the efficiency of obtaining adequate patient sample sizes. Sites will be solicited to participate in the study through a combination of written and in- person requests by the study team, CCDR leadership and site PI and administrator outreach efforts. We have prepared a two-page summary to aid site recruitment efforts, including: study objectives, innovation, timeline, roles and responsibilities. We expect the study to have a staged rollout where we first initiate recruitment among the 4 pilot sites that have already agreed to participate, then recruit sites to meet geographic diversity and recruitment goals.

Once sites have been recruited to the study, site staff will initiate patient recruitment. We have engaged with individuals at primarily minority-serving sites and larger sites to increase the likelihood of their participation. We will evaluate recruitment targets with attention paid to the inclusion of adequate representation across minority and socioeconomically disadvantaged groups.

Time commitment of patients, research staff, physicians or other study participants:

A guiding principle in designing the study is minimization of staff and patient time commitments. Based on our pilot testing, we expect each recruited site staff commitments to entail the following: 1- 5 hours for identification of all eligible patients; 10-15 minutes for each patient consent and 30-45 minutes for each patient registration, including medical record abstraction. We expect the patient survey to take 60 minutes to complete based on study team work on other structured interviews with cancer patients (details available upon request) and

will be conducted by the survey team via telephone. We expect 45 minutes will be required for practice survey completion among practices recruiting eligible patients.

### 10.3 Waivers of patient consent

### 10.3.1 Pre-screening

A waiver of informed consent for the prescreening described in Section 7.3 is justified for the following reasons:

- Pre-screening eligibility review of medical records by clinical research staff does not adversely affect the rights or welfare of subjects because medical record information is screened to establish preliminary eligibility prior to approaching a potential subject about a study,
- Pre-screening eligibility review of medical records cannot practicably be done without the waiver due to the number of services participating in which patients may be eligible. Pre-screening eligibility decreases the burden on patients of introducing a research study to subjects who can easily be identified through pre-screening activities as not eligible for a research study,
- Pre-screening eligibility review of medical records involves no more than minimal risk because the person accessing the information has undergone training in confidentiality of medical records, and the records are viewed solely to pre-screen for eligibility criteria and only minimal information found in the pre-screening process will be recorded for research purposes. Names of patients (and other unique identifiers) who are deemed ineligible will not be recorded.
10.3.2 **Waiver of element of informed consent**

We request a waiver of the specific element of consent regarding medical injury to patient. This study does not involve medical treatment, and it is not expected that patients will suffer medical injury due to participation in this trial. We have therefore removed the “What happens if I am injured because I took part in this study” section of the NCI Model Consent Template. Omission of this information does not constitute greater than minimal risk and would not adversely affect the patient.

11.0 **REFERENCES**

1. Ubel P et al. Full Disclosure – Out-of-Pocket-Costs as Side Effects; N Engl J Med 2013; 369:1484- 1486. <http://www.nejm.org/doi/full/10.1056/NEJMp1306826>
2. Zulllig LI, Peppercorn JM, Schrag D, Taylor DH, Lu Y, Samsa G, Abernethy AP, Zafar SY. Financial distress, use of cost-coping strategies, and adherence to prescription medication among patients with cancer. JOP Nov 1, 2013:60s- 63s. <http://jop.ascopubs.org/content/9/6S/60s.abstract>
3. Stump TK, Eghan N, Egleston BL, Hamilton O, Pirollo M, Schwartz JS, Armstrong K, Beck JR, Meropol NJ, Wong YN. Cost concerns of patients with cancer. JOP Sep 1, 2013:251-7. <http://jop.ascopubs.org/content/9/5/251.abstract>
4. National Survey of Households Affected by Cancer; USA Today, Kaiser Family Foundation, and Harvard School of Public Health 2006: Kaiser Family Foundation Publication #7590.

https://kaiserfamilyfoundation.files.wordpress.com/2013/01/7591.pdf

5. Felder TM, Bennett CL. Special Series: State of Oncology Practice - Commentary: Can Patients Afford to Be Adherent to Expensive Oral Cancer Drugs?: Unintended Consequences of Pharmaceutical Development JOP Nov 1, 2013:64s-66s. <http://jop.ascopubs.org/content/> early/2013/09/17/JOP.2013.001167.full.pdf
6. Shankaran V, Ramsey S. Addressing the Financial Burden of Cancer Treatment: From Copay to Can’t Pay. JAMA Oncol. Published online April 09, 2015. doi:10.1001/jamaoncol.2015.0423.
7. PDQ® Adult Treatment Editorial Board. PDQ Financial Toxicity and Cancer Treatment. Bethesda, MD: National Cancer Institute. Available at: <http://www.cancer.gov/about-cancer/managing-> care/financial-toxicity-hp-pdq.
8. <http://www.asco.org/advocacy/asco-action-brief-value-cancer-care>
9. PDQ® Adult Treatment Editorial Board. PDQ Financial Toxicity and Cancer Treatment. Bethesda, MD: National Cancer Institute. Available at: <http://www.cancer.gov/about-cancer/managing-> care/financial-toxicity-hp-pdq.
10. Einav L, Finkelstein A, Ryan S, Schrimpf P, Cullen M. Selection on Moral Hazard in Health Insurance. The American Economic Review, 2016:103(1), 178-219.
11. Handel B. Adverse Selection and Inertia in Health Insurance Markets: When Nudging Hurts. The American Economic Review, 2013:103(7), 2643-2682.
12. Levy H,, DeLeire T. What Do People Buy When They Don't Buy Health Insurance and What Does That Say about Why They Are Uninsured? Inquiry 2008:45(4): 365-79.
13. Kowalski A. The Early Impact of the Affordable Care Act, State by State. Brookings Papers on Economic Activity, 2014:277-333.
14. Einav L, Jenkins M, Levin J. The impact of credit scoring on consumer lending. The RAND Journal of Economics, 2013:44(2), 249-274.
15. Chetty R, Looney A, Kroft K.. Salience and Taxation: Theory and Evidence. The American Economic Review, 2009:99(4),1145-1177.
16. Liebman JB, Luttmer EFP. The Perception of Social Security Incentives for Labor Supply and Retirement: The Median Voter Knows More Than You’d Think. Tax Policy and the Economy 2012:26.1;1-42.
17. Howard D, Bach PB, Berndt ER, Conti RM. Pricing in the Market for Anticancer Drugs. Journal of Economic Perspectives. 2015;29(1,Winter):139–162. .
18. <http://www.skainfo.com/database_finder.php?select=Physicians>
19. https://seer.cancer.gov/data/citation.html
20. EORTC Quality of Life: EORTC QLQ-C30. [http://groups.eortc.be/qol/eortc-qlq-c30.](http://groups.eortc.be/qol/eortc-qlq-c30) Accessed January 2, 2018.
21. Aaronson NK, Ahmedzai S, Bergman B, Bullinger M, Cull A, Duez NJ, Filiberti A, Flechtner H, Fleishman SB, de Haes JC, et al. The European Organization for Research and Treatment of Cancer QLQ-C30: A quality-of-life instrument for use in international clinical trials in oncology. J Natl Cancer Inst. 1993 Mar 3; 85(5): 365-376.
22. Hjermstad MJ, Fossa SD, Bjordal K, Kaasa S. Test/retest study of the European Organization for Research and Treatment of Cancer Core Quality-of-Life Questionnaire. J. Clin. Oncol. 1995 May;13(5): 1249-1254.
23. Agency for Healthcare Research. Medical Expenditure Panel Survey: MEPS Survey Questionnaires. https://meps.ahrq.gov/mepsweb/survey_comp/survey.jsp. Revised July 14, 2017.

Accessed December 12, 2017

24. Stull DE, Leidy NK, Parasuraman B, Chassany O. Optimal recall periods for patient-reported outcomes: challenges and potential solutions. Curr Med Res Opin. 2009 Apr;25(4):929-42. doi: 10.1185/03007990902774765.
25. <http://www.healthmeasures.net/images/PROMIS/manuals/PROMIS_Global_Scoring_Manual.pdf>
26. <https://euroqol.org/wp-content/uploads/2016/09/EQ-5D-5L_UserGuide_2015.pdf>
27. https://link.springer.com/content/pdf/10.1007%2Fs11136-017-1722-2.pdf
28. Centers for Disease Control and Prevention. National Health Interview Survey: NHIS Data, Questionnaires and Related Documentation. https://[www.cdc.gov/nchs/nhis/data-questionnaires-](http://www.cdc.gov/nchs/nhis/data-questionnaires-) documentation.htm. Revised October 4, 2017. Accessed January 2, 2018.
29. Centers for Disease Control and Prevention. Behavioral Risk Factor Surveillance System: BRFSS Questionnaires. https://[www.cdc.gov/brfss/questionnaires/index.htm.](http://www.cdc.gov/brfss/questionnaires/index.htm) Revised August 25, 2017.

Accessed January 2, 2018

30. RAND American Life Panel. Well Being 64 – Basic Financial Literacy. https://alpdata.rand.org/index.php?page=data&p=showmodule&syid=64&meid=3. Accessed January 2, 2018
31. EuroQol. EQ-5D: EQ-5D-5L. <https://euroqol.org/eq-5d-instruments/eq-5d-5l-about/> Accessed January 2, 2018.
32. Van Reenen, M., Janssen, B. EQ-5D-5L User Guide: Basic information on how to use the EQ-5D- 5L instrument. 2015EuroQol Research Foundation. Netherlands: EuroQol Research Foundation, 2015. https://euroqol.org/wp-content/uploads/2016/09/EQ-5D-5L_UserGuide_2015.pdf.
33. <https://euroqol.org/wp-content/uploads/2016/09/EQ-5D-5L_UserGuide_2015.pdf>
34. 31 Health Measures – Patient-Reported Outcomes Measurement Information System (PROMIS). [http://www.healthmeasures.net/explore-measurement-systems/promis.](http://www.healthmeasures.net/explore-measurement-systems/promis) Accessed January 2, 2018.
35. Hays, R. D., Bjorner, J. B., Revicki, D. A., Spritzer, K. L., & Cella, D. (2009). Development of physical and mental health summary scores from the patient-reported outcomes measurement information system (PROMIS) global items. Quality of Life Research, 18(7), 873–880
36. <http://www.healthmeasures.net/score-and-interpret/calculate-scores>
37. Schwartz, C. E., Finkelstein, J. A., & Rapkin, B. D. (2017). Appraisal assessment in patient- reported outcome research: methods for uncovering the personal context and meaning of quality of life. Quality of Life Research, 1-10.
38. Rapkin BD, Garcia I, Michael W, Zhang, J., Schwartz CE. Distinguishing appraisal and personality influences on quality of life in chronic illness: Introducing the Quality-of-Life Appraisal Profile version 2. Quality of Life Research, 2017. Published online first June 7, 2017. DOI: 10.1007/s11136-017-1600-y.
39. Rapkin BD, Garcia I, Michael W, Schwartz CE. Development of practical outcome measures to account for individual differences and temporal changes in quality-of-life appraisal: The Brief

Appraisal Profile. Quality of Life Research, 2017. Published online first November 10, 2017. DOI: 10.1007/s11136-017-1722-2

40. Jamshidian, M., & Jalal, S. (2010). Tests of homoscedasticity, normality, and missing completely at random for incomplete multivariate data. Psychometrika, 75(4), 649-674. doi:10.1007/s11336- 010-9175-3
41. van Buuren, S., & Groothuis-Oudshoorn, K. (2011). Mice: Multivariate imputation by chained equations in R. Journal of Statistical Software, 45(3), 1-67. doi:10.18637/jss.v045.i03
42. R Core Team. (2016). R: A language and environment for statistical computing. R Foundation for Statistical Computing, Vienna, Austria. URL <http://www.R-project.org/>
43. Rubin, D. B. (1987). Multiple Imputation for Nonresponse in Surveys. New York: John Wiley & Sons.
44. Jefrey L. Huntington, Amylou Dueck. Handling Missing Data. Current Problems in Cancer, Vol. 29, Issue 6, p317–325, November-December, 2005
45. Collins, L. M., & Lanza, S. T. (2010). Latent class and latent transition analysis: With applications in the social, behavioral and health sciences. Hoboken, NJ: Wiley.
46. Lanza ST, Tan X, Bray BC: Latent class analysis with distal outcomes: a flexible model based approach. Structural Equation Modeling 2013; 20: 1–26.
47. Cohen J. Statistical Power Analysis for the Behavioral Sciences, 2nd Edition. Lawrence Erlbaum, 1998.
48. Norman GR, Sloan JA, Wyrwich KW: Interpretation of changes in health-related quality of life: the remarkable universality of half a standard deviation. Med Care 41(5):582-592, 2003
    12.0 **MODEL INFORMED CONSENT FORM**

**Study Title for Participants:**

Measuring Financial Difficulty in Patients with Blood Cancers

**Official Study Title for Internet Search on** [**http://www.ClinicalTrials.gov**](http://www.clinicaltrials.gov/)**:**

Protocol Alliance A231602CD: Assessing Financial Difficulty in Patients with Blood Cancers

**Overview and Key Information** **What am I being asked to do?**

We are asking you to take part in a research study. This study has public funding from the National Cancer Institute (NCI), part of the National Institutes of Health (NIH) in the United States Department of Health and Human Services. We do research studies to try to answer questions about how to prevent, diagnose, and treat diseases like cancer.

We are asking you to take part in this research study because you have either chronic lymphocytic leukemia and multiple myeloma or both.

**Taking part in this study is your choice.**

You can choose to take part or you can choose not to take part in this study. You also can change your mind at any time. Whatever choice you make, you will not lose access to your medical care or give up any legal rights or benefits.

This document has important information to help you make your choice. Take time to read it. Talk to your doctor, family, or friends about the risks and benefits of taking part in the study. It’s important that you have as much information as you need and that all your questions are answered. See the “Where can I get more information?” section for resources for more clinical trials and general cancer information.

**Why is this study being done?**

This study is being done to answer the following question:

How often do financial problems happen for patients with your type of cancer?

We are doing this study because we wish to better understand patients’ financial experiences related to their cancer treatment. We are also interested in identifying ways to improve patient access to resources at their treatment sites.

**What is the usual approach to use of medical information for research?**

Hospitals or doctor’s offices usually use a “Release of Medical Information” form to get medical information from patients. For this study, we are using a consent form that describes what type of information we want from your medical records. Your signing the consent form gives us permission to use this information from your medical records along with your answers to the questionnaires for research.

**What are my choices if I decide not take part in this study?**

Your decision to participate (or not to participate) in this research study will NOT affect your cancer treatment. If you decide not to take part in this research study, you have other choices. For example:

- You can get treatment for your cancer without being on a study
- You may choose to take part in a different study, if one is available

**What will happen if I decide to take part in this study?**

If you decide to take part in this study, you will be asked to complete a telephone survey within 8 weeks after joining the study. You will be asked to provide your name, and telephone number so that the study team survey staff can call you to conduct the survey. We think it will take about one hour to complete the telephone survey. More information about this survey is provided below.

**What are the risks and benefits of taking part in this study?**

There are both risks and benefits to taking part in this study. It is important for you to think carefully about these as you make your decision.

**Risks**

We want to make sure you know about a few key risks right now. We give you more information in the “What risks can I expect from taking part in this study?” section.

- You may feel uncomfortable being asked about your physical and emotional health and your finances.
- There is a small risk that your study information could become known to someone who is not involved in performing or monitoring this study. However, we make every effort to protect your privacy.

**Benefits**

You are not expected to have any direct medical benefit from participating in this study.

However, the information you provide will help us to better understand the financial impact of cancer and learn about ways to help other patients like you avoid financial problems during treatment.

**If I decide to take part in this study, can I stop later?**

Yes, you can decide to stop taking part in the study at any time.

If you decide to stop, let your study doctor know as soon as possible. You also may choose to stop participation or skip any questions in the survey that you do not feel comfortable answering.

Any data collected before you decide to withdraw will not be used for research. There will be no further follow up or interaction required with the study team once you decide to stop.

**Are there other reasons why I might stop being in the study?**

Yes. The study doctor may take you off the study if:

- The study is stopped by the National Cancer Institute (NCI), Institutional Review Board (IRB), or study sponsor (the Alliance). The study sponsor is the organization who oversees the study.

This study is conducted by the Alliance for Clinical Trials in Oncology, a national clinical research group supported by the National Cancer Institute. The Alliance is made up of cancer doctors, health professionals, and laboratory researchers, whose goal is to develop better treatments for cancer, to prevent cancer, to reduce side effects from cancer, and to improve the quality of life of cancer patients.

**It is important that you understand the information in the informed consent before making your decision.** Please read, or have someone read to you, the rest of this document. If there is anything you don’t understand, be sure to ask your study staff or nurse.

**What is the purpose of this study?**

Studies have shown that cancer patients may be at high risk for financial problems because of the cost of treatment. These financial problems can be stressful and sometimes might cause patients to avoid or refuse treatment. The purpose of this study is to measure how often financial problems happen in patients with your type of cancer, using questionnaires that collect information about finances and quality of life. In order to get a full picture of the financial impact of chronic lymphocytic leukemia and multiple myeloma, we also want to collect information from your medical records.

You have been asked to participate in this study because you have chronic lymphocytic leukemia or multiple myeloma. This research does not involve medical treatment. Our findings will hopefully help us to better understand the financial impact of cancer and come up with ways to help patients avoid financial problems during treatment.

There will be about 500 people taking part in this study.

**What are the study groups?**

In this study, all study participants will follow the same procedure.

**What exams, tests and procedures are involved in this study?**

If you agree to take part in the study, you will be asked to complete a telephone survey within 8 weeks after joining the study. You will be asked to provide your name, and telephone number so that study team survey staff from the University of North Carolina can call you to conduct the survey. We think it will take about one hour to complete the telephone survey. At the end of the survey, you will be asked for your address so that they can mail a gift card to you. You may refuse to receive the gift if you do not want to provide your address.

We will obtain the following information directly from you through the survey. Topics covered in the survey will include:

- Information about your health insurance, your current work status and some limited information about your household's finances and costs (for example, your monthly income and assets, household debt, and insurance) in the past year;
- Information about whether you have had difficulties paying medical bills and non- medical expenses, your financial worries, your out of pocket costs, any advice that you sought regarding financial and non-financial support;
- Information about whether you have asked for help with medical treatment for cancer (for example, getting help to travel to your doctor's office or access to low cost or free prescription drugs required to treat your cancer);
- Information about your physical and emotional health; and
- Some questions to measure your “financial literacy,” which will involve solving some basic math problems.

We will obtain the following information from your medical record:

- Basic information about you (for example, age, date of birth, gender and race), your insurance status, general health, presence of any other health conditions, and your cancer (for example, diagnosis, date of diagnoses, names of prescription drug treatments you have received or will receive).

**What risks can I expect from taking part in this study?**

**General risks**

There are very few risks to you. If you choose to take part in this study,

- You may feel uncomfortable being asked about your physical and emotional health and your finances.
- There is a small risk that your study information could become known to someone who is not involved in performing or monitoring this study. However, we make every effort to protect your privacy.

**What are my responsibilities in this study?**

If you choose to take part in this study you will need to:

- Complete the telephone survey within 8 weeks after joining the study.

**What are the costs of taking part in this study?**

There are no costs to you for taking part in this study.

After completing the survey, you will be mailed a gift card of $20, in appreciation of your time. The University of North Carolina study team survey staff who is conducting the survey will ask for your mailing address and mail the gift card to you. You may refuse to receive the gift if you do not want to provide your address.

**Who will see my medical information?**

Your privacy is very important to us. The study doctors will make every effort to protect it. The study doctors have a privacy permit to help protect your records if there is a court case.

However, some of your medical information may be given out if required by law. If this should happen, the study doctors will do their best to make sure that any information that goes out to others will not identify who you are.

Some of your health information, such as details of your cancer, your physical and emotional health, your work, health insurance and household finances will be kept by the study sponsor in a central research database. However, your name and contact information will not be put in the database. If information from this study is published or presented at scientific meetings, your name and other personal information will not be used.

To help make sure your information remains private, study staff will use a secure electronic submission program (Medidata Rave) approved by the National Cancer Institute (NCI) to send us the information from your medical record. All data collected during this study will be kept in a password protected, secure, firewalled (blocks unauthorized access) database at the Alliance. A minimum number of authorized staff will be given access to these data. Your personal information including your name, address, and telephone number will not be shared with investigators who are analyzing the data or reporting results.

There are organizations that may look at or receive copies of some of the information in your study records. Your health information in the research database also may be shared with these organizations. They must keep your information private, unless required by law to give it to another group.

- The Alliance for Clinical Trials in Oncology
- The NCI Central IRB, which is a group of people who review the research with the goal of protecting the people who take part in the study.
- The NCI and the groups it works with to review research.

In addition to storing data in the study database, data from studies that are publicaly funded may also be shared broadly for future research with protections for your privacy. The goal of this data sharing is to make more research possible that may improve people’s health. Your study records may be stored and shared for future use in public databases. However, your name and other personal information will not be used.

Some types of future research may include looking at your information and information from other patients to see who had side effects across many studies or comparing new study data with older study data. However, right now we don’t know what research may be done in the future using your information. This means that:

- You will not be asked if you agree to take part in the specific future research studies using your health information.
- You and your study doctor will not be told when or what type of research will be done.
- You will not get reports or other information about any research that is done using your information.

**Where can I get more information?**

You may visit the NCI web site at <http://cancer.gov/> for more information about studies or general information about cancer. You may also call the NCI Cancer Information Service to get the same information at: 1-800-4-CANCER (1-800-422-6237).

A description of this clinical trial will be available on [http://www.ClinicalTrials.gov,](http://www.ClinicalTrials.gov/) as required by U.S. Law. This Web site will not include information that can identify you. At most, the Web site will include a summary of the results. You can search this Web site at any time.

You can talk to the study staff about any questions or concerns you have about this study. Contact the study staff (*insert name of study staff*) at (*insert telephone number, and email address if appropriate*).

For questions about your rights while in this study, call the (*insert name of organization or center*) Institutional Review Board at (*insert telephone number*).

**My signature agreeing to take part in the study**

I have read this consent form or had it read to me. I have discussed it with the study doctor and my questions have been answered. I will be given a signed and dated copy of this form. I agree to take part in the study.

**Participant’s signature**

Date of signature

**Signature of person(s) conducting the informed consent discussion**

Date of signature

**APPENDIX I	PRACTICE SURVEY**

**OVERVIEW**

| **DOMAIN** | **# ITEMS** | **REFERENCE PERIOD** | **TYPE(S) OF RESPONSE** |
| --- | --- | --- | --- |
| Practice characteristics | 4 | Now | Yes, No; Categorical, Fill in |
| Clinical practice characteristics | 2 | Now | Categorical |
| Health information technology | 6 | Now | Yes, No; Categorical |
| Cancer care delivery systems | 6 | Now | Categorical |
| Patient centered Financial navigation, Transportation and Pyschosocial support services | 40 | Now and future | Yes, No; Categorical, Fill in |

**Practice Survey**

The goal of this survey is to assess the patient centered resources NCORP Community and Minority Underserved Sites offer to their patients and their families undergoing cancer diagnosis and treatment. This assessment will provide useful information for the development of interventions to improve patient access to effective and affordable cancer treatment. In the survey, we will use the term “practice” to refer to a clinic, cancer center, physician practice, or other setting where patients with cancer are enrolled in a CCDR study. Like the CCDR Landscape study, your responses should refer to a single CTEP ID where patients that you will enroll in this study have received some or all of their care. If your practice encompasses several sites with different CTEP IDs, you should complete a separate survey for each.

Some CTEP IDs cover multiple physical practice locations. If practice characteristics differ by location, please base your answers on the location where the largest number of patients are treated.

**Section 1. Contact Information**

NCORP Name:

First Name:

Last Name:

Job Title:

Phone Number:

E-mail:

Practice CTEP ID:

Practice Name Associated with CTEP ID:

**Section 2. Practice Characteristics**

1. Does your practice include:
   a. Outpatient clinic in or on a hospital campus *(check one)* ◻Yes	◻No

*(if yes)*, how many different locations?

b. Free-standing outpatient clinic or Private/Group Practice *(check one)* ◻Yes	◻No

*(if yes)*, how many different locations?

c. Inpatient services *(check one)* ◻Yes	◻No

*(if yes)*, how many different locations?

2. Does your practice self-identify as a safety net hospital? *(check one)* ◻Yes	◻No
3. Is your practice affiliated with a designated critical access hospital? *(check one)* ◻Yes	◻No
4. Which of these bests describe the ownership of your practice? *(check one)*

- Independently owned (i.e. single hospital or small regional network (up to three hospitals) or an independent clinic/physician practice)
- Hospital, clinic or physician practice (i.e. owned by a large, regional multi-state health system that **does** include a health plan)
- Hospital, clinic or physician practice (i.e. owned by a large, regional multi-state health system that **does not** include a health plan)
- HMO/Payer owned
- Publicly owned (e.g. state, county, city)
- University owned
- Other

*(if other)*, specify:

**Section 3. Clinical Practice Characteristics**

5. Please provide the estimated number of oncology providers at your practice *(if none, report 0)*

All oncology focused physicians:

All nurse practitioners/physician assistants specializing in the treatment of cancer:

Total number of physicians specializing in the treatment of blood cancers:

Of this total, the number of physicians specializing in the treatment of leukemia:

Of this total, the number of physicians specializing in the treatment of myeloma:

The following questions ask about resources available directly through your practice or through another practice, affiliated hospital or referral service.

6. Do the oncology patients at your practice have access to the following provider?
   a. Psychiatrist *(check one)*
      - Yes, routinely available at the location where they receive cancer treatment or follow-up
      - Not available at that location, but routinely accessed through a clinic or hospital that is affiliated with the same health care system as our oncology practice
      - Not available at our affiliates, but we routinely refer patients to specific outside practices or providers for this service (i.e. local or national organizations that provide cancer services)
      - No routine referral – arrangements are made on a case-by-case basis
      - Not available
   b. Psychologist *(check one)*
      - Yes, routinely available at the location where they receive cancer treatment or follow-up
      - Not available at that location, but routinely accessed through a clinic or hospital that is affiliated with the same health care system as our oncology practice
      - Not available at our affiliates, but we routinely refer patients to specific outside practices or providers for this service (i.e. local or national organizations that provide cancer services)
      - No routine referral – arrangements are made on a case-by-case basis
      - Not available
   c. Neuropsychologist *(check one)*
      - Yes, routinely available at the location where they receive cancer treatment or follow-up
      - Not available at that location, but routinely accessed through a clinic or hospital that is affiliated with the same health care system as our oncology practice
      - Not available at our affiliates, but we routinely refer patients to specific outside practices or providers for this service (i.e. local or national organizations that provide cancer services)
      - No routine referral – arrangements are made on a case-by-case basis
      - Not available
   d. Registered Nurse *(check one)*
      - Yes, routinely available at the location where they receive cancer treatment or follow-up
      - Not available at that location, but routinely accessed through a clinic or hospital that is affiliated with the same health care system as our oncology practice
      - Not available at our affiliates, but we routinely refer patients to specific outside practices or providers for this service (i.e. local or national organizations that provide cancer services)
      - No routine referral – arrangements are made on a case-by-case basis
      - Not available
   e. Rehabilitative medicine *(check one)*
      - Yes, routinely available at the location where they receive cancer treatment or follow-up
      - Not available at that location, but routinely accessed through a clinic or hospital that is affiliated with the same health care system as our oncology practice
      - Not available at our affiliates, but we routinely refer patients to specific outside practices or providers for this service (i.e. local or national organizations that provide cancer services)
      - No routine referral – arrangements are made on a case-by-case basis
      - Not available
   f. Integrative health specialist *(check one)*
      - Yes, routinely available at the location where they receive cancer treatment or follow-up
      - Not available at that location, but routinely accessed through a clinic or hospital that is affiliated with the same health care system as our oncology practice
      - Not available at our affiliates, but we routinely refer patients to specific outside practices or providers for this service (i.e. local or national organizations that provide cancer services)
      - No routine referral – arrangements are made on a case-by-case basis
      - Not available
   g. Clinical pharmacist (medication therapy management) *(check one)*
      - Yes, routinely available at the location where they receive cancer treatment or follow-up
      - Not available at that location, but routinely accessed through a clinic or hospital that is affiliated with the same health care system as our oncology practice
      - Not available at our affiliates, but we routinely refer patients to specific outside practices or providers for this service (i.e. local or national organizations that provide cancer services)
      - No routine referral – arrangements are made on a case-by-case basis
      - Not available
   h. Social worker *(check one)*
      - Yes, routinely available at the location where they receive cancer treatment or follow-up
      - Not available at that location, but routinely accessed through a clinic or hospital that is affiliated with the same health care system as our oncology practice
      - Not available at our affiliates, but we routinely refer patients to specific outside practices or providers for this service (i.e. local or national organizations that provide cancer services)
      - No routine referral – arrangements are made on a case-by-case basis
      - Not available
   i. Patient navigator (including nurse navigator and social worker navigator) *(check one)*
      - Yes, routinely available at the location where they receive cancer treatment or follow-up
      - Not available at that location, but routinely accessed through a clinic or hospital that is affiliated with the same health care system as our oncology practice
      - Not available at our affiliates, but we routinely refer patients to specific outside practices or providers for this service (i.e. local or national organizations that provide cancer services)
      - No routine referral – arrangements are made on a case-by-case basis
      - Not available

**Section 4. Health Information Technology**

7. Does your practice have an outpatient Electronic Health Record (EHR) in which cancer patient data are recorded? *(check one)* ◻Yes	◻No
8. Does your practice have a patient portal? *(check one)* ◻Yes	◻No

**Communication with Patients**

9. Which of the following technologies does your practice routinely use to communicate with oncology patients? (*if a-e are all answered “no”, go to question 11*)
   a. E-mail *(check one)* ◻Yes	◻No
   b. Text *(check one)* ◻Yes	◻No
   c. Mobile apps *(check one)* ◻Yes	◻No
   d. Patient portal *(check one)* ◻Yes	◻No
   e. Other *(check one)* ◻Yes	◻No

*(if yes)*, specify:

10. What information does your practice communicate to patients using these technologies?
    a. Treatment (lab or test results, new medications, follow-up between visits) *(check one)*

◻Yes	◻No

b. Administrative (appointment reminders or scheduling, medication refills) *(check one)*

◻Yes	◻No

c. Financial/billing *(check one)* ◻Yes	◻No
d. Patient education *(check one)* ◻Yes	◻No
e. Opportunities to volunteer for research studies or patient survey *(check one)* ◻Yes	◻No
f. Other *(check one)* ◻Yes	◻No

*(if yes)*, specify:

**Other Capabilities**

11. Does your practice routinely document the linguistic needs of patients? *(check one)* ◻Yes	◻No
    a. *(If yes)*, are the linguistic needs documented in a searchable discrete field (e.g. drop down options or radio buttons) in the EHR? *(check one)* ◻Yes	◻No
12. Can your practice access the complete medical care charges for patients who are enrolled in your research projects? *(check one)* ◻Yes	◻No
    a. *(If yes)*, can this information be accessed through electronic data systems? *(check one)*

◻Yes	◻No

**Section 5. Cancer Care Delivery Systems**

**Oral Chemotherapy Management**

For the following questions, “oral chemotherapy” includes traditional oral cytotoxic agents (e.g., chlorambucil), molecularly targeted agents (e.g., imatinib, mesylate), and hormonal agents (e.g., tamoxifen).

13. Which of the following best describes oral chemotherapy management at your practice? *(check one)*

- Patients are managed by providers (MD, PA, NP, RN, PharmD) during scheduled office visits, and followed up as needed, if patients contact them with concerns
- Patients are managed by providers (MD, PA, NP, RN, PharmD) during scheduled office visits AND routinely followed up by phone between office visits by a staff member
- Other

*(If other)*, specify:

14. Are patients given explicit instructions on when and how to contact the oncology practice if they encounter problems with oral medication like adherence, side effects or filling prescriptions? *(check one)* ◻Yes	◻No

*(If yes)*, How can they reach you?

i. 24/7 call *(check one)* ◻Yes	◻No
ii. During business hours only by e-mail *(check one)* ◻Yes	◻No
iii. During business hours only by patient portal *(check one)* ◻Yes	◻No
iv. During business hours only by text *(check one)* ◻Yes	◻No
v. Other *(check one)* ◻Yes	◻No

*(If other)*, specify:

15. Does your practice use a dedicated staff member(s) located at your practice for oral chemotherapy pre-authorizations? *(check one)* ◻Yes	◻No
16. Does your practice own or have a contractual relationship with a pharmacy? *(check one)* ◻Yes

◻No

a. *(If yes)*, What services does the pharmacy provide to patients?

i. Dispensing medication *(check one)* ◻Yes	◻No
ii. Providing mail order medication *(check one)* ◻Yes	◻No
iii. Counseling patients on medication use *(check one)* ◻Yes	◻No
iv. Counseling patients on financial assistance programs *(check one)* ◻Yes	◻No
v. Other *(check one)* ◻Yes	◻No

*(If other)*, specify:

**Patient Reported Health and Financial Literacy**

17. Does your practice routinely screen patients for health literacy? *(check one)* ◻Yes	◻No
    a. *(if yes)*, do you use a standardized screening tool? ◻Yes	◻No

*(if yes)*, please specify the name of the tool used

18. Does your practice routinely screen patients for financial literacy? *(check one)* ◻Yes	◻No
    a. *(if yes)*, do you use a standardized screening tool? ◻Yes	◻No

*(if yes)*, please specify the name of the tool used

**Section 6. Patient-centered financial navigation, transportation and psychosocial support services**

The following questions ask about financial navigation, transportation and psychosocial resources that your practice **provides directly to patients**. If your practice accesses any of these resources through another practice or affiliated hospital, please respond “No” to that question.

**Financial Navigation Services**

*Defining “Financial Navigation”: Financial navigation refers to processes by which patients and their families are aided in affording care after a cancer diagnosis to avoid adverse financial consequences and hardship associated with cancer treatment (e.g., education about and assistance with accessing appropriate financial programs and services).*

**Current Practices: Financial Navigation Services**

19. Does your practice screen patients for their interest in receiving help with financial navigation for their medical care? *(check one)* ◻Yes	◻No
    a. *(if yes),* how is screening implemented in your practice?
i. Electronic health record *(check one)*	◻Yes	◻No
ii. Patient intake form *(check one)*	◻Yes	◻No
iii. Vitals or other form completed by medical professionals *(check one)*	◻Yes	◻No
iv. Distress screening tool *(check on)* ◻Yes	◻No
v. Other *(check one)*	◻Yes	◻No

*(if other)*, specify:

20. What financial navigation services are currently offered at your practice to all cancer patients or their families? *(if a-g are all answered no, skip to question 23)*
    a. Help understanding medical bills and out of pocket costs generated from your practice *(check one)*	◻Yes	◻No
    b. Structuring payment plans for patients late in paying medical bills generated from your practice *(check one)*	◻Yes	◻No
    c. Referral to local or national charities to help pay for outstanding medical debts *(check one)*

◻Yes	◻No

d. Referral to local or national charities to help pay for co-pays, cost of medical care, and/or cost of prescriptions *(check one)*	◻Yes	◻No
e. Referral to pharmaceutical manufacturer to help cover drug costs *(check one)*	◻Yes	◻No
f. Referral to state and local insurance benefit support (sign up for Medicaid coverage, for ACA exchange coverage, for individual insurance plans) *(check one)* ◻Yes	◻No
g. Other *(check one)*	◻Yes	◻No

(If other), specify:

21. How are these financial navigation services communicated to your patients and their caregivers?
    a. Posted written notices in waiting rooms and individual exam rooms *(check one)*

◻Yes	◻No

b. Written materials available in waiting rooms and individual exam rooms *(check one)*

◻Yes	◻No

c. Orally offered by practice providers (nurses, NPs, physicians) *(check one)*	◻Yes	◻No
d. On the hospital website *(check one)*	◻Yes	◻No
e. Via e-mail correspondence *(check one)*	◻Yes	◻No
f. Other *(check one)*	◻Yes	◻No

*(if other)*, specify:

22. Which types of providers are involved in providing referrals or services for the financial navigation services listed above?
    a. Financial navigator who serves cancer patients *(check one)*	◻Yes	◻No
    b. Financial navigator who is not dedicated to cancer patients (e.g., serves entire clinic) *(check one)*	◻Yes	◻No
    c. Social worker *(check one)*	◻Yes	◻No
    d. Billing staff *(check one)*	◻Yes	◻No
    e. Doctors/nurses *(check one)*	◻Yes	◻No
    f. Health educators *(check one)*	◻Yes	◻No
    g. Case managers *(check one)*	◻Yes	◻No
    h. Pharmacy staff *(check one)*	◻Yes	◻No
    i. Outside services (e.g., patient support groups) *(check one)*	◻Yes	◻No
    j. Other *(check one)*	◻Yes	◻No

*(if other)*, specify:

The next question asks about the **potential challenges experienced by your practice** with regards to financial navigation services.

23. In your practice, do any of the following issues make it difficult to provide financial navigation services to patients?
    a. Lack of staff awareness about financial navigation services *(check one)*	◻Yes	◻No

*(If yes)*, how much *(check one)*:

◻A little bit

◻Somewhat

◻Quite a bit

◻Very much

b. Lack of availability of financial navigation services *(check one)*	◻Yes	◻No

*(If yes)*, how much *(check one)*:

◻A little bit

◻Somewhat

◻Quite a bit

◻Very much

c. Lack of staff time or capacity to arrange these financial navigation services *(check one)*	◻Yes

◻No

*(If yes)*, how much *(check one)*:

◻A little bit

◻Somewhat

◻Quite a bit

◻Very much

**Anticipated Changes to Financial Navigation Services Offered by your Practice to Cancer Patients**

24. Are there currently any plans to enhance the financial navigation services or resources available to patients at your practice? *(check one)*	◻Yes	◻No (*if no, skip to question 28)*
25. When do you anticipate these changes to current services to begin? *(check one)*

- Currently in progress
- Within 1-3 months
- Within 4-6 months
- Within 7-12 months
- Within the next 12 months
- Greater than 12 months

26. What is prompting the implementation of the enhancement(s)?

- Patient population has expressed financial difficulties *(check one)*	◻Yes	◻No
- Patient population has requested financial assistance *(check one)*	◻Yes	◻No
- Medicare payers require financial navigation *(check one)*	◻Yes	◻No
- Financial navigation is practice norm *(check one)*	◻Yes	◻No
- Other *(check one)*	◻Yes	◻No

*(if other)*, specify:

27. Please tell us about the nature of the enhancement(s).
28. Are there currently any plans to reduce the financial navigation services available to patients at your practice? *(check one)* ◻Yes ◻No *(if no, skip to question 32)*
29. When do you anticipate these reductions will begin? *(check one)*

- Currently in progress
- Within 1-3 months
- Within 4-6 months
- Within 7-12 months
- Within the next 12 months
- Greater than 12 months

30. What is prompting these reductions in services?
    a. Insufficient staffing to keep up this effort *(check one)*	◻Yes	◻No
    b. Closure of outside agencies or programs *(check one)*	◻Yes	◻No
    c. Reduction in coverage for services *(check one)*	◻Yes	◻No
    d. End of grant-funded programs *(check one)*	◻Yes	◻No
    e. Service is under-utilized *(check one)*	◻Yes	◻No
    f. Other *(check one)*	◻Yes	◻No

*(if other)*, specify:

31. Please tell us about the nature of the reduction(s) to services.

**Transportation Services**

The next set of questions are focused on **transportation services** that your practice **provides directly to patients**.

**Current Practices: Transportation Services**

32. Does your practice screen patients for interest in help getting to needed office visits? *(check one)*

◻Yes	◻No

a. *(if yes)*, how is screening is implemented in your practice?

i. Electronic health record *(check one)* ◻Yes	◻No
ii. Patient intake form *(check one)* ◻Yes	◻No
iii. Vitals or other form completed by medical professionals *(check one)*	◻Yes	◻No
iv. Other *(check one)*	◻Yes	◻No

*(if other)*, specify:

33. What transportation services are currently offered at your practice to all cancer patients or their families? *(if a-e are all answered no, skip to question 36)*
    a. Transportation services to and from your practice (i.e. volunteer drivers, transportation rentals, shuttle buses) *(check one)*	◻Yes	◻No
    b. Transportation vouchers (i.e. bus, taxi, Uber, Lyft, etc.) *(check one)*	◻Yes	◻No
    c. Gas vouchers, gas money or parking vouchers *(check one)*	◻Yes	◻No
    d. Special subsidized transportation for patients with limited mobility *(check one)*	◻Yes

◻No

e. Other *(check one)*	◻Yes	◻No

*(if other)*, specify:

34. How are these transportation services communicated to your patients and their caregivers?
    a. Posted written notices in waiting rooms and individual exam rooms *(check one)*

◻Yes	◻No

b. Written materials available in waiting rooms and individual exam rooms *(check one)*

◻Yes	◻No

c. Orally offered by practice providers (nurses, NPs, physicians) *(check one)*	◻Yes	◻No
d. On the hospital website *(check one)*	◻Yes	◻No
e. Via e-mail correspondence *(check one)*	◻Yes	◻No
f. Other *(check one)*	◻Yes	◻No

*(if other)*, specify:

35. Which types of providers are involved in providing referrals or services for the transportation services listed above?
    a. Social worker *(check one)* ◻Yes	◻No
    b. Psychologist *(check one)* ◻Yes	◻No
    c. Doctors/nurses *(check one)* ◻Yes	◻No
    d. Health Educators *(check one)* ◻Yes	◻No
    e. Nutritionists *(check one)* ◻Yes	◻No
    f. Case Managers *(check one)* ◻Yes	◻No
    g. Other *(check one)* ◻Yes	◻No

*(if other)*, specify:

The next question asks about the **potential challenges experienced by your practice** with regards to transportation services.

36. In your practice, do any of the following issues make it difficult to provide transportation services to patients?
    a. Lack of staff awareness about transportation services *(check one)*	◻Yes	◻No

*(If yes)*, how much *(check one)*:

◻A little bit

◻Somewhat

◻Quite a bit

◻Very much

b. Lack of availability of transportation services *(check one)*	◻Yes	◻No

*(If yes)*, how much *(check one)*:

◻A little bit

◻Somewhat

◻Quite a bit

◻Very much

c. Lack of staff time or capacity to arrange these transportation services *(check one)*	◻Yes	◻No

*(If yes)*, how much *(check one)*:

◻A little bit

◻Somewhat

◻Quite a bit

◻Very much

d. Coordination with transportation service providers (for example, if service is provided off site or through a company) *(check one)* ◻Yes	◻No

*(If yes)*, how much *(check one)*:

◻A little bit

◻Somewhat

◻Quite a bit

◻Very much

e. Low uptake of these transportation services by patients due to cost or other factors *(check one) (check one)* ◻Yes	◻No

*(If yes)*, how much *(check one)*:

◻A little bit

◻Somewhat

◻Quite a bit

◻Very much

f. The responsibility of scheduling or following-up on these services falls on the patient *(check one)* ◻Yes	◻No

*(If yes)*, how much *(check one)*:

◻A little bit

◻Somewhat

◻Quite a bit

◻Very much

g. The transportation distance is too far (i.e. patient resides in a rural location) *(check one)* ◻Yes

◻No

*(If yes)*, how much *(check one)*:

◻A little bit

◻Somewhat

◻Quite a bit

◻Very much

**Anticipated Changes to Transportation Services Offered by your Practice to Cancer Patients**

37. Are there currently any plans to enhance the transportation services or resources available to patients at your practice? *(check one)*	◻Yes	◻No (*if no, skip to question 41)*
38. When do you anticipate these changes to current services to begin? *(check one)*

- Currently in progress
- Within 1-3 months
- Within 4-6 months
- Within 7-12 months
- Within the next 12 months
- Greater than 12 months

39. What is prompting the implementation of the enhancement(s)?

- Patient population has expressed transportation difficulties *(check one)*	◻Yes	◻No
- Patient population has requested transportation assistance *(check one)*	◻Yes	◻No
- Transportation navigation is practice norm *(check one)*	◻Yes	◻No
- Other *(check one)*	◻Yes	◻No

*(if other)*, specify:

40. Please tell us about the nature of the enhancement(s).
41. Are there currently any plans to reduce the transportation services available to patients at your practice? *(check one)*	◻Yes	◻No *(if no, skip to question 45)*
42. When do you anticipate these reductions will begin? *(check one)*

- Currently in progress
- Within 1-3 months
- Within 4-6 months
- Within 7-12 months
- Within the next 12 months
- Greater than 12 months

43. What is prompting these reductions in services?
    a. Staff Layoff *(check one)*	◻Yes	◻No
    b. Insufficient staffing to keep up this effort *(check one)*	◻Yes	◻No
    c. Closure of outside agencies or programs *(check one)*	◻Yes	◻No
    d. Reduction in coverage for services *(check one)*	◻Yes	◻No
    e. End of grant-funded programs *(check one)*	◻Yes	◻No
    f. Service is under-utilized *(check one)*	◻Yes	◻No
    g. Difficulty in maintaining or staffing the program *(check one)*	◻Yes	◻No
    h. Other	*(check one)*	◻Yes	◻No
44. Please tell us about the nature of the reduction(s) to services.

**Psychosocial Services**

The next set of questions ask about psychosocial resources your practice provides directly to patients. The term “psychosocial” encompasses the patient’s social, emotional and spiritual well-being.

**Current Practices: Psychosocial Services**

45. Does your practice screen patients for interest in psychosocial services or resources?

*(check one)* ◻Yes	◻No

a. *(if yes),* how is psychosocial screening implemented in your practice?

i. Electronic health record *(check one)*	◻Yes	◻No
ii. Patient intake form *(check one)*	◻Yes	◻No
iii. Vitals or other form completed by medical professionals *(check one)*	◻Yes	◻No
iv. Distress screening tool *(check one)* ◻Yes	◻No
v. Other *(check one)*	◻Yes	◻No

*(if other)*, specify:

46. What psychosocial services or resources are currently offered at your practice to all cancer patients and their families? *(If a-i are all answered no, skip to question 49)*
    a. Assistance with **housing and housing costs**, including rent subsidy, payment for utilities, accessibility and transitions, *(check one)*	◻Yes	◻No
    b. Assistance with **food and nutrition,** including nutritional counseling, access to healthy food, home delivery of meals. *(check one)*	◻Yes	◻No
    c. Individual counseling *(check one)*	◻Yes	◻No
    d. Group counseling *(check one)* ◻Yes	◻No
    e. Family counseling *(check one)*	◻Yes	◻No
    f. Mutual help support groups *(check one)*	◻Yes	◻No
    g. Meditation *(check one)* ◻Yes	◻No
    h. Peer Support or Patient to Patient Programs *(check one)*	◻Yes	◻No
    i. Other *(check one)*	◻Yes	◻No

*(If other),* specify:

47. How are these psychosocial services communicated to your patients and their caregivers?
    a. Posted written notices in waiting rooms and individual exam rooms *(check one)*

◻Yes	◻No

b. Written materials available in waiting rooms and individual exam rooms *(check one)*

◻Yes	◻No

c. Orally offered by practice providers (nurses, NPs, physicians) *(check one)*	◻Yes	◻No
d. On the hospital website *(check one)*	◻Yes	◻No
e. Via e-mail correspondence *(check one)*	◻Yes	◻No
f. Other *(check one)*	◻Yes	◻No

*(if other)*, specify:

48. Which types of providers are involved in providing referrals or services for the psychosocial services listed above?
    a. Social worker *(check one)* ◻Yes	◻No
    b. Psychologist *(check one)* ◻Yes	◻No
    c. Doctors/nurses *(check one)* ◻Yes	◻No
    d. Health Educators *(check one)* ◻Yes	◻No
    e. Nutritionists *(check one)* ◻Yes	◻No
    f. Case Managers *(check one)* ◻Yes	◻No
    g. Other *(check one)* ◻Yes	◻No

*(if other)*, specify:

The next question asks about the **potential challenges experienced by your practice** with regards to psychosocial services.

49. In your practice, do any of the following issues make it difficult to provide psychosocial services to patients?
    a. Lack of staff awareness about psychosocial services *(check one)*	◻Yes	◻No

*(If yes)*, how much *(check one)*:

◻A little bit

◻Somewhat

◻Quite a bit

◻Very much

b. Lack of availability of psychosocial services *(check one)*	◻Yes	◻No

*(If yes)*, how much *(check one)*:

◻A little bit

◻Somewhat

◻Quite a bit

◻Very much

c. Lack of staff time or capacity to arrange these psychosocial services *(check one)*	◻Yes	◻No

*(If yes)*, how much *(check one)*:

◻A little bit

◻Somewhat

◻Quite a bit

◻Very much

d. Low uptake of these psychosocial services by patients due to cost or other factors *(check one)*

◻Yes	◻No

*(If yes)*, how much *(check one)*:

◻A little bit

◻Somewhat

◻Quite a bit

◻Very much

**Anticipated Changes to Psychosocial Services Offered by your Practice to Cancer Patients**

50. Are there currently plans to enhance the psychosocial services or resources available to patients at your practice? *(check one)*	◻Yes	◻No (*if no, skip to question 54)*
51. When do you anticipate these changes to current services will begin? *(check one)*

- Currently in progress
- Within 1-3 months
- Within 4-6 months
- Within 7-12 months
- Within the next 12 months
- Greater than 12 months

52. What is prompting the implementation of the enhancement(s)?

- Patient population has expressed psychosocial difficulties *(check one)*	◻Yes	◻No
- Patient population has requested psychosocial program *(check one)*	◻Yes	◻No
- Psychosocial program navigation is a practice norm *(check one)*	◻Yes	◻No
- Other *(check one)*	◻Yes	◻No

*(if other)*, specify:

53. Please tell us about the nature of the enhancement(s).
54. Are there currently any plans to reduce the psychosocial services available to patients at your practice? *(check one)*	◻Yes	◻No *(if no, skip to question 58)*
55. When do you anticipate these reductions will begin? *(check one)*

- Currently in progress
- Within 1-3 months
- Within 4-6 months
- Within 7-12 months
- Within the next 12 months
- Greater than 12 months

56. What is prompting these reductions in services?
    a. Insufficient staffing to keep up this effort *(check one)*	◻Yes	◻No
    b. Closure of outside agencies or programs *(check one)*	◻Yes	◻No
    c. Reduction in coverage for services *(check one)*	◻Yes	◻No
    d. End of grant-funded program *(check one)*	◻Yes	◻No
    e. Service is under-utilized *(check one)*	◻Yes	◻No
    f. Difficulty in maintaining or staffing the program *(check one)*	◻Yes	◻No
    g. Other *(check one)*	◻Yes	◻No

*(if other)*, specify:

57. Please tell us about the nature of the reduction(s) to services.
58. If you identified accessing **any resources** through another practice or affiliated hospital, please include the name of the practice or affiliated hospital along with their CTEP ID (if possible): [ALLOW FOR MULTIPLE ENTRIES]

Name of practice or parent hospital:

CTEP ID:

**Thank you for taking the time to complete this survey. Your participation will provide valuable information on practice infrastructure and capacities which will help us develop patient-focused interventions.**

**APPENDIX II	PATIENT SURVEY**

Contents

**STARTING PHONE INTERVIEW WITH PARTICIPANT PART 1: FINANCIAL DIFFICULTY**

**PART 2: PATIENT REPORT OF SEEKING AND RECEIVING FINANCIAL SUPPORT PART 3: PATIENT SOCIODEMOGRAPHIC INDICATORS**

**PART 4: HEALTH AND WELL-BEING CLOSING TO INTERVIEW**

**STARTING PHONE INTERVIEW WITH PARTICIPANT**

1. **Call patient:**
   a. Someone answers the phone 🡪 Go to 2
   b. Goes to voicemail: On the 1^st^ and 4^th^ call attempt, leave message: “Hello, my name is (FULL NAME) and I am calling from the University of North Carolina about a health study. We are trying to get in touch with [PARTICIPANT FIRST AND LAST NAME]. We’ll try you back again later, or you can call us at 919-962- xxxx.”
      - Indicate contact attempt in RAVE Patient Contact Log as *Call not answered; message left on voicemail*
      - Make up to 8 attempts to reach participant the first time, until marking patient as *No further contact* in RAVE Patient Contact Log.
   c. No response, and no voicemail reached after 12 rings
i. Indicate contact attempt in RAVE Patient Contact Log as *Call not answered; unable to leave message*
ii. Make up to 8 attempts to reach participant the first time, until marking patient as *No further contact* in RAVE Patient Contact Log.
2. **Someone answers phone**. Say “Hello, my name is [FULL NAME]. I am calling from the University of North Carolina for a health study. May I please speak with [PARTICIPANT FIRST AND LAST NAME]?
   a. They indicate that they are the participant. 🡪 Go to 3
   b. They indicate that the participant is not available.
i. “OK, thank you for letting me know. When would be a good time for me to speak with [him/her]?”
   1. Schedule date/time to call back; or ask about participant’s general availability over the next week.
   2. If person asks if they can take a message for the participant, say “Thank you. Please let him/her know that I am calling from the University of North Carolina about a health study. We’ll try this number back again at another time.”
   3. Indicate contact attempt in RAVE Patient Contact Log as

*Participant not available.*

c. They indicate the participant is no longer at this number.

i. “Thank you for letting me know. Do you know [his/her] new number?”
   1. Indicate contact attempt in RAVE Patient Contact Log as

*Participant no longer at this number.*

2. If new number is known, verify updated information with lead study coordinator.
3. If number is unknown, contact lead study coordinator to discuss next steps.
d. They indicate there is no one by the participant’s name at this number

i. “Let me just confirm that I dialed correctly. Did I reach you at ###-###- ####?”
   1. Correct number called: “I’m sorry. I must have the wrong number in my records. Thank you for your time.”
      a. Indicate contact attempt in RAVE Patient Contact Log as

*No one by participant’s name at phone number*

b. Contact lead study coordinator to discuss next steps.
2. Incorrect number: “I’m sorry. I must have misdialed. Thank you for your time.”
   a. Re-dial correct number.
e. They hang up or refuse to talk

i. Indicate contact attempt in RAVE Patient Contact Log as *Hang up or participant refuses to come to phone*
ii. If response appears to be indicative of a soft refusal, make one additional attempt to reach participant before marking patient as *No further contact* in RAVE Patient Contact Log.
    f. They indicate the participant has died
i. “I am so sorry to hear that. Thank you for your time.”
ii. Indicate contact attempt in RAVE Patient Contact Log as *Participant Died*
3. **Participant is at the phone:** “Hi [Name]. I am calling about the study – Assessing Financial Difficulty in Patients with Blood Cancers, that you enrolled in through the [CLINIC NAME]. The purpose of the study is to help us to better understand patients’ financial experiences related to their cancer treatment and to identify ways to improve patient access to resources at their treatment sites. I would like to have you complete the study survey over the phone today. It will take a little over an hour. Is this a good time?
   a. Yes, it’s a good time 🡪 Go to 4
   b. Not a good time, but participant can reschedule: “I'd be happy to call you back at a later time. When would be a better time for me to speak with you?”
i. Schedule date/time to call back; or ask about participant’s general

availability over the next week.

ii. Indicate contact attempt in RAVE Patient Contact Log as *Survey Not Started. Patient did not have time.*
iii. Make up to 8 attempts to reschedule survey, until marking patient as *No further contact* in RAVE Patient Contact Log.
     c. Participant does not want to participate in the study. “OK. May I ask why you don't want to participate?”
i. Upon 2^nd^ soft refusal or 1st hard refusal: “OK, thank you for your time.”
ii. Indicate contact attempt in RAVE Patient Contact Log as one of the following:
    1. *Survey Not Started. Patient refusal: patient reported physical reason, including fatigue*
    2. *Survey Not Started. Patient refusal: patient reported mental/emotional reason, including stress*
    3. *Survey Not Started. Patient refusal: patient did not give a reason*
    4. *Survey Not Started. Patient refusal: patient did not want to be contacted*
    5. *Survey Not Started. Patient refusal: other, specify*
4. **DOB verification. “**Before I continue, I need to ask you a few questions to confirm that our information is correct.
   a. “Is your date of birth [DOB]?”
i. Yes 🡪 Go to 5
ii. No: “Since we have some incorrect information, we’ll have to check our records and call you back. When would be a good time for me to call back?”
5. **Risk of discomfort and right of refusal**: “There is a chance that some of the questions may make you feel uncomfortable. You have the right to refuse to answer any questions you don’t want to answer and you can stop participating at any time. Is this OK?”
   a. Yes 🡪 Go to 6
   b. No: “OK. May I ask why you don't want to participate?”
i. Upon 2^nd^ soft refusal or 1st hard refusal: “OK, thank you for your time.”
ii. Indicate contact attempt in RAVE Patient Contact Log as one of the following:
    1. *Survey Not Started. Patient refusal: patient reported physical reason, including fatigue*
    2. *Survey Not Started. Patient refusal: patient reported mental/emotional reason, including stress*
    3. *Survey Not Started. Patient refusal: patient did not give a reason*
    4. *Survey Not Started. Patient refusal: patient did not want to be contacted*
    5. *Survey Not Started. Patient refusal: other, specify*
6. **Protection of privacy**: “To protect your privacy, we will assign you a unique identification number. We will store all your answers only with that number and not your name. Only trained project staff will have access to your answers. Is this OK?”
   a. **Yes** 🡪 **Go to 7**
   b. No: “OK. May I ask why you don't want to participate?”
i. Upon 2^nd^ soft refusal or 1st hard refusal: “OK, thank you for your time.”
ii. Indicate contact attempt in RAVE Patient Contact Log as one of the following:
    1. *Survey Not Started. Patient refusal: patient reported physical reason, including fatigue*
    2. *Survey Not Started. Patient refusal: patient reported mental/emotional reason, including stress*
    3. *Survey Not Started. Patient refusal: patient did not give a reason*
    4. *Survey Not Started. Patient refusal: patient did not want to be contacted*
    5. *Survey Not Started. Patient refusal: other, specify*
7. **Questions:** “Do you have any questions before we begin?”
   a. Yes, and questions can be answered 🡪 Answer questions and then begin the Participant Survey
   b. Yes, and questions require more information 🡪 “Thank you – I need some time to look into this; I’m going to discuss your question with our study team and get back to you. Would you like to proceed with the phone interview today or would you rather wait until I can talk with our study team and answer your question?”
   c. No 🡪 Begin Participant Survey

**DURING SURVEY – PARTICIPANT NEEDS TO DISCONTINUE PARTICIPATION**

1. **Patient refuses to continue: “**OK. May I ask why you no longer want to continue?”
   a. Upon 2^nd^ soft refusal or 1st hard refusal: “OK, thank you for your time.”
   b. Indicate contact attempt in RAVE Patient Contact Log as one of the following:
      i. *Survey Incomplete. Patient refusal: patient reported physical reason, including fatigue*
      ii. *Survey Incomplete. Patient refusal: patient reported mental/emotional*

*reason, including stress*

iii. *Survey Incomplete. Patient refusal: patient said did not like content of questions*
iv. *Survey Incomplete. Patient refusal: patient did not give a reason*
v. *Survey Incomplete. Patient refusal: other, specify:*

2. **Patient ran out of time and wants to continue at a later date:** “I'd be happy to call you back at a later time. When would be a better time for me to speak with you?
   a. Schedule date/time to call back; or ask about participant’s general availability over the next week.
   b. Indicate contact attempt in RAVE Patient Contact Log as *Survey Incomplete.*

*Patient ran out of time*

c. Make up to 8 attempts to reach participant to continue survey, until marking patient as *No further contact* in RAVE Patient Contact Log.

3. **Staff needs to discontinue**
   a. Indicate contact attempt in RAVE Patient Contact Log as one of the following:
      i. *Survey Incomplete. Staff unable to administer, specify:*
      ii. *Survey Incomplete. Staff elects to discontinue, specify:*
   b. Contact lead study coordinator to discuss issue causing staff discontinuation..

**Part 1: Financial Difficulty**

The first set of questions are about financial difficulty you might have had because of medical bills. Please think back over the past 12 months -- that would be from now back to [Month] of last year.

**Difficulties paying medical bills**

1. In the past 12 months, did you have problems paying or were unable to pay any medical bills? Include bills for doctors, hospitals, therapists, medication, equipment, nursing home or home. *(check one)*
   - Yes
   - No
2. Do you or anyone in your family currently have medical bills that you are unable to pay at all? *(check one)*
   - Yes
   - No
3. Do you or anyone in your family currently have medical bills that are being paid off over time? This could include medical bills being paid off with a credit card, through personal loans, or bill paying arrangements with hospitals or other providers. The bills can be from earlier years as well as this year. *(check one)*
   - Yes
   - No

**Delays in treatments, foregoing recommended treatment**

The next set of questions are about delays in medical care or skipping medical care because of costs.

4. During the past 12 months, have you or someone in your family delayed medical care because you were worried about the cost (do not include dental care)? *(check one)*
   - Yes
   - No
5. During the past 12 months, was there a time when you or someone in your family needed medical care but didn’t get it because you couldn’t afford it? *(check one)*
   - Yes
   - No
6. During the past 12 months, was there a time when you needed one of the following, but did not get it because you couldn’t afford it?
a. Prescription medication *(check one)*
   - Yes
   - No
b. Mental health care or counseling *(check one)*
   - Yes
   - No
c. Dental care *(check one)*
   - Yes
   - No
d. Eyeglasses *(check one)*
   - Yes
   - No
e. Cancer-related medical care *(check one)*
   - Yes
   - No
f. Non-cancer related medical care *(check one)*
   - Yes
   - No
g. Follow-up care *(check one)*
   - Yes
   - No

**Difficulties covering non-medical expenses**

7. Have you or anyone in your house had to make any of the following financial sacrifices in the past 5 years because of debt related to medical care?
a. Reduced spending on vacation or leisure activities *(check one)*
   - Yes
   - No
b. Reduced spending on purchasing large items (e.g. a car) *(check one)*
   - Yes
   - No
c. Reduced spending on basics (e.g. food and clothing) *(check one)*
   - Yes
   - No
d. Delayed or reduced spending on home improvement *(check one)*
   - Yes
   - No
e. Used savings set aside for other purposes (e.g. retirement, educational funds, family support)

*(check one)*

- Yes
- No

f. Made a change to living situation (e.g. sold, refinanced or moved to a smaller residence) *(check one)*
   - Yes
   - No
g. Other *(check one)*
   - Yes
   - No

*(if yes)*, specify:

**Patient report of financial difficulty**

8. In the last 12 months, has your physical condition or medical treatment caused you financial difficulties? *(check one)*
   - Not at all
   - A little
   - Quite a bit
   - Very much

**Financial worries**

The next set of questions are about financial worries.

9. If you get sicker or have an accident, how worried are you that you will not be able to pay your medical bills? *(check one)*
   - Very worried
   - Somewhat worried
   - Not worried
10. How often in the last 12 months would you say you were worried or stressed about having enough money to pay your rent or mortgage? *(check one)*
    - Always
    - Usually
    - Sometimes
    - Rarely
    - Never
11. How often in the last 12 months would you say you were worried or stressed about having enough money to buy nutritious meals? *(check one)*
    - Always
    - Usually
    - Sometimes
    - Rarely
    - Never
12. How often in the last 12 months would you say you were worried or stressed about having enough money to pay household utilities such as water, gas, and electricity? *(check one)*
    - Always
    - Usually
    - Sometimes
    - Rarely
    - Never
13. Did you, your spouse or significant other ever stay at a job in part because he/she was concerned about losing health insurance for the family? *(check one)*
    - Yes
    - No
    - Does not apply
14. How worried are you that you will lose your current health insurance in the next year? *(check one)*
    - Very worried
    - Somewhat worried
    - Not worried

**Patient Medications**

Now, I’d like to ask you about your current medications.

15. How many different prescription medications are you currently taking? *(if response is “I don’t know,” “Not answered,” or “Not asked” please type the response in the text box provided)*
16. During the past 12 months, were any of the following true for you:
a. You skipped medication doses to save money *(check one)*
   - Yes
   - No

*(If Yes),* Was this a medication for treating: *(check one)*

- Cancer
- Another condition
- Both

b. You took less medicine to save money *(check one)*
   - Yes
   - No

*(If Yes)*, Was this a medication for treating: *(check one)*

- Cancer
- Another condition
- Both

c. You delayed filling a prescription to save money *(check one)*
   - Yes
   - No

*(If Yes)*, Was this a medication for treating: *(check one)*

- Cancer
- Another condition
- Both

d. You asked your doctor for a lower cost medication to save money *(check one)*
   - Yes
   - No

*(If Yes)*, Was this a medication for treating: *(check one)*

- Cancer
- Another condition
- Both

e. You bought prescription drugs from another country to save money *(check one)*
   - Yes
   - No

*(If Yes)*, Was this a medication for treating: *(check one)*

- Cancer
- Another condition
- Both

f. You used alternative therapies to save money *(check one)*
   - Yes
   - No

*(If Yes)*, Was this a substitute treatment for: *(check one)*

- Cancer
- Another condition
- Both

g. You shopped around at pharmacies to get a medication at a lower price *(check one)*
   - Yes
   - No

(If Yes), Was this a medication for treating: *(check one)*

- Cancer
- Another condition
- Both

**Patient reported medical care and out of pocket costs associated with medical care**

The next questions ask about different kinds of financial burden you or your family may have experienced because of your cancer, its treatment, or the lasting effects of that treatment.

17. In the past 12 months, have you or has anyone in your family had to borrow money or go into debt because of your cancer, its treatment or the lasting effects of that treatment? *(check one)*
    - Yes
    - No
18. In the past 12 months, did you or your family file for bankruptcy because of your cancer, its treatment, or the lasting effects of that treatment? *(check one)*
    - Yes
    - No
19. In the past 12 months, have you or your family had to make any other kinds of financial sacrifices because of your cancer, its treatment, or the lasting effects of that treatment? *(check one)*
    - Yes
    - No
20. In the past 12 months, have you ever worried about having to pay large medical bills related to your cancer? *(check one)*
    - Yes
    - No
21. Please think about medical care visits for cancer, its treatment, or the lasting effects of that treatment in the past 12 months. Have you ever been unable to cover your share of those visits? *(check one)*
    - Yes
    - No

**Patient concerns regarding health care navigation, finances, daily life, effects of treatment and psychosocial effects**

Now I’m going to ask you about 15 items that you may have worried about during the past 12 months with multiple myeloma (MM) or chronic lymphocytic leukemia (CLL) and its treatment. For each item, please tell me if it is something you were NOT worried about, were SOMEWHAT worried about, or were VERY worried about during the past 12 months.

With regards to living with multiple myeloma or chronic lymphocytic leukemia, over the past 12 months how much were you worried about…..

22. Understanding all of your treatment options *(check one)*
    - Very worried
    - Somewhat worried
    - Not worried
23. Having access to the best medical care *(check one)*
    - Very worried
    - Somewhat worried
    - Not worried
24. Coordinating all of your doctor appointments *(check one)*
    - Very worried
    - Somewhat worried
    - Not worried
25. The financial cost of your medical treatment *(check one)*
    - Very worried
    - Somewhat worried
    - Not worried
26. Finding financial aid to help pay for medical care *(check one)*
    - Very worried
    - Somewhat worried
    - Not worried
27. The effort needed to manage changes to your financial situation *(check one)*
    - Very worried
    - Somewhat worried
    - Not worried
28. Your disease affecting your professional life *(check one)*
    - Very worried
    - Somewhat worried
    - Not worried
29. Your disease interfering with your leisure activities *(check one)*
    - Very worried
    - Somewhat worried
    - Not worried
30. Your disease limiting your everyday functioning *(check one)*
    - Very worried
    - Somewhat worried
    - Not worried
31. Short term side effects of treatment *(check one)*
    - Very worried
    - Somewhat worried
    - Not worried
32. Long term side effects of treatment *(check one)*
    - Very worried
    - Somewhat worried
    - Not worried
33. Spending too much time in the hospital *(check one)*
    - Very worried
    - Somewhat worried
    - Not worried
34. Possibility of dying from your disease *(check one)*
    - Very worried
    - Somewhat worried
    - Not worried
35. Coping with the emotional demands of your situation *(check one)*
    - Very worried
    - Somewhat worried
    - Not worried
36. Becoming a burden to those who care for you *(check one)*
    - Very worried
    - Somewhat worried
    - Not worried

**Part 2: Patient Report of Seeking and Receiving Financial Support**

**Advice regarding financial or non-financial support**

Now I want to ask you about your experiences with getting financial support to help pay for cancer care.

1. In the past 12 months, have you asked for or received advice on how to pay for any aspect of cancer care? *(check one)*

#### (If yes, go to question 2. If “no,” go to question 3).

- Yes
- No

2. Who did you ask?
1. Family / friends *(check one)*
   - Yes
   - No
2. Doctors / nurses *(check one)*
   - Yes
   - No
3. Social workers *(check one)*
   - Yes
   - No
4. Financial navigators / counselors *(check one)*
   - Yes
   - No
5. Pharmacists *(check one)*
   - Yes
   - No
6. Church *(check one)*
   - Yes
   - No
7. Union *(check one)*
   - Yes
   - No
8. Local community organization *(check one)*
   - Yes
   - No
9. National patient support organization or foundation *(check one)*
   - Yes
   - No
10. Social media groups *(check one)*
    - Yes
    - No
11. Others *(check one)*
    - Yes
    - No

*(if yes)*, specify:

**Formal support for specific purposes**

Now I am going to ask a series of questions about support you may have received from an agency, foundation or organization. For each set of questions, I will ask about a specific type of assistance, when you became aware of this assistance, how this assistance affected your cost, and if anyone helped you apply for support. Later I will ask you about support you have received from family and friends.

3. In the past 12 months have you received any type of financial support from an agency, a foundation, a hospital through charity care or another type of organization? *(check one)*
   - Yes
   - No

*(If yes)*, what did this support help pay for:

a. Prescription medications (e.g., co-pay assistance programs, pharmaceutical manufacturer patient assistance programs, etc.)? *(check one)*
   - Yes
   - No

***(If yes),* answer the following questions i-iv.**

i. When did you become aware of this type of support during your cancer journey? *(check one)*
   - After diagnosis
   - Prior to first prescription fill
   - When prescribed additional medications (e.g., supportive care agents)
   - After change to treatment plan
   - After change in employment status
   - After change in insurance status
   - Following Medicare enrollment
   - I was unaware of this type of support
   - Other

*(if other,)* Specify

ii. How did this assistance affect your cost? *(check one)*
    - It reduced the cost a little
    - It reduced the cost somewhat
    - It reduced the cost a lot
    - It eliminated the cost
iii. Did someone working with your doctor’s practice help you obtain this assistance?

*(check one)*

- Yes
- No

*(If yes)*, who helped you?

- Financial navigator / counselor *(check one)*
  - Yes
  - No
- Social worker *(check one)*
  - Yes
  - No
- Billing staff *(check one)*
  - Yes
  - No
- Physician office / clinic staff (e.g., doctors, nurses) *(check one)*
  - Yes
  - No
- Pharmacy staff *(check one)*
  - Yes
  - No
- I completed it on my own *(check one)*
  - Yes
  - No
- An application was not required *(check one)*
  - Yes
  - No
- Other *(check one)*
  - Yes
  - No

*(If yes,)* specify:

iv. How did you obtain the prescription medicines (e.g., co-pay assistance programs, pharmaceutical manufacturer patient assistance programs, etc.)?

- I received the medicines for free *(check one)*
  - Yes
  - No
- Drug discount card *(check one)*
  - Yes
  - No
- Drug coupon *(check one)*
  - Yes
  - No
- Pharmaceutical manufacturer co-pay assistance program *(check one)*
  - Yes
  - No
- National patient support organization or foundation co-pay assistance program *(check one)*
  - Yes
  - No
- Hospital / clinic *(check one)*
  - Yes
  - No
- Local community organization *(check one)*
  - Yes
  - No
- Other *(check one)*
  - Yes
  - No

*(If yes,)* specify:

b. Insurance premiums and medical care (e.g., hospital bills) *(check one)*
   - Yes
   - No

***(If yes),* answer the following questions i-iii.**

i. When did you become aware of this type of support during your cancer journey? *(check one)*
   - After a change in employment status
   - After a change in insurance status
   - After a change in insurance provider
   - After a change in insurance plan
   - Following Medicare enrollment
   - After my premiums increased
   - When I was unable to afford my out-of-pocket costs
   - I was unaware of this type of support
   - Other

*(if other, specify)*

ii. How did this assistance affect your cost? *(check one)*
    - It reduced the cost a little
    - It reduced the cost somewhat
    - It reduced the cost a lot
    - It eliminated the cost
iii. Did someone working with your doctor’s practice help you obtain this assistance? *(check one)*
     - Yes
     - No

*(If Yes),* who helped you?

- Financial navigator / counselor *(check one)*
  - Yes
  - No
- Social worker *(check one)*
  - Yes
  - No
- Billing staff *(check one)*
  - Yes
  - No
- Physician office / clinic staff (e.g., doctors, nurses) *(check one)*
  - Yes
  - No
- Pharmacy staff *(check one)*
  - Yes
  - No
- I completed it on my own *(check one)*
  - Yes
  - No
- An application was not required *(check one)*
  - Yes
  - No
- Other *(check one)*
  - Yes
  - No

*(If yes,)* specify:

c. Transportation to and from medical appointments *(check one)*
   - Yes
   - No

***(If yes),* answer the following questions i-iii.**

i. When did you become aware of this type of support during your cancer journey? *(check one)*
   - When I was unable to drive myself to an appointment
   - When family / friends were unable to drive me to an appointment
   - When I was unable to afford the cost of transportation (e.g., bus fare, taxi)
   - When I had to reschedule an appointment to ensure transportation (e.g., accommodate schedules of those driving me)
   - When I had to cancel an appointment due to lack of transportation
   - I was unaware of this type of support
   - Other

*(if other, specify)*

ii. How did this assistance affect your cost? *(check one)*
    - It reduced the cost a little
    - It reduced the cost somewhat
    - It reduced the cost a lot
    - It eliminated the cost
iii. Did someone working with your doctor’s practice help you obtain this assistance? *(check one)*
     - Yes
     - No

*(If Yes)*, who helped you?

- Financial navigator / counselor *(check one)*
  - Yes
  - No
- Social worker *(check one)*
  - Yes
  - No
- Billing staff *(check one)*
  - Yes
  - No
- Physician office / clinic staff (e.g., doctors, nurses) *(check one)*
  - Yes
  - No
- Pharmacy staff *(check one)*
  - Yes
  - No
- I completed it on my own *(check one)*
  - Yes
  - No
- An application was not required *(check one)*
  - Yes
  - No
- Other *(check one)*
  - Yes
  - No

*(If yes,)* specify:

d. Living expenses (e.g., groceries, utilities, rent) *(check one)*
   - Yes
   - No

***(If yes),* answer the following questions i-iii.**

i. when did you become aware of this type of support during your cancer journey? *(check one)*
   - After a change in employment status
   - After a change in insurance status
   - Following Medicare enrollment
   - When I fell behind with my bills
   - When I cut back on expenses
   - When I no longer could afford my rent / mortgage
   - When family / friends were unable to help me
   - I was unaware of this type of support
   - Others

(*if other, specify)*

ii. How did this assistance affect your cost? *(check one)*
    - It reduced the cost a little
    - It reduced the cost somewhat
    - It reduced the cost a lot
    - It eliminated the cost
iii. Did someone working with your doctor’s practice help you obtain this assistance? *(check one)*
     - Yes
     - No

*(If Yes)*, who helped you?

- Financial navigator / counselor *(check one)*
  - Yes
  - Not
- Social worker *(check one)*
  - Yes
  - No
- Billing staff *(check one)*
  - Yes
  - No
- Physician office / clinic staff (e.g., doctors, nurses) *(check one)*
  - Yes
  - No
- Pharmacy staff *(check one)*
  - Yes
  - No
- I completed it on my own *(check one)*
  - Yes
  - No
- An application was not required *(check one)*
  - Yes
  - No
- Other
  - Yes
  - No

*(If yes,)* specify:

**Support from family and friends for specific purposes**

Now I would like to ask you about support you have received from family and friends [this includes neighbors, coworkers (e.g., unions), church, internet fundraising campaigns]

*(NOTE: if response is “I don’t know,” “Not answered,” or “Not asked” please type the response in the text box provided when applicipable)*

4. Have you received money from family and friends in the past 12 months to help pay for **medical bills**? *(check one)* ***(If “no,” go to question 6)***
   - Yes
   - No
5. What is the total dollar amount of financial support for medical bills that you received in the past 12 months?
6. Have you received money from family and friends in the past 12 months to help pay for non- medical expenses (e.g., groceries, utilities, childcare, rent)? *(check one)* **(*If “no,” go to question 8)***
   - Yes
   - No
7. What is the total dollar amount of financial support for non-medical expenses that you received in the past 12 months?
8. In the past 12 months have your family and friends provided other kinds of support?

#### (If “no,” go to question 10)

- Yes
- No

9. What kinds of support?
1. Picking up prescriptions for you *(check one)*
   - Yes
   - No
2. Transportation to and from medical appointments *(check one)*
   - Yes
   - No
3. Coming to clinic appointments and waiting with you *(check one)*
   - Yes
   - No
4. Visiting you in the hospital *(check one)*
   - Yes
   - No
5. Buying groceries *(check one)*
   - Yes
   - No
6. Running errands *(check one)*
   - Yes
   - No
7. Providing childcare *(check one)*
   - Yes
   - No
8. Cleaning and laundry *(check one)*
   - Yes
   - No
9. Yard work and home repairs *(check one)*
   - Yes
   - No
10. Bringing over food *(check one)*
    - Yes
    - No
11. Keeping you company at home *(check one)*
    - Yes
    - No
12. Other
    - Yes
    - No

*(If yes,)* specify:

**Time burden of financial aspects of cancer treatment**

Now I would like to ask you about the amount of time you or your family members spending identifying way to pay for treatment.

10. How much has trying to find ways to pay for your treatment interfered with daily life for you or your family? *(check one)*
    - Not at all
    - A little bit
    - Somewhat
    - Quite a bit
    - Very much

**Challenges of paying for treatment** *(if response is “I don’t know,” “Not answered,” or “Not asked” please type the response in the text box provided)*

11. What have been the three most challenging things about obtaining money to pay for treatment?
12. If you were talking to another person who needed to obtain money to pay for treatment, what would you recommend to them?

**Part 3: Patient Sociodemographic Indicators**

Now I would like to ask you some questions about your employment, income, and debt.

*(NOTE: if response is “I don’t know,” “Not answered,” or “Not asked” please type the response in the text box provided when applicipable)*

**Patient Employment Status**

1. Do you currently have a job for pay or own a business? *(check one)* ***(If “no,” go to question 4)***
   - Yes
   - No
2. Often the actual number of hours people work is different from the number of hours on which their salaries are based. How many hours per week do you usually work at your main job? Include all the hours you usually spend working on this job, including paid sick leave except for any unpaid travel to and from the job.

Enter number of hours

3. Please think about the time you have worked this year, including paid vacation, sick leave, or other paid leave. How many weeks did you work for pay either full or part time? If worked less than one week, enter ‘1’ for number of weeks

Enter number of weeks

4. What is the **main** reason you do not work currently? *(check one)*
   - Could not find work
   - Retired
   - Unable to work because ill/disabled
   - On temporary layoff
   - Taking care of home or family
   - Wanted some time off
   - Waiting to start new job
   - Other

*(if other, specify)*

5. Do you have health insurance through a job you hold or through your union? By this, I mean insurance which pays for hospital bills, doctor bills, or other health expenses. *(check one)*
   - Yes
   - No

**Household monthly income, assets**

The next set of questions are about household income and assets.

6. Over the last year, what was the total income of the household you live in? *(check one)*
   - Less than $20,000
   - $20,000-39,999
   - $40,000-59,999
   - $60,000-79,999
   - $80,000-99,999
   - $100,000 or more
   - Don’t know
7. Does anyone in your immediate family (you, your spouse or significant other, your child, your sibling, your parent) own the home you are living in? *(check one)* ***(If “no,” go to question 12)***
   - Yes
   - No
8. *(If Yes),* Who in your family owns your home?
a. You *(check one)*
   - Yes
   - No
b. Your spouse or significant other *(check one)*
   - Yes
   - No
c. Someone else in your family *(check one)*
   - Yes
   - No
9. Approximately what is the value of your home if it was sold today?	*(check one)*
   - $0-25,000
   - $25,001-50,000
   - $50,001-100,000
   - $100,001-250,000
   - $250,001-500,000
   - $500,001or more
10. Are there any mortgages or other loans outstanding on this home? *(check one)*

#### (If “no,” go to question 12)

- Yes
- No

11. How much is currently owed on these mortgages or loans? *(check one)*
    - $0-25,000
    - $25,001-50,000
    - $50,001-100,000
    - $100,001-250,000
    - $250,001-500,000
    - $500,001or more

**Household debt**

The next set of questions are about financial debt.

12. Do you or anyone in the family have other debts such as credit card balances, car loans, debts owed to medical providers, life insurance policy loans, loans from relatives and so forth? *(check one)* ***(If “no,” go to question 17)***
    - Yes
    - No
13. What is the total amount owed on this other debt? *(check one)*
    - $0-25,000
    - $25,001-50,000
    - $50,001-100,000
    - $100,001-250,000
    - $250,001-500,000
    - $500,001 or more
14. Have you ever been sent to collections because of debts you were unable to pay on time or at all?

*(check one)*

- Yes
- No

15. Have you ever filed for bankruptcy because of debts you were unable to pay? *(check one)* ***(If “no” go to question 17)***
    - Yes
    - No
16. What was the most recent year in which you filed for bankruptcy? *(if response is “I don’t know,” “Not answered,” or “Not asked” please type the response in the text box provided)*

**Patient insurance**

Now I would like to ask you about your health insurance.

17. Do you currently have health insurance that covers the following services: ***(if no to a-c, go to question 20)***
a. Outpatient care *(check one)*
   - Yes
   - No
b. Hospital *(check one)*
   - Yes
   - No
c. Prescription coverage *(check one)*
   - Yes
   - No
18. Which type of health insurance coverage do you now have?

- Private Insurance *(check one)*
  - Yes
  - No
- Medicare *(check one)*
  - Yes
  - No
- Medicare and Private Insurance *(check one)*
  - Yes
  - No
- Medicaid *(check one)*
  - Yes
  - No
- Medicaid and Medicare *(check one)*
  - Yes
  - No
- Military or Veterans Sponsored, Not Otherwise Specified (NOS) *(check one)*
  - Yes
  - No
- Military Sponsored (including CHAMPUS & TRICARE) *(check one)*
  - *Yes*
  - No
- Veterans Sponsored *(check one)*
  - Yes
  - No
- Self Pay (no insurance) *(check one)*
  - Yes
  - No
- No Means of Payment (no insurance) *(check one)*
  - Yes
  - No
- Other *(check one)*
  - Yes
  - No

*(If yes,)* specify:

- Unknown *(check one)*
  - Yes
  - No

19. What is your household annual deductible for medical care for your insurance plan? *(Check one)*
    - Less than $1300
    - $1,301 to $2,600
    - More than $2600

**Financial literacy**

This next set of questions are a bit different. I am going to ask you five math questions about money and interest rates. You can take your time answering them, there is no rush.

20. Suppose you had $100 in a savings account and the interest rate was 2% per year. After 5 years, how much do you think you would have in the account if you left the money to grow? *(check one)*
    - More than $102
    - Exactly $102
    - Less than $102
21. Suppose you had $100 in a savings account and the interest rate is 20% per year and you never withdraw money or interest payments. After 5 years, how much would you have in this account in total? *(check one)*
    - More than $200
    - Exactly $200
    - Less than $200
22. Imagine that the interest rate on your savings account was 1% per year and inflation was 2% per year. After 1 year, how much would you be able to buy with the money in this account? *(check one)*
    - More than today
    - Exactly the same
    - Less than today
23. Assume a friend inherits $10,000 today and his sibling inherits $10,000 three years from now. Who is richer because of the inheritance? *(check one)*
    - My friend
    - His sibling
    - They are equally rich
24. Suppose that in the year 2020, your income has doubled and prices of all goods have doubled too. In 2020, how much will you be able to buy with your income? *(check one)*
    - More than today
    - The same
    - Less than today

**Patient demographics**

Now I would like to ask some questions about you. *(if response is “I don’t know,” “Not answered,” or “Not asked” please type the response in the text box provided when applicable)*

25. What is your zip code?
26. What is your sex? *(check one)*
    - Female
    - Male
27. Are you: *(check one)*
    - Hispanic or Latino
    - Not Hispanic or Latino
28. What is your race?

- American Indian or Alaska Native *(check one)*
  - Yes
  - No
- Asian *(check one)*
  - Yes
  - No
- Black or African American *(check one)*
  - Yes
  - No
- Native Hawaiian or other Pacific Islander *(check one)*
  - Yes
  - No
- White *(check one)*
  - Yes
  - No
- Not reported *(check one)*
  - Yes
  - No
- Unknown *(check one)*
  - Yes
  - No

29. Were you born in the United States? *(check one)* ***(If yes, go to question 31)***
    - Yes
    - No
30. How many years have you lived in the United States?	Years
31. What is your first language? *(check one)* ***(If ‘English’ go to question 33)***
    - English
    - Spanish
    - Other,
      - *(if other, specify)*
32. How well do you speak English? *(check one)*
    - Very Well
    - Well
    - Not well
    - Not at all
33. What is your marital status? *(check one)*
    - Married
    - Widowed
    - Divorced
    - Separated
    - Never Married

**Patient Household Composition**

34. How many people currently live in your **main household** (including you)?
35. How many of them are younger than 18 years old?
36. How many of them are 62 years or older?

**Patient Education Status**

37. What is the highest educational degree you obtained as of December 31, 2018? *(check one)*
    - GED
    - High School Diploma
    - Associate’s Degree
    - Bachelor’s Degree
    - Master’s Degree
    - No Degree/Diploma
    - Other

**Part 4: Health and Well-being**

Now I would like to ask you a series of questions about your health and well-being. This is the last part of the interview today. We are almost done.

**PROMIS Global Health 10**

1. In general, would you say your health is *(check one)*
   - Excellent
   - Very Good
   - Good
   - Fair
   - Poor
2. In general, would you say your quality of life is*: (check one)*
   - Excellent
   - Very Good
   - Good
   - Fair
   - Poor
3. In general, how would you rate your physical health: *(check one)*
   - Excellent
   - Very Good
   - Good
   - Fair
   - Poor
4. In general, how would you rate your mental health, including your mood and ability to think: *(check one)*
   - Excellent
   - Very Good
   - Good
   - Fair
   - Poor
5. In general, how would you rate your satisfaction with your social activities and relationships: *(check one)*
   - Excellent
   - Very Good
   - Good
   - Fair
   - Poor
6. In general, please rate how well you carry out your usual social activities and roles (This includes activities at home, at work and in your community, and responsibilities as a parent, child, spouse, employee, friend, etc.) *(check one)*
   - Excellent
   - Very Good
   - Good
   - Fair
   - Poor
7. To what extent are you able to carry out your everyday physical activities such as walking, climbing chairs, carrying groceries or moving a chair? *(check one)*
   - Completely
   - Mostly
   - Moderately
   - A Little
   - Not At All
8. In the past 7 days, how often have you been bothered by emotional problems such as feeling anxious, depressed, or irritable? *(check one)*
   - Never
   - Rarely
   - Sometimes
   - Often
   - Always
9. In the past 7 days, how would you rate your fatigue on average? *(check one)*
   - None
   - Mild
   - Moderate
   - Severe
   - Very Severe
10. How would you rate your pain on average, where 0 means “no pain” and 10 means “the worst imaginable pain”? (response options: integers 0 through 10) (check one)
    - 0
    - 1
    - 2
    - 3
    - 4
    - 5
    - 6
    - 7
    - 8
    - 9
    - 10

**Self-reported health (EQ-5D 5 items)**

EQ-5D DESCRIPTIVE SYSTEM: INTRODUCTION

First I am going to read out some questions. Each question has a choice of five answers. Please tell me which answer best describes your health TODAY. Do not choose more than one answer in each group

of questions.

*(Note to interviewer: it may be necessary to remind the respondent regularly that the timeframe is*

*TODAY. It may also be necessary to repeat the questions verbatim)*

11. MOBILITY

First I'd like to ask you about mobility. Would you say that: *(check one)*

- You have no problems walking?

- You have slight problems walking?

- You have moderate problems walking?

- You have severe problems walking?

- You are unable to walk?

12. SELF-CARE

Next I'd like to ask you about self-care. Would you say that: *(check one)*

- You have no problems washing or dressing yourself?

- You have slight problems washing or dressing yourself?

- You have moderate problems washing or dressing yourself?

- You have severe problems washing or dressing yourself?

- You are unable to wash or dress yourself?

13. USUAL ACTIVITIES

Next I'd like to ask you about your usual activities, for example work, study, housework, family or leisure activities. Would you say that: *(check one)*

- You have no problems doing your usual activities?

- You have slight problems doing your usual activities?

- You have moderate problems doing your usual activities?

- You have severe problems doing your usual activities?

- You are unable to do your usual activities?

14. PAIN / DISCOMFORT

Next I'd like to ask you about pain or discomfort. Would you say that: *(check one)*

- You have no pain or discomfort?

- You have slight pain or discomfort?

- You have moderate pain or discomfort?

- You have severe pain or discomfort?

- You have extreme pain or discomfort?

15. ANXIETY / DEPRESSION

Finally I'd like to ask you about anxiety or depression. Would you say that *: (check one)*

- You are not anxious or depressed?

- You are slightly anxious or depressed?

- You are moderately anxious or depressed?

- You are severely anxious or depressed?

- You are extremely anxious or depressed?

EQ VAS: INTRODUCTION

Now, I would like to ask you to say how good or bad your health is TODAY.

I'd like you to try to picture in your mind a scale that looks a bit like a thermometer. Can you do that? The best health you can imagine is marked 100 (one hundred) at the top of the scale and the worst health you can imagine is marked 0 (zero) at the bottom.

EQ VAS: TASK

I would now like you to tell me the point on this scale where you would put your health today. *(if response is “I don’t know,” “Not answered,” or “Not asked” please type the response in the text box provided)*

16. Your health today

**Brief Appraisal Inventory**

In this last section, we want to understand the things that were on your mind as you completed this survey today. For example, we could ask people whether they have been thinking about getting a new car. Some people may have thought about needing a car often throughout the survey while others may not have thought about cars at all. There are also people who do not even drive, so the idea of getting a car would not be applicable to them. For each question below, please indicate how frequently it was on your mind as you completed the interview today, or if the question does not apply to you.

During the survey today, how often did you think about…

16. Maintaining a positive outlook, even when things are going badly *(check one)*
    - Always
    - Often
    - Sometimes
    - Rarely
    - Never
    - Not applicable
    - Refused to answer
17. Achieving a calmer, more peaceful or healthier lifestyle *(check one*)
    - Always
    - Often
    - Sometimes
    - Rarely
    - Never
    - Not applicable
    - Refused to answer
18. Solving problems that you have with health care or the health care system *(check one)*
    - Always
    - Often
    - Sometimes
    - Rarely
    - Never
    - Not applicable
    - Refused to answer
19. Impressions and assumptions that others have about you because of your health *(check one)*
    - Always
    - Often
    - Sometimes
    - Rarely
    - Never
    - Not applicable
    - Refused to answer
20. How you compare to others whose health does not limit them *(check one)*
    - Always
    - Often
    - Sometimes
    - Rarely
    - Never
21. Being free of money problems *(check one)*
    - Always
    - Often
    - Sometimes
    - Rarely
    - Never
    - Not applicable
    - Refused to answer
22. Increasing your volunteer work to help others in your community *(check one)*
    - Always
    - Often
    - Sometimes
    - Rarely
    - Never
    - Not applicable
    - Refused to answer
23. Growing spiritually *(check one)*
    - Always
    - Often
    - Sometimes
    - Rarely
    - Never
    - Not applicable
    - Refused to answer
24. Staying active and productive *(check one)*
    - Always
    - Often
    - Sometimes
    - Rarely
    - Never
    - Not applicable
    - Refused to answer
25. Resolving problems in your living situation *(check one)*
    - Always
    - Often
    - Sometimes
    - Rarely
    - Never
    - Not applicable
    - Refused to answer
26. Preparing your family for the ups and downs of your health condition *(check one)*
    - Always
    - Often
    - Sometimes
    - Rarely
    - Never
    - Not applicable
    - Refused to answer
27. Setting conflicts with people in your life *(check one)*
    - Always
    - Often
    - Sometimes
    - Rarely
    - Never
    - Not applicable
    - Refused to answer
28. Finding romance *(check one)*
    - Always
    - Often
    - Sometimes
    - Rarely
    - Never
    - Not applicable
    - Refused to answer
29. Learning to accept yourself as you are *(check one)*
    - Always
    - Often
    - Sometimes
    - Rarely
    - Never
    - Not applicable
    - Refused to answer
30. Things that do not usually come to mind, except because of this survey *(check one)*
    - Always
    - Often
    - Sometimes
    - Rarely
    - Never
    - Not applicable
    - Refused to answer
31. Having dreams and goals that are different from most people your own age *(check one)*
    - Always
    - Often
    - Sometimes
    - Rarely
    - Never
    - Not applicable
    - Refused to answer
32. Spending more time with yoru family before your health worsens *(check one)*
    - Always
    - Often
    - Sometimes
    - Rarely
    - Never
    - Not applicable
    - Refused to answer
33. Remaining independent and being able to get around on your own *(check one)*
    - Always
    - Often
    - Sometimes
    - Rarely
    - Never
    - Not applicable
    - Refused to answer
34. Being rid of obligations and responsibilities *(check one)*
    - Always
    - Often
    - Sometimes
    - Rarely
    - Never
    - Not applicable
    - Refused to answer
35. Trying not to complain about your health to others *(check one)*
    - Always
    - Often
    - Sometimes
    - Rarely
    - Never
    - Not applicable
    - Refused to answer
36. How you compare to others facing similar health issues *(check one)*
    - Always
    - Often
    - Sometimes
    - Rarely
    - Never
    - Not applicable
    - Refused to answer
37. Increasing your travel for leisure or for visiting with people *(check one)*
    - Always
    - Often
    - Sometimes
    - Rarely
    - Never
    - Not applicable
    - Refused to answer
38. Accomplishing new goals at work *(check one)*
    - Always
    - Often
    - Sometimes
    - Rarely
    - Never
    - Not applicable
    - Refused to answer

**Closing Question:**

I have asked you a lot of questions today.

39. Is there anything else you think it’s important for us to know about financial challenges related to cancer treatment? *(if response is “I don’t know,” “Not answered,” or “Not asked” please type the response in the text box provided)*

**CONCLUDING PHONE INTERVIEW WITH PARTICIPANT**

1. Thank the patient: “This completes our interview. Thank you for taking the time to answer these questions today. The purpose of this study is to help us to better understand the financial impact of cancer and come up with ways to help patients avoid financial problems during treatment.”
2. Gift card: “We would like to mail you a $20 gift card to thank you for your participation in this study. We would also like to send you some information about organizations that can help find financial support to pay for your cancer care, organizations that can help you understand your disease and treatment options, and that can help you find professional and peer support. Would you be comfortable providing your mailing address, so that we can send you the gift card and information?”
   a. Yes: “Great, could you please provide your full street address? City? State? Zip Code?”
      i. Confirm address: “OK. I have [**repeat address**]. Is this correct?”
      ii. “Thank you. You should receive the gift card at this address within 4 weeks. Please contact Michelle at (919) 962-5378 if you do not receive it.”
   b. No/refuses gift card: “Ok. That’s fine.”
3. Connect patient to resource organizations: If you’d like, I can go ahead and give you the phone numbers today for two organizations that provide resources to individuals with [Chronic lymphocytic leukemia / myeloma]. The first organization is called the Leukemia & Lymphoma Society—this organization can help you find financial support, and they can help you understand your disease and treatment options and find professional and peer support for your cancer care. The second organization is called Cancer Care, and they provide free counseling services to anyone affected by cancer, and they also can help you find financial assistance to pay for your cancer care. Would you like me to give you the phone numbers to these organizations?”
   a. Yes:
      i. “For the Leukemia & Lymphoma Society, you can reach one of their Information Specialists at **1-800-955-4572**. Would you like me to repeat this number?”
      ii. “For Cancer Care, you can reach a social worker at **1-800-813-HOPE (4673).** Would you like me to repeat this number?”
   b. No: “OK.“
   c. If already contacted LLS or Cancer Care before: “OK, there are several other patient organizations that provide these types of services. Is there a particular type of resource that you are looking for?”
      i. Review resource list and provide phone numbers for one or two other organizations.
4. Confidentiality and questions: “As a reminder, everything we discussed today will be kept confidential. Do you have any questions about this study, now that we are finished with the interview?”

Conclude: “Thank you again for your time. Goodbye.
